# Two neurostructural subtypes: results of machine learning on brain images from 4,291 individuals with schizophrenia

**DOI:** 10.1101/2023.10.11.23296862

**Authors:** Yuchao Jiang, Cheng Luo, Jijun Wang, Lena Palaniyappan, Xiao Chang, Shitong Xiang, Jie Zhang, Mingjun Duan, Huan Huang, Christian Gaser, Kiyotaka Nemoto, Kenichiro Miura, Ryota Hashimoto, Lars T. Westlye, Genevieve Richard, Sara Fernandez-Cabello, Nadine Parker, Ole A. Andreassen, Tilo Kircher, Igor Nenadić, Frederike Stein, Florian Thomas-Odenthal, Lea Teutenberg, Paula Usemann, Udo Dannlowski, Tim Hahn, Dominik Grotegerd, Susanne Meinert, Rebekka Lencer, Yingying Tang, Tianhong Zhang, Chunbo Li, Weihua Yue, Yuyanan Zhang, Xin Yu, Enpeng Zhou, Ching-Po Lin, Shih-Jen Tsai, Amanda L. Rodrigue, David Glahn, Godfrey Pearlson, John Blangero, Andriana Karuk, Edith Pomarol-Clotet, Raymond Salvador, Paola Fuentes-Claramonte, María Ángeles Garcia-León, Gianfranco Spalletta, Fabrizio Piras, Daniela Vecchio, Nerisa Banaj, Jingliang Cheng, Zhening Liu, Jie Yang, Ali Saffet Gonul, Ozgul Uslu, Birce Begum Burhanoglu, Aslihan Uyar Demir, Kelly Rootes-Murdy, Vince D. Calhoun, Kang Sim, Melissa Green, Yann Quidé, Young Chul Chung, Woo-Sung Kim, Scott R. Sponheim, Caroline Demro, Ian S. Ramsay, Felice Iasevoli, Andrea de Bartolomeis, Annarita Barone, Mariateresa Ciccarelli, Arturo Brunetti, Sirio Cocozza, Giuseppe Pontillo, Mario Tranfa, Min Tae M. Park, Matthias Kirschner, Foivos Georgiadis, Stefan Kaiser, Tamsyn E Van Rheenen, Susan L Rossell, Matthew Hughes, William Woods, Sean P Carruthers, Philip Sumner, Elysha Ringin, Filip Spaniel, Antonin Skoch, David Tomecek, Philipp Homan, Stephanie Homan, Wolfgang Omlor, Giacomo Cecere, Dana D Nguyen, Adrian Preda, Sophia Thomopoulos, Neda Jahanshad, Long-Biao Cui, Dezhong Yao, Paul M. Thompson, Jessica A. Turner, Theo G.M. van Erp, Wei Cheng, ENIGMA Schizophrenia Consortium, ZIB Consortium, Jianfeng Feng

**Author notes:** Corresponding Authors to: Jianfeng Feng.

## Abstract

Machine learning can be used to define subtypes of psychiatric conditions based on shared clinical and biological foundations, presenting a crucial step toward establishing biologically based subtypes of mental disorders. With the goal of identifying subtypes of disease progression in schizophrenia, here we analyzed cross-sectional brain structural magnetic resonance imaging (MRI) data from 4,291 individuals with schizophrenia (1,709 females, age=32.5 years±11.9) and 7,078 healthy controls (3,461 females, age=33.0 years±12.7) pooled across 41 international cohorts from the ENIGMA Schizophrenia Working Group, non-ENIGMA cohorts and public datasets. Using a machine learning approach known as Subtype and Stage Inference (SuStaIn), we implemented a brain imaging-driven classification that identifies two distinct neurostructural subgroups by mapping the spatial and temporal trajectory of gray matter (GM) loss in schizophrenia. Subgroup 1 (n=2,622) was characterized by an early cortical-predominant loss (ECL) with enlarged striatum, whereas subgroup 2 (n=1,600) displayed an early subcortical-predominant loss (ESL) in the hippocampus, amygdala, thalamus, brain stem and striatum. These reconstructed trajectories suggest that the GM volume reduction originates in the Broca’s area/adjacent fronto-insular cortex for ECL and in the hippocampus/adjacent medial temporal structures for ESL. With longer disease duration, the ECL subtype exhibited a gradual worsening of negative symptoms and depression/anxiety, and less of a decline in positive symptoms. We confirmed the reproducibility of these imaging-based subtypes across various sample sites, independent of macroeconomic and ethnic factors that differed across these geographic locations, which include Europe, North America and East Asia. These findings underscore the presence of distinct pathobiological foundations underlying schizophrenia. This new imaging-based taxonomy holds the potential to identify a more homogeneous sub-population of individuals with shared neurobiological attributes, thereby suggesting the viability of redefining existing disorder constructs based on biological factors.

## 1. Introduction

A key goal of biological psychiatry is to define biological subtypes of major mental disorders, based on objective measures derived from imaging and other biomarkers [1]. In psychiatry, redefining subtypes of psychiatric disorders based on neural mechanisms augmenting clinical behavioral criteria presents significant benefits. By using biological characteristics to more efficiently identify biologically homogeneous clinical cohorts, clinical trials could more effectively discern the biological effects of a given intervention. Artificial intelligence (AI) methods such as machine learning can be applied to brain imaging [2] to categorize individuals based on their profiles of brain metrics, and holds great potential for revealing the underlying neurobiological mechanisms associated with disorder subtypes [3].

Schizophrenia is one of the most severely disabling psychiatric disorders with a life-time prevalence of 1%; it affects approximately 26 million people worldwide [4]. The etiology of schizophrenia is still not fully understood. Current knowledge implicates multiple neurobiological mechanisms and pathophysiologic processes [5, 6]. Furthermore, people diagnosed with schizophrenia show a substantial heterogeneity in clinical symptoms [7], disease progression [8], treatment response [9], and other biological markers [10, 11]. In addition, currently available treatments are not aligned with specific pathophysiological pathways/targets, which limits effectiveness of treatment selection [12]. Establishing a new taxonomy by identifying distinct subtypes based on neurobiological data could help resolve some of these heterogeneity-induced challenges.

Machine learning algorithms are increasingly used to subtype brain disorders [13-16]. Prior studies have primarily focused on grouping individuals into distinct categories without considering disease progression [17, 18]. A major obstacle to identifying distinct patterns of neuro-pathophysiological progression (referred to as progression subtypes) stems from the lack of sufficient longitudinal data covering the lifespan of the disorder. Recently, a novel data-driven machine learning approach known as Subtype and Stage Inference (SuStaIn) was introduced [19]. SuStaIn uses a large number of cross-sectional observations, derived from single time-point MRI scans, to identify clusters (subtypes) of individuals with common trajectory of disease progression (i.e., the sequence of MRI abnormalities across different brain regions) in brain disorders [20-22]. By applying SuStaIn to MRI data from individuals with schizophrenia, primarily collected from the Chinese population, we found that the progression of gray matter loss in schizophrenia can be better characterized through two distinct phenotypes: one characterized by a cortical-predominant progression, originating in the Broca’s area/fronto-insular cortex, and another marked by a subcortical-predominant progression, starting in the hippocampus [22]. Such brain-based taxonomies may reflect neurostructural subtypes with shared pathophysiological foundations, with relevance for neurobiological classification [22]. However, the generalisability of the two neurostructural subtypes to diverse populations outside of China, and external validation of the subgrouping is required before applying this knowledge to stratify clinical trials.

The Enhancing Neuro Imaging Genetics through Meta-Analysis (ENIGMA, http://enigma.ini.usc.edu) consortium is dedicated to conducting large-scale analyses by pooling brain imaging data from research teams worldwide, using standardized image processing protocols. Previously, ENIGMA published findings revealing thinner cerebral cortex, smaller surface area, and altered subcortical volumes in schizophrenia compared to controls [23, 24]. Here, we included structural MRI data obtained from 4,291 individuals diagnosed with schizophrenia and 7,078 healthy controls from 41 international cohorts from ENIGMA schizophrenia groups worldwide and other non-ENIGMA datasets (**Supplementary Table 1**). The large sample size allowed us to conduct systematic and comprehensive analyses to verify the reproducibility and generality of neurostructural subtypes of schizophrenia across regions/locations and disease stages. This study’s aims were: (1) to validate the two neurostructural subtypes with distinct trajectories of neuro-pathophysiological progression in schizophrenia, (2) to verify the reproducibility and generality of the neurostructural subtypes, in subsamples across the world and across disease stages, and (3) to characterize subtype-specific signatures in terms of neuroanatomy and clinical symptomatic trajectory.

Together, these analyses aim to create a new, easily accessible (with a single anatomical MRI), interpretable (based on ‘progressive’ pathology) and robustly generalizable (across ethnic, sex and language differences) taxonomy of subtypes that share common neurobiological mechanisms in schizophrenia. If proven effective, other complex neuropsychiatric disorders with high heterogeneity [25, 26], such as major depressive disorder, autism spectrum disorder, and obsessive-compulsive disorder, could also benefit from such a subtyping paradigm. This has the potential to transition the field of psychiatry from syndrome-based to both syndrome- and biology-based stratifications of mental disorders.

## 2. Results

### 2.1 Two biotypes with distinct pathophysiological progression trajectories

Distinct patterns of spatiotemporal progression of pathophysiological progression were identified using SuStaIn, based on cross-sectional MRI data from 4,222 individuals diagnosed with schizophrenia (1,683 females, mean age=32.4±11.9 years) and 7,038 healthy subjects (3,440 females, mean age=33.0±12.6 years) (**Table.1**). A 2-fold cross-validation procedure resulted in an optimal number of K=2 clusters (subtypes) as determined by the largest Dice coefficient (**Fig.1a**), indicating the best consistency of the subtype labeling across all individuals between for a model in two independent schizophrenia populations. **Fig.1b** shows that only 1.2% of people were moved from subtype 1 to subtype 2, and 7.5% were moved from subtype 2 to subtype 1, indicating that 91.3% of individuals’ subtype labels were consistent between the SuStaIn classifications from two non-overlapping data folds. These findings suggest the presence of two stable schizophrenia biotypes with distinct ‘trajectories’ of pathophysiological progression (here we put trajectory in quotes as the typical sequence of disease progression is reconstructed from cross-sectional data).

**Figure 1.**
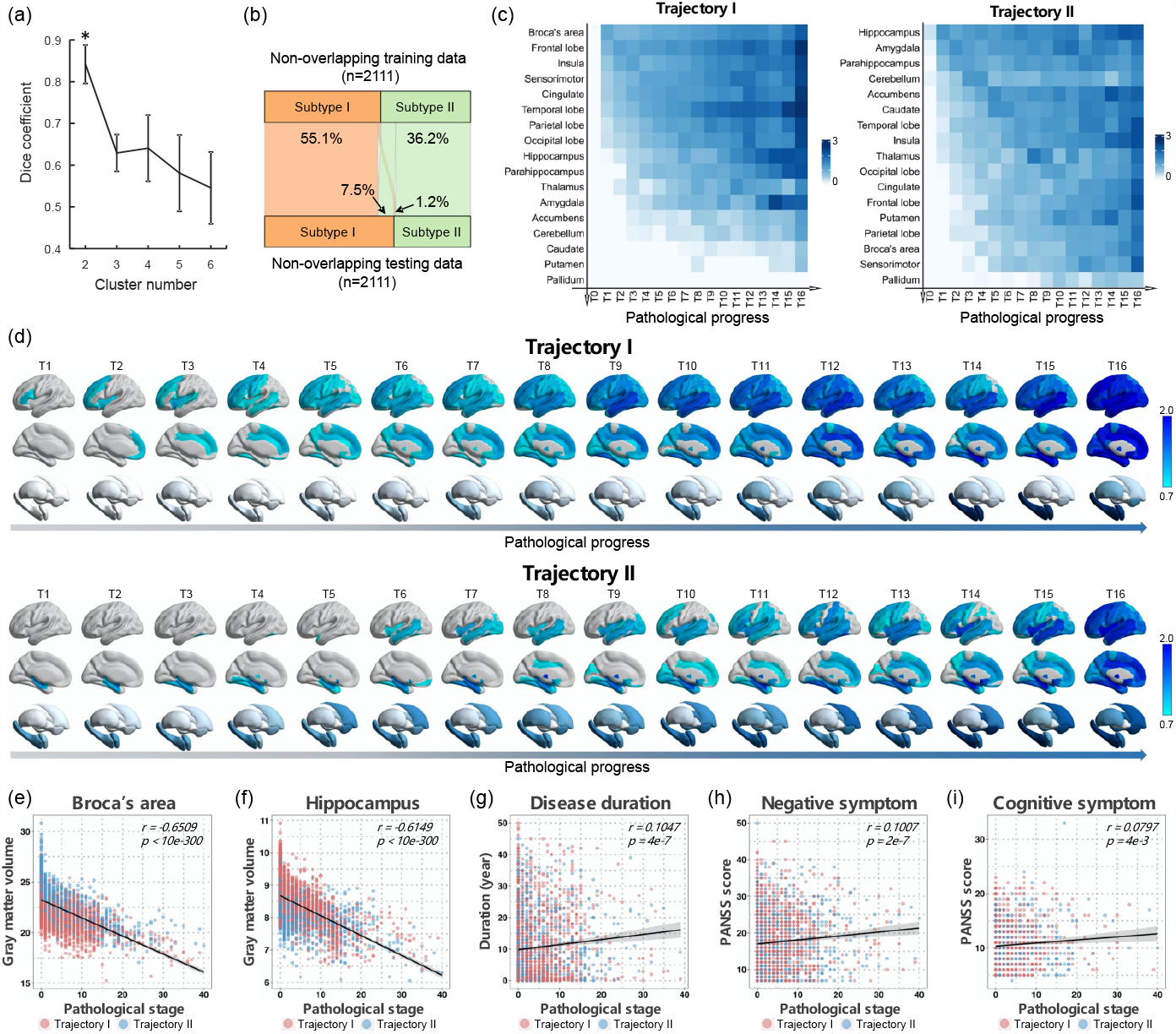
Two pathophysiological progression trajectories in schizophrenia. (**a**) Dice coefficient indicates that K=2 is the optimal number of subtypes with best consistency of the subtype labeling between two independent schizophrenia populations using non-overlap 2-folds cross-validation procedure. Data are presented as mean values +/-SD. (**b**) The proportion of individuals whose subtype labels keep consistent by non-overlap cross-validation procedure. (**c**) Sequences of regional volume loss across seventeen brain regions for each ‘trajectory’ via SuStaIn are shown in y-axis. The heatmap shows regional volume loss in which biomarker (y-axis) in a particular ‘temporal’ stage (T0-T16) in the trajectory (x-axis). The Color bar represents the degree of gray matter volume (GMV) loss in schizophrenia relative to healthy controls (i.e., z score). (**d**) Spatiotemporal pattern of pathophysiological ‘trajectory’. The z-score images are mapped to a glass brain template for visualization. Spatiotemporal pattern of gray matter loss displays a progressive pattern of spatial extension along with later ‘temporal’ stages of pathological progression, that is distinct between trajectories. (**e-f**) Pathological stages of SuStaIn are correlated with reduced gray matter volume of Broca’s area and hippocampus. (**g-i**) Pathological stages of SuStaIn are correlated with longer disease duration, worse negative symptoms and worse cognitive symptoms.

Region of interest (ROI)-wise gray matter volume (GMV) z-scores, at each stage of the ‘trajectory’ for each subtype, show the sequence of regional volume loss across the 17 brain regions for each ‘trajectory’ (**Fig.1c**). To visualize the spatiotemporal pattern of each ‘trajectory’, z-score whole brain images were mapped to a glass brain template (**Fig.1d**). These maps show a progressive pattern of spatial expansion along with later ‘temporal’ stages of pathological progression distinct for each ‘trajectory’. Specifically, ‘trajectory’ 1 displayed an ‘early cortical-predominant loss’ biotype. It was characterized by an initial reduction in Broca’s area, followed by adjacent fronto-insular regions, then extending to the rest of the neocortex, and finally to the subcortex (**Fig.1d**). Conversely, ‘trajectory’ 2 exhibited an ‘early subcortical-predominant loss’ biotype where volume loss began in the hippocampus, spread to the amygdala and parahippocampus, and then extended to the accumbens and caudate before affecting the cerebral cortex (**Fig.1d**). The two ‘trajectories’ were highly consistent with our previous findings in a predominantly Chinese schizophrenia cohort [22]. The phenotypic subtypes, based on the different pathophysiological ‘trajectories’, are thus replicated in a large cross-geography sample, confirming the presence of two different neuropathological pathways with different anatomical origins in schizophrenia [22].

### 2.2 Trajectories are repeated in first-episode and medication-naÏve samples

The sample size of this study was large enough to allow further exploratory analyses to identify pathophysiological progression trajectories in more homogeneous subsamples of schizophrenia. Here, we re-estimated the SuStaIn ‘trajectories’ based on a subsample of data from individuals with first-episode schizophrenia with illness duration less than two years (N=1,122; 513 females, mean age=25.4 ± 8.6 years), and a subsample of medication-naÏve individuals with schizophrenia (N=718, 353 females, mean age=23.7± 7.8 years) (**Supplementary Table 3**). In both subsamples, we replicated the two ‘trajectories’ with either the Broca’s area or the hippocampus as the sites of origin (**Extended Data Fig.1**), indicating that the two initiating regions - ranking ahead other regional deficits - are the pathological effects of the disease itself, rather than medication-induced effects. Broca’s area and the hippocampus may therefore be candidate targets for intervention in schizophrenia, as these two brain regions were affected early in the disease process.

### 2.3 Trajectories are reproducible for samples from different parts of the world

To examine whether the ‘trajectories’ were reproducible for samples from different parts of the world, we divided all samples into several sub-cohorts based on where the samples were obtained (**Extended Data Fig.5**). Here, samples from China, Japan, South Korea and Singapore were classified into the East Asian ancestry (EAS) cohort. Samples from Europe, the United States, Canada and Australia were classified into the European ancestry (EUR) cohorts (**Supplementary Table 4**). In addition, Chinese, Japanese, European and North American cohorts were further classified by their site locations by their site locations in terms of geographic distribution (**Supplementary Table 4**). Such a division was based on the similar ethnic or environmental factors for each country, region, or continent and the size of subsample, which need to be sufficient to conduct a reliable inference of the SuStaIn trajectory. We found that two ‘trajectories’ (the optimal number was also K=2, which separately re-estimated in each cohort) - with Broca’s area leading and the hippocampus leading - were also repeated in EAS (**Fig.2a**) and EUR (**Fig.2b**) cohorts. In addition, the spatiotemporal pattern of each ‘trajectory’ showed strong, significant correlations between the EAS and EUR cohorts (‘trajectory’ 1, *r*=0.948, *p*<0.001; ‘trajectory’ 2, *r*=0.842, *p*<0.001; Spearman correlation test). This high level of similarity in the trajectories was also observed between cohorts from other locations (**Fig.2c**). This suggests that the two biotypes with distinct ‘trajectories’ of pathophysiological progression in schizophrenia are robust, and their classification patterns are independent of macro-environmental or ethnogenetic factors.

**Figure 2.**
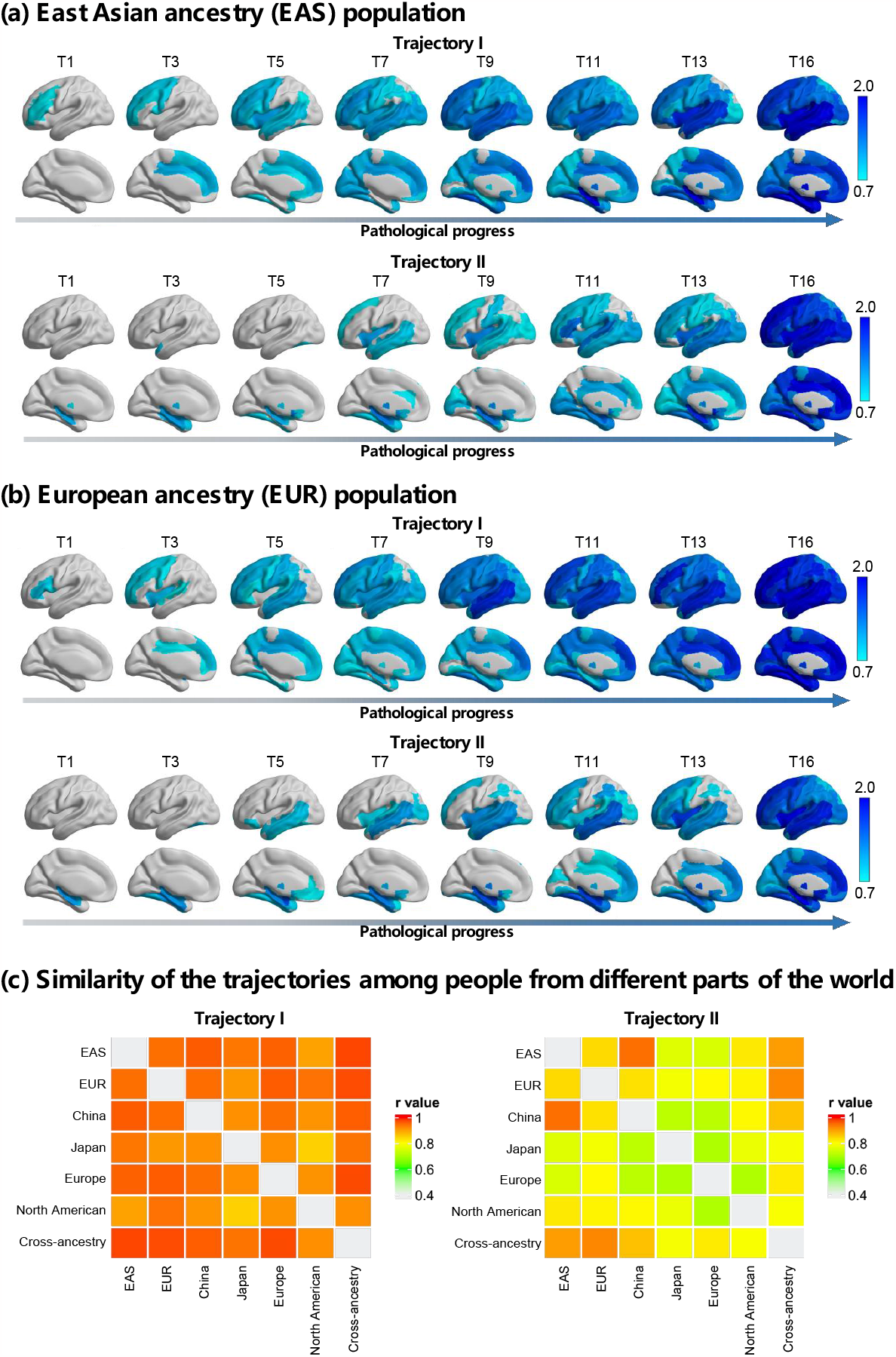
Trajectories are reproducibility for samples from different locations of the world. Two sets of ‘trajectories’ are separately derived from two non-overlapping location cohorts, that are (**a**) East Asian ancestry (EAS) cohort, and (**b**) European ancestry (EUR) cohort. The Color bar represents the degree of gray matter volume (GMV) loss in schizophrenia relative to healthy controls (i.e., z score). (**c**) The similarity of the spatiotemporal pattern of each ‘trajectory’ between any two of cohorts is shown by the heatmap. The color bar of the heatmap represents the similarity, which is quantified via the Spearman correlation coefficient between the trajectories from two cohorts. A total of six location cohorts are classified by where the sample locate at, including the EAS, EUR, China, Japan, Europe and North America. The whole sample is labelled as a cross-ancestry cohort.

### 2.4 Trajectories are associated with neurophysiological, pathological and neuropsychological progressions in schizophrenia

The SuStaIn calculated the probability of each patient belonging to a specific ‘trajectory’ and further assigned them to a sub-stage within that trajectory. Individuals who were assigned to the later stages of the ‘trajectory’ showed significant correlation with less GMV of Broca’s area (**Fig.1e**, *r*=0.651, *p*<0.0001) and hippocampus (**Fig.1f**, *r*=0.615, *p*<0.0001). In addition, the later stages were correlated with longer disease duration (**Fig.1g**, *r*=0.105, *p*<0.0001), worse negative symptoms (**Fig.1h**, *r*=0.101, *p*<0.0001) and worse cognitive symptoms (**Fig.1i**, *r*=0.080, *p*=0.004). These results suggest that the SuStaIn ‘trajectory’ reflects the underlying neural progression in schizophrenia.

### 2.5 Subtype-specific signatures in neuroanatomical pathology

To characterize subtype-specific neuroanatomical signatures, we assessed regional morphological measures using FreeSurfer in a subsample including 1,840 individuals with schizophrenia and 1,780 healthy controls. A total of 330 regional morphological measures in cortical thickness, cortical surface area, cortical volume, subcortical volume and subregion segmentation were quantified (see **Methods**).

Regional morphological z-scores (i.e., normative deviations from healthy control group) for each subtype were computed and compared (**Fig.3)**. Morphological z-scores of all brain regions and inter-subtype comparisons are provided in the **Supplementary Table S5**. Briefly, compared to healthy controls, average cortical volume/area reduction was only observed in subtype 1 (**Extended Data Fig.2a-b**), though both subtype 1 and subtype 2 exhibited a moderate reduction in average cortical thickness (**Extended Data Fig.2c**). Additionally, largest effects for cortical thickness/volume/area were located within the superior frontal regions for subtype 1 and in the superior/medial temporal regions for subtype2 (**Supplementary Table S5**). As for subcortical volume, larger effects for volumes of hippocampus, amygdala, thalamus, accumbens and brain stem were observed in subtype 2 compared to subtype 1 (**Extended Data Fig.2d-h**). The hippocampal/amygdala subregions with the most significant reduction for subtype 2 were located in the molecular layer and cortico-amygdaloid transition area (**Extended Data Fig.3-4**). Interestingly, we observed that, compared to healthy controls, the striatum (i.e., caudate, putamen) was larger among subtype 1 patients and smaller among subtype 2 patients (**Extended Data Fig.2i-j**). The difference in the striatum between the two subtypes was also replicated in a subsample of medication-naive individuals with schizophrenia (**Supplementary Table S6**). The main findings of subtype-specific neuroanatomical signatures are described in **Table** Taken together, subtype 1 exhibited greater deficits in cortical morphology but enlargement volume of striatum, whereas subtype 2 displayed more severe volume loss in the subcortical regions including hippocampus, amygdala, thalamus, brain stem as well as the striatum.

**Figure 3.**
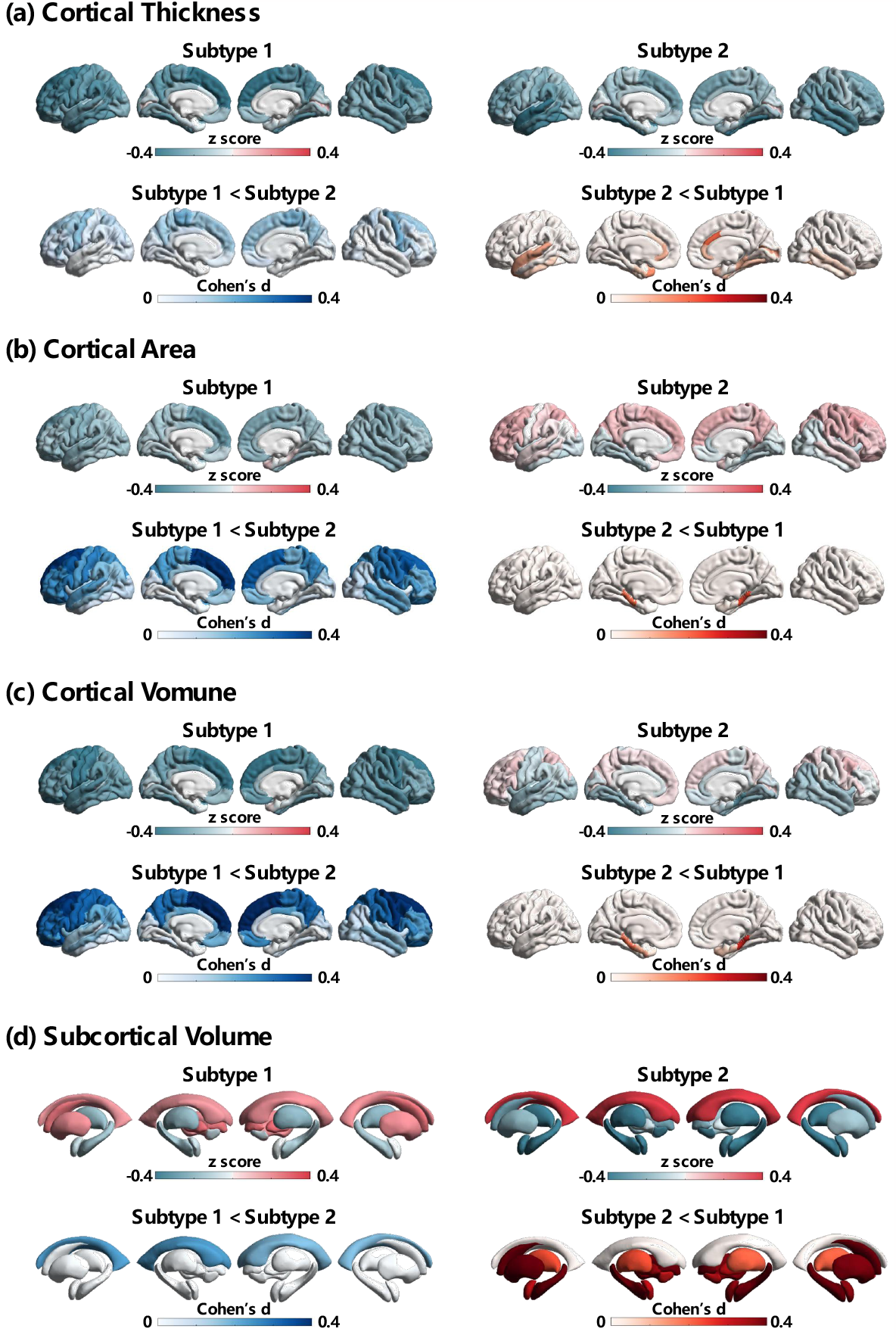
Subtype-specific signatures in neuroanatomical pathology. Regional Morphological z-scores (i.e., normative deviations from healthy control group) for each subtype are mapped to a brain template for visualization. Effect size of inter-subtype difference is quantified using Cohen’s d.

### 2.5 Clinical characterization of subtypes

A total of 2,622 (62.1%) individuals with schizophrenia were assigned to subtype 1 and the remaining 1,600 patients (37.9%) were assigned to subtype 2. The two subtypes exhibit no significant difference in the age, sex, illness duration or PANSS scores (**Table 1**). To further characterize the psychotic symptomatic trajectory as disease progresses for each subtype, we further defined three subgroups according to illness duration (early stage: <2 years; middle stage: 2-10 years; late stage: >10 years). The results suggested distinct trajectories of psychotic symptoms between the two subtypes (**Fig.4** and **Table 3**). Specifically, lower positive symptom severity was observed in late stage patients compared early stage patients in both subtypes (subtype 1, F=37.4, p=1.60e-16; subtype2, F=41.9, p=4.68e-18); however, at the late stage, subtype 1 exhibited worse positive symptom compared to subtype 2 (t=2.8, p=0.005). With the increase of the disease course, subtype 1 showed a gradual worsening of negative symptom (F=4.6, p=9.98e-3), whereas the negative symptoms of subtype 2 remained stable across the three stages of the disease course (F=0.1, p=0.884). Additionally, a gradual worsening of depression/anxiety was only observed in subtype 1 (F=5.9, p=2.86e-3), which showed worse depression/anxiety at the late stage, compared to subtype 2 (t=2.1, p=0.036).

**Figure 4.**
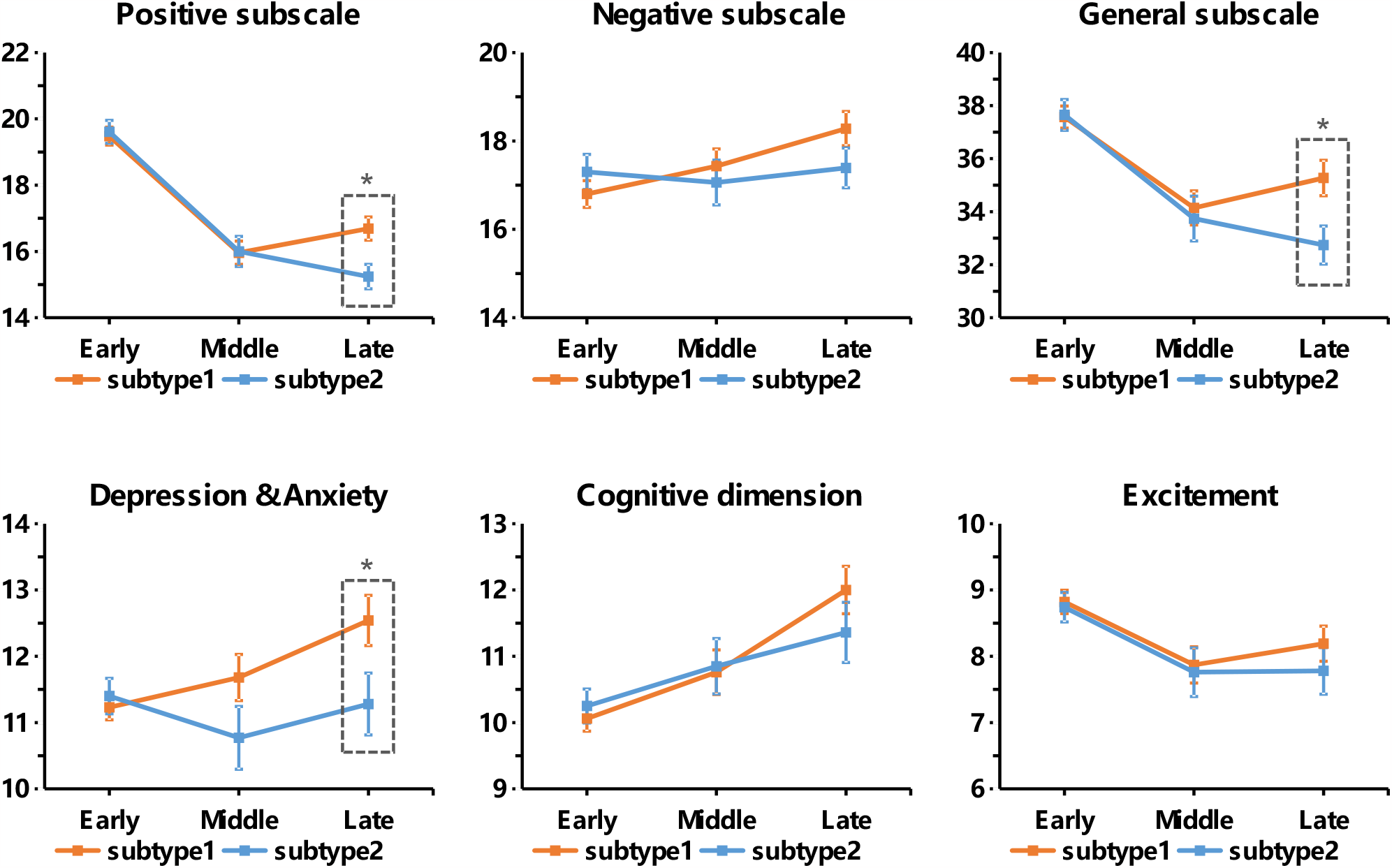
Symptomatic trajectories across three stages of disease duration. Individuals of each subtype are divided into three subgroups according to their illness durations (early stage: ≤2 years; middle stage: 2-10 years; late stage: >10 years). Data are presented as mean values +/-se. * *p*<0.05.

## 3 Discussion

Our study, applying a machine learning algorithm to brain MRI data from over 4,000 individuals with schizophrenia, has revealed two distinct neurostructural subtypes based on patterns of neuro-pathological progression. These subtypes are reproducible and generalizable across different subsamples and illness stages, independent of macroeconomic and ethnic factors that differed across collection locations. Specific patterns of neuroanatomical pathology for each subtype were uncovered. Subtype 1 is characterized by early cortical-predominant loss that first occurs in the Broca’s area/fronto-insular cortex, and shows adverse signatures in cortical morphology and an enlarged striatum. In contrast, subtype 2 is marked by early subcortical-predominant loss that first appears in the hippocampus, and displays significant volume loss in subcortical regions, including the hippocampus, amygdala, thalamus, brain stem and striatum. Additionally, we observed distinct trajectories of specific symptoms clusters in these two subtypes: as disease progresses, subtype 1 exhibited a gradual worsening of negative and depression/anxiety symptoms, and less of a decline in positive symptoms compared to subtype 2.

Despite the growing body of evidence pointing to group-level gray matter volume deficits in various brain regions - especially in frontal and temporal regions - as well as altered subcortical volume in schizophrenia [27], substantial individual variations persist within this population [11, 28]. These inter-individual differences in brain structure may stem from two primary sources of variation. First, differences in underlying etiology and pathogenesis could result in varying clinical characteristics (referred to as phenotypic heterogeneity) [6, 29]. Second, relative differences among subjects in the stage of dynamic progression (known as temporal heterogeneity) could further increase differences in the clinical presentation [30, 31]. Such variations suggest that the pathological progression of schizophrenia might not be attributed to a single unified pathophysiological process. Indeed, our neurostructural subtypes uncovered two patterns of gray matter loss trajectory through brain structural imaging. Several studies also reported dynamic patterns of accelerated gray matter loss over time in individuals with schizophrenia [32, 33]. In addition, staging of trajectory within subtype reflects the underlying neurophysiological, pathological and neuropsychological progressions in schizophrenia. Furthermore, we demonstrated that the phenotypic difference in the intrinsic neuro-pathophysiological trajectory was reproducible across samples worldwide, independent of macroeconomic and ethnic factors that differed across these sites.

The Broca’s area/fronto-insular cortex and hippocampus are identified separately in subtype 1 and subtype 2 as the first regions to show gray matter deficits. This is consistent with our prior finding based on individuals with schizophrenia primarily collected from the Chinese population [22]. Furthermore, the current study replicates the same two primary regions in a medication-naÏve and a first-episode cohort, suggesting that these neuropathological changes are a reflection of the disease process, rather than medication effects. Broca’s area and the fronto-insular cortex have been extensively implicated in schizophrenia [34], supporting Crow’s linguistic primacy hypothesis [35] and a triple-network model of the disorder [36]. Moreover, in individuals with psychosis, reductions in the inferior frontal cortex preceding the initial psychotic episode have been reported [37, 38]. A prior study reported reduced dopamine release in the prefrontal cortex in patients with schizophrenia [39]. In relation to hippocampal pathology, research has emphasized the hippocampus as one of the initial regions to display volumetric loss in schizophrenia [40, 41]. The hippocampus is thought to be involved in potential glutamatergic dysfunction in schizophrenia [6]. Decreased levels of the NMDA co-agonist D-serine were linked to neurobiological alterations similar to those seen in schizophrenia, including hippocampal volume loss [42]. These findings offer evidence regarding the specific neuroanatomical locations where gray matter loss is first observed in the schizophrenia subtypes. These two potential origins could also offer a new viewpoint on the pathological ‘spread’ of the disorder.

The new subtyping method employed exhibits high potential for distinguishing neurostructural subtypes with shared pathophysiological foundations. Notably, subtype 1 displayed larger volume of the striatum, while subtype 2 demonstrated reduced volume. The striatum plays a key role in the dopamine system, which contributes to psychotic symptoms [43]. Nevertheless, studies of striatal pathology have reported inconsistent differences between patients and controls [6]. The variability of the striatum is greater in patients than in controls, which relates to overall structural morphometry [27], dopamine D2 receptor and transporter levels [44]. This indicates that differences might exist within subgroups of the disorder [6]. In addition, it is still uncertain whether the discrepancy in striatum between cases and controls indicates a primary pathology or an effect of antipsychotic treatment [6]. Interestingly, this study’s subtype-specific striatal differences were replicated in a subset of individuals who had not received antipsychotic treatment, suggesting that striatal variability persists even in those without antipsychotic treatment. In addition, a recent study reveals a more pronounced and widespread pattern of thinner cortex in deficit schizophrenia, a clinically defined subtype with primary, enduring negative symptoms, compared to non-deficit schizophrenia [45]. This also suggests the existence of distinct subtypes distinguished by unique neuroimaging features. Taken together, our neurostructural subtyping differentiated subgroups with unique pathological features, thereby enhancing our understanding of the neurobiological mechanisms underlying schizophrenia.

The two newly identified subtypes may have several potential therapeutic implications. While the underlying mechanisms associated with a subtype-specific symptomatic trajectory remain unclear, our research shows divergent long-term clinical outcomes between the two neurostructural subtypes. As the disease advanced, for subtype 1, the negative and depression/anxiety symptoms gradually worsened; for subtype 2 these symptoms remained stable. In addition, subtype1 experienced worse positive symptoms than subtype 2 at the late stage of disease (i.e., duration > 10 years). This is consistent with a prior study that reported greater gray matter reduction in frontal regions in treatment-resistant compared with treatment-responsive individuals with schizophrenia [46]. Another intriguing aspect is that our prior research on treatment-resistant schizophrenia demonstrated that electroconvulsive therapy (ECT) can substantially enhance the volume of the hippocampus and insula; this is also associated with psychotic symptom alleviation [47-49]. Notably, these two brain regions were also identified as the ‘origins’ of gray matter loss separately in each subtype. This observation raises the possibility of exploring neuromodulation interventions, such as transcranial magnetic stimulation (TMS), to target these specific brain regions.

This study has several limitations. First, while the SuStaIn algorithm estimates pathophysiological trajectories from cross-sectional MRI data, it remains crucial to validate these outcomes with longitudinal data to verify the brain changes with disease progression over time. Second, the current study benefits from a large sample size, but the inclusion of data from various sites could potentially be influenced by confounding factors, including diverse cohorts, scanners, and locations. Harmonization methods have been employed to alleviate disparities across MRI acquisition protocols. Nonetheless, it remains essential to collect a sufficiently large sample from multi-centers under a standard imaging protocol and experimental paradigm. Third, a substantial portion of individuals with schizophrenia were likely to have received or currently use medications, and data from medication-naÏve/free individuals were only available for a subset of the datasets. One important limitation is the assumption of progressive pathology in schizophrenia (discrete events of tissue loss or continuous downward drift), when applying SuStaIn. The few existing very long-term imaging studies in schizophrenia support this stance [50] but selection bias cannot be fully overcome in the recruitment process for neuroimaging studies. Routine anatomical MRI for every person with psychosis seeking help, with periodic repeats, may provide better view of the validity of progressive pathology in the future.

In summary, our study reveals two distinct neurostructural schizophrenia subtypes based on patterns of pathological progression of gray matter loss. We extend the reproducibility and generalisability of these brain imaging-based subtypes across illness stages, medication treatments and different sample locations worldwide, independent of macroeconomic and ethnic factors that differed across these sites. The identified subtypes exhibit distinct signatures of neuroanatomical pathology and psychotic symptomatic trajectories, highlighting the heterogeneity of the neurobiological changes associated with disease progress. This new imaging-based taxonomy shows potential for identification of homogeneous subsamples of individuals with shared neurobiological characteristics. This may be a first crucial step in the transition from only syndrome-based to both syndrome- and biology-based identification of mental disorder subtypes in the near future.

## 4 Methods

### 4.1 Study samples

This study analyzed cross-sectional T1-weighted structural MRI data from a total of 4,291 individuals diagnosed with schizophrenia (1,709 females, mean age=32.5±11.9 years) and 7,078 healthy controls (3,461 females, mean age=33.0±12.7 years). These datasets came from 21 cohorts of ENIGMA schizophrenia working groups from various countries around the world, 11 cohorts collected from Chinese hospitals over the last ∼10 years, and 9 cohorts from publicly available datasets, i.e., HCP-EP [51], JP-SRPBS [52], fBIRN [53], MCIC [54], NMorphCH [55], NUSDAST [56], DS000030 [57], DS000115 [58] and DS004302 [59]. The datasets came from various countries around the world (**Extended Data Fig.5**). Details of demographics, geographic location, clinical characteristics, and inclusion/exclusion criteria for each cohort may be found in the **Supplementary Information** (**Supplementary Table S1-2**).

The severity of symptoms was evaluated by the Positive and Negative Syndrome Scale (PANSS) [60], including a positive scale (total score of P1-P7), a negative scale (total score of N1-N7), a general psychopathology scale (total score of G1-G16) and total score. In addition, phenotypic characteristics were further quantified in three dimensions, such as cognitive (total score of P2, N5, G5, G10, G11), depression/anxiety (total score of G1, G2, G3, G6, G15) and excitement (total score of P4, P7, G44, G14) via a five-factor model of schizophrenia [61].

All sites obtained approval from their local institutional review boards or ethics committees, and written informed consent from all participants and/or their legal guardians. The present study was carried out under the approve from the Medical Research Ethics Committees of Fudan University (Number: FE222711).

### 4.2 Image acquisition, processing and quality control

T1-weighted structural brain MRI scans were acquired at each study site. We used a standardized protocol for image processing using the ENIGMA Computational Anatomy Toolbox (CAT12) across multiple cohorts (https://neuro-jena.github.io/enigma-cat12/). These protocols enable region-based gray matter volume (GMV) measures for image data based on the automated anatomical (AAL3) atlas [62]. Further details of image acquisition parameters and quality control may be found in **Supplementary Information** (**Supplementary Table S1-2**).

### 4.3 Data harmonization

The ROI-wise GMV measures were first adjusted by regressing out the effects of sex, age, the square of age, site and total intracranial volume (TIV) using a regression model [22]. Subsequently, a harmonization procedure was performed using the ComBat algorithm for correcting multi-site data [63]. The adjusted values were transformed as z-scores (i.e., normative deviations) relative to the healthy control group. We multiplied these z-scores by -1 so that the z-score increases as regional GMV decreases. Finally, we removed these samples if they were marked as a statistical outlier (>5 standard deviations away from the global mean). After the quality control, 11,260 individuals were included, of which 4,222 were schizophrenia patients (1,683 females, mean age=32.4±12.4 years) and 7,038 healthy subjects (3,440 females, mean age=33.0±12.4 years).

### 4.4 Disease progress modelling

To uncover diverse patterns of pathophysiological progression from cross-sectional only MRI data and cluster individuals into groups (subtypes), we employed a novel machine learning approach - Subtype and Stage Inference (SuStaIn) [19]. The methodology of SuStaIn has been described in detail previously [19]. Here, we briefly describe the main parameter choices specific to the current study. The SuStaIn model requires an M x N matrix as input. M represents the number of cases (M=4,222). N is the number of biomarkers (N=17). 17 gray matter biomarkers that were previously used for SuStaIn modelling in schizophrenia [22]. Here, all of the AAL3 regions of whole brain were separated and merged into 17 regions of interest (ROIs) [22], including frontal lobe, temporal lobe, parietal lobe, occipital lobe, insula, cingulate, sensorimotor, Broca’s area, cerebellum, hippocampus, parahippocampus, amygdala, caudate, putamen, pallidum, accumbens and thalamus (**Supplementary Table S6**). We then ran the SuStaIn algorithm with 25 start points and 100,000 Markov Chain Monte Carlo (MCMC) iterations [19] to estimate the most likely sequence that describes spatiotemporal pattern of pathophysiological progression (i.e., ‘trajectory’).

SuStaIn can identify diverse trajectories of pathophysiological progression given a subtype number K. We fitted the model for K=2-6 subtypes (‘trajectories’), separately. The optimal number of subtypes was determined according to the reproducibility of individual subtyping via a two-fold cross-validation procedure, as described previously [22]. Specifically, all individuals were randomly split into two non-overlapping folds. For each fold, we trained the SuStaIn model. For each individual, the trained SuStaIn model provides a subtype label. We measured the consistency of the subtype labeling across all individuals between two folds by using the Dice coefficient. This above procedure was repeated ten times. The largest Dice coefficient was obtained for K=2 (see **Figure 1a)**, indicating the best consistency based on cross-validation. Finally, the two-cluster model of SuStaIn was fitted to the entire sample. The most probable sequence (i.e., the order of biomarkers) was evaluated for each ‘trajectory’ via SuStaIn. For each individual, SuStaIn calculated the probability (ranging from 0 to 1) of belonging to each ‘trajectory’, and assigned the individual into a sub-stage of the maximum likelihood ‘trajectory’ through MCMC iterations. We also estimated the SuStaIn ‘trajectories’ based on a subsample from individuals with first-episode schizophrenia whose illness duration was less than two years (N=1,122, 513 females, mean age=25.4±8.6 years), and a subsample of medication-naÏve individuals with schizophrenia (N=718, 353 females, mean age=23.7±7.8 years).

### 4.5 Visualization of pathophysiological progression trajectory

To visualize the spatiotemporal patterns of pathophysiological progression, we calculated the mean z-score of regional GMV across individuals belonging to the same substage of each SuStaIn ‘trajectory’. The images of ROI-wise GMV z-scores were mapped into a glass brain template via visualization tools implemented in ENIGMA Toolbox (https://enigma-toolbox.readthedocs.io/en/latest/index.html) and BrainNetViewer (https://www.nitrc.org/projects/bnv/).

To examine whether the SuStaIn stage (a continuous indicator of the ‘temporal’ stage of SuStaIn ‘trajectory’) is associated with pathological processes and clinical characteristics in schizophrenia, we performed Spearman correlations between the SuStaIn stages and the degree of brain atrophy (i.e., regional GMV) in schizophrenia. We also examined whether SuStaIn stages were linked to disease duration, severity of symptoms, and phenotypic characteristics.

### 4.6 Neuroanatomical signatures using regional morphological measures

To further characterize the neuroanatomical signatures associated with each subtype, we conducted regional morphological analyses in a subsample including 1,840 individuals with schizophrenia and 1,780 healthy controls. Brain morphological measures, such as cortical thickness, cortical surface area, cortical volume and subcortical volume, were quantified using FreeSurfer (version 7.3, http://surfer.nmr.mgh.harvard.edu/). A total of 68x3 regional measures for cortical thickness, cortical surface area and cortical volume were extracted based on the DK atlas [64], along with 14 subcortical regions (bilaterally nucleus accumbens, amygdala, caudate, hippocampus, pallidum, putamen and thalamus) and 2 lateral ventricles. In addition, we performed an automated subregion segmentation (https://surfer.nmr.mgh.harvard.edu/fswiki/SubregionSegmentation) for the hippocampal substructures (n=38 subregions) [65], the nuclei of the amygdala (n=18) [66], the thalamic nuclei (n=50) [67], and the brain stem structures (n=4) [68], yielding a total of 110 subregional volumetric measures.

Regional morphological measures for each individual with schizophrenia were adjusted by regressing out the effects of sex, age, the square of age, TIV and site, and then transformed to z-scores (i.e., normative deviations from healthy control group). The mean regional morphological z-score across individuals belonging to each subtype was calculated, and mapped to brain templates for visualization of neuroanatomical signature deviation for each subtype relative to healthy population. To further manifest subtype-specific signature in neuroanatomical pathology, we compared the regional morphological z-scores between the two subtypes using two sample *t*-tests. Multiple comparisons were corrected by family wise error (FWE) correction.

### 4.7 Distinct symptom profiles between subtypes

To characterize the psychotic symptomatic trajectory with disease duration increases for each subtype, we further divided the individuals of each subtype into three subgroups according to their illness durations (early stage: <2 years; middle stage: 2-10 years; late stage: >10 years). We compared the difference of symptoms among the three stages of disease in each subtype using ANOVA. Two sample *t*-tests were performed to compare the inter-subtype differences separately between each of the stages.

## Data availability

Data of NMorphCH, FBIRN and NUSDAST were obtained from the SchizConnect, a publicly available website (http://www.schizconnect.org/documentation#by_project). The NMorphCH dataset and NUSDAST dataset were download through a query interface at the SchizConnect (http://www.schizconnect.org/queries/new). The FBIRN dataset was download from https://www.nitrc.org/projects/fbirn/. The DS000115 dataset was download from OpenfMRI database (https://www.openfmri.org/). The DS000030 dataset was available at https://legacy.openfmri.org/dataset/ds000030/. The DS004302 dataset was available at https://openneuro.org/datasets/ds004302/versions/1.0.1. The HCP-EP dataset was available at https://www.humanconnectome.org/study/human-connectome-project-for-early-psychosis/. The Japanese SRPBS Multi-disorder MRI Dataset was available at https://bicr-resource.atr.jp/srpbsopen/. Requests for ENIGMA data can be applied via the ENIGMA Schizophrenia Working Group (https://enigma.ini.usc.edu/ongoing/enigma-schizophrenia-working-group/). Requests for raw and analyzed data can be made to the corresponding author (J.Feng) and will be promptly reviewed by the Fudan University Ethics Committee to verify whether the request is subject to any intellectual property or confidentiality obligations.

## Code availability

SuStaIn algorithm is available on the UCL-POND GitHub (https://github.com/ucl-pond/). T1-weighted images were processed using the Computational Anatomy Toolbox for Standardized Processing of ENIGMA Data (https://neuro-jena.github.io/enigma-cat12/). A protocol for the current data processing is available at https://docs.google.com/document/d/1lb9v0v4j_OrgAKDh6_9fl3Hz2Wcfg46c/edit/.

FreeSurfer (version 7.3, http://surfer.nmr.mgh.harvard.edu/) was used to quantify various morphological measures, such as cortical thickness, cortical surface area, cortical volume and subcortical volume. The visualization of ROI-wise z-score images was conducted using BrainNetViewer (https://www.nitrc.org/projects/bnv/).

## Acknowledgements

This work was supported by the grant from Science and Technology Innovation 2030-Brain Science and Brain-Inspired Intelligence Project (No. 2022ZD0212800). This work was supported by National Natural Science Foundation of China (No. 82202242, 82071997). This work was supported by the projects from China Postdoctoral Science Foundation (No. BX2021078, 2021M700852). This work was supported by the Shanghai Rising-Star Program (No. 21QA1408700) and the Shanghai Sailing Program (22YF1402800) from Shanghai Science and Technology Committee. This work was supported by National Key R&D Program of China (No. 2019YFA0709502), the grant from Shanghai Municipal Science and Technology Major Project (No. 2018SHZDZX01), ZJ Lab, and Shanghai Center for Brain Science and Brain-Inspired Technology, and the grant from the 111 Project (No. B18015). ASRB: Supported by NHMRC Project Grants (630471 & 1081603). ESO: Supported by Ministry of Health of the Czech Republic, grants nr. NU21-08-00432 and NU20-04-00393. Supported by Ministry of Health, Czech Republic - DRO 2021 (“Institute for Clinical and Experimental Medicine - IKEM, IN: 00023001”). FOR2107: Funded by the German Research Foundation (Deutsche Forschungsgemeinschaft DFG; Forschungsgruppe/Research Unit FOR2107). Principal investigators (PIs) with respective areas of responsibility in the FOR2107 consortium are: Work Package WP1, FOR2107/MACS cohort and brainimaging: Tilo Kircher (speaker FOR2107; DFG grant numbers KI588/14-1, KI588/14-2, KI588/20-1, KI588/22-1), Udo Dannlowski (co-speaker FOR2107; DA 1151/5-1, DA 1151/5-2, DA1151/6-1), Axel Krug (KR 3822/5-1, KR 3822/7-2), Igor Nenadic (NE2254/1-2, NE2254/3-1, NE2254/4-1, NE2254/2-1), Carsten Konrad (KO 4291/3-1). IMH: Supported by research grants from the National Healthcare Group, Singapore (SIG/05004; SIG/05028), and the Singapore Bioimaging Consortium (RP C009/2006) research grants awarded to Kang Sim. JBUN: Supported by Korean Mental Health Technology R&D Project, Ministry of Health & Welfare, Republic of Korea (HL19C0015) and a grant of the Korea Health Technology R&D Project through the Korea Health Industry Development Institute (KHIDI), funded by the Ministry of Health & Welfare, Republic of Korea (HR18C0016). OLIN: Supported by MH106324. Osaka: Supported by JSPS KAKENHI (Grant Number JP21K12153), ABiS (JSPS KAKENHI Grant Number JP22H04926), Brain/MINDS & beyond studies (Grant Number JP18dm0307002) from the AMED. PENS: Supported by department of Veterans Affairs CSR&D (I01CX000227) and National Institutes of Health (U01MH108150) to Scott R. Sponheim. RomeSL: Supported by Italian Ministry of Health, grant Ricerca Corrente RC 23, RF-2019-12370182. SoCAT: Supported by THE SCIENTIFIC AND TECHNOLOGICAL RESEARCH COUNCIL OF TÜRKIYE. SWIFT: Supported by a NARSAD grant from the Brain & Behavior Research Foundation (28445) and by a Research Grant from the Novartis Foundation (20A058). UCISZ: Supported by the National Institute of Mental Health of the National Institutes of Health under award number R21MH097196. UNINA: Supported by #NEXTGENERATIONEU (NGEU) and funded by the Ministry of University and Research (MUR), National Recovery and Resilience Plan (NRRP), project MNESYS (PE0000006) – A Multiscale integrated approach to the study of the nervous system in health and disease (DN. 1553 11.10.2022). COBRE: Supported by NIMH: 5R01MH094524-1; 3R01MH121246; 1P20RR021938. TOPSY: Supported by the Canada First Research Excellence Fund, awarded to the Healthy Brains, Healthy Lives initiative at McGill University (through a New Investigator Supplement) and the Monique H. Bourgeois Chair in Developmental Disorders. He receives a salary award from the Fonds de recherche du Québec - Santé (FRQS). OSLO: Supported by The Research Council of Norway (249795, 300767), the South-Eastern Norway Regional Health Authority (2014097, 2019101), KG Jebsen Stiftelsen, the European Research Council under the European Union s Horizon 2020 research and Innovation program (802998), and the European Union-funded Horizon Europe project ‘environMENTAL’ (no. 101057429). HCP-EP: Research using Human Connectome Project for Early Psychosis (HCP-EP) data reported in this publication was supported by the National Institute of Mental Health of the National Institutes of Health under Award Number U01MH109977. JP-SRPBS: Data used in the preparation of this work were obtained from the DecNef Project Brain Data Repository (https://bicr-resource.atr.jp/srpbsopen/) gathered by a consortium as part of the Japanese Strategic Research Program for the Promotion of Brain Science (SRPBS) supported by the Japanese Advanced Research and Development Programs for Medical Innovation (AMED). FBIRN: Data used for this study were downloaded from the FunctionBIRN Data Repository, supported by grants to the Function BIRN (U24-RR021992) Testbed funded by theNational Center for Research Resources at the National Institutes of Health, U.S.A. MCIC: The imaging data and demographic information was collected and shared by [University of Iowa, University of Minnesota, University of New Mexico, Massachusetts General Hospital] the Mind Research Network supported by the Department of Energy under Award Number DE-FG02-08ER64581. NMorphCH: Data collection and sharing was funded by NIMH grant R01 MH056584. NUSDAST: Data collection and sharing was funded by NIMH grant 1R01 MH084803. This work is supported by the Zhangjiang International Brain Biobank (ZIB) Consortium.

## Competing Interests Statement

Lena Palaniyappan reports personal fees from Janssen Canada, Otsuka Canada, SPMM Course Limited UK and the Canadian Psychiatric Association; book royalties from Oxford University Press; and investigator-initiated educational grants from Sunovion, Janssen Canada and Otsuka Canada, outside the submitted work. Tilo Kircher received unrestricted educational grants from Servier, Janssen, Recordati, Aristo, Otsuka, neuraxpharm. Philipp Homan has received grants and honoraria from Novartis, Lundbeck, Mepha, Janssen, Boehringer Ingelheim, Neurolite outside of this work. Ole A. Andreassen is a consultant to Cortechs.ai and received speakers honorarium from Lundbeck, Janssen, Sunovion. These interests played no role in the research reported here. Other authors disclose no conflict of interest.

**Table 1.**
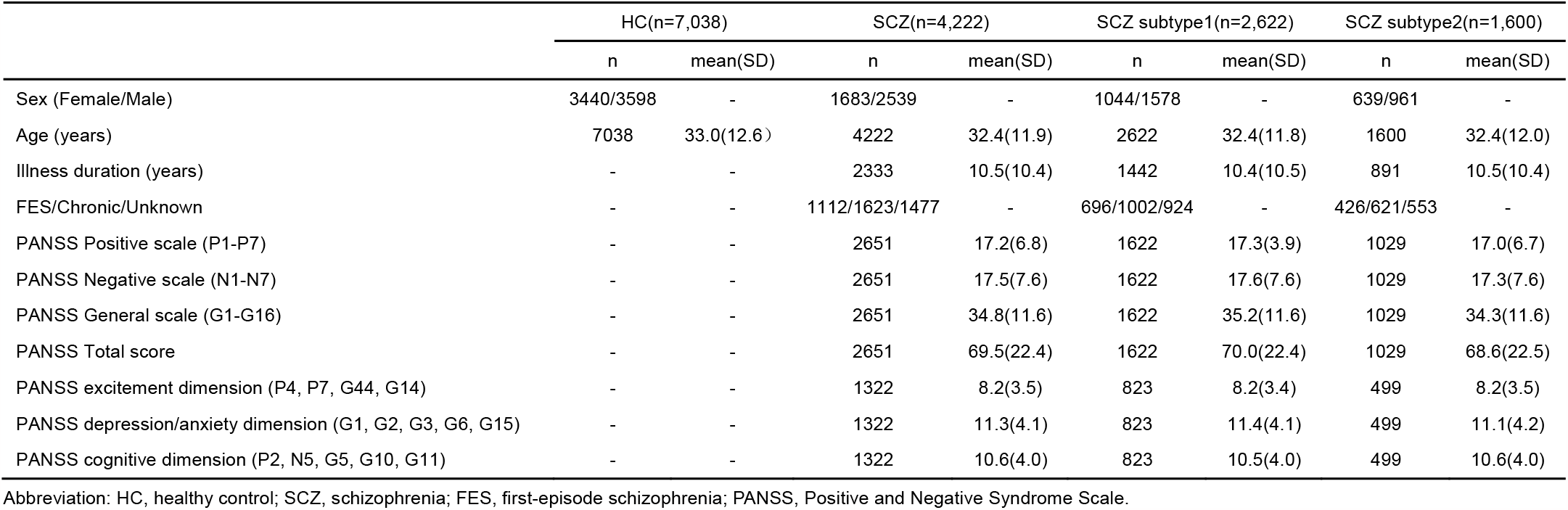
Demographic and clinical characteristics in the primary sample including 4,222 schizophrenia patients and 7,038 healthy controls.

|  | HC(n=7,038) |  | SCZ(n=4,222) |  | SCZ subtype1(n=2,622) |  | SCZ subtype2(n=1,600) |  |
| --- | --- | --- | --- | --- | --- | --- | --- | --- |
|  | n | mean(SD) | n | mean(SD) | n | mean(SD) | n | mean(SD) |
| Sex (Female/Male) | 3440/3598 | - | 1683/2539 | - | 1044/1578 | - | 639/961 | - |
| Age (years) | 7038 | 33.0(12.6) | 4222 | 32.4(11.9) | 2622 | 32.4(11.8) | 1600 | 32.4(12.0) |
| Illness duration (years) | - | - | 2333 | 10.5(10.4) | 1442 | 10.4(10.5) | 891 | 10.5(10.4) |
| FES/Chronic/Unknown | - | - | 1112/1623/1477 | - | 696/1002/924 | - | 426/621/553 | - |
| PANSS Positive scale (P1-P7) | - | - | 2651 | 17.2(6.8) | 1622 | 17.3(3.9) | 1029 | 17.0(6.7) |
| PANSS Negative scale (N1-N7) | - | - | 2651 | 17.5(7.6) | 1622 | 17.6(7.6) | 1029 | 17.3(7.6) |
| PANSS General scale (G1-G16) | - | - | 2651 | 34.8(11.6) | 1622 | 35.2(11.6) | 1029 | 34.3(11.6) |
| PANSS Total score | - | - | 2651 | 69.5(22.4) | 1622 | 70.0(22.4) | 1029 | 68.6(22.5) |
| PANSS excitement dimension (P4, P7, G44, G14) | - | - | 1322 | 8.2(3.5) | 823 | 8.2(3.4) | 499 | 8.2(3.5) |
| PANSS depression/anxiety dimension (G1, G2, G3, G6, G15) | - | - | 1322 | 11.3(4.1) | 823 | 11.4(4.1) | 499 | 11.1(4.2) |
| PANSS cognitive dimension (P2, N5, G5, G10, G11) | - | - | 1322 | 10.6(4.0) | 823 | 10.5(4.0) | 499 | 10.6(4.0) |

Abbreviation: HC, healthy control; SCZ, schizophrenia; FES, first-episode schizophrenia; PANSS, Positive and Negative Syndrome Scale.

**Table 2.** Main findings of subtype-specific neuroanatomical signatures.

| Morphometry measures | Subtype-specific neuroanatomical signatures |
| --- | --- |
| Cortical Thickness/Volume/Area | a) Both subtype1 and subtype2 exhibit a moderate degree in the average cortical thickness reduction.<br>b) Reduction of average cortical volume/area is only observed in the subtype1.<br>c) The worst reduction of cortical thickness/volume/area is located within the superior frontal regions for the subtype1, but in the superior/medial temporal regions for the subtype2. |
| Subcortical Volume | a) Enlargement of lateral ventricle is found in both subtype1 and subtype2, but much larger in the subtype2.<br>b) Worse loss volumes of hippocampus, amygdala, thalamus and accumbens are observed in the subtype2, compared to the subtype1.<br>c) Volumes of striatum (i.e., caudate, putamen) are increased in the subtype1, but decreased in the subtype2, compared to the healthy population. |
| Hippocampus segmentation | a) Volume loss in hippocampal subregions is worse in the subtype2, compared to the subtype1.<br>b) The most significant volume loss is in the molecular layer for the subtype2. |
| Amygdala segmentation | a) The subtype2 shows worse volume loss in amygdala subregions, compared to the subtype1.<br>b) The most significant decrease in volume is in the cortico-amygdaloid transition area for both the subtypes. |
| Thalamus segmentation | a) The subtype2 shows worse volume loss in thalamus subregions, compared to the subtype1. |
| Brain stem segmentation | a) Volume loss of brain stem subregions is only observed in the subtype2. |

**Table 3.** Symptom scores for each subtype at different stages of disease duration.

| Symptoms | Subtype 1 |  |  | F test |  | Subtype 2 |  |  | F test |  |
| --- | --- | --- | --- | --- | --- | --- | --- | --- | --- | --- |
|  | Early<br>(n=579) | Middle<br>(n=362) | Late<br>(n=400) | F | P | Early<br>(n=371) | Middle<br>(n=216) | Late<br>(n=282) | F | P |
| PANSS Positive scale (P1-P7) | 19.5(6.4) | 16.0(6.7) | 16.7(7.0)* | 37.4 | 1.60E-16 | 19.6(6.4) | 16.0(6.7) | 15.2(6.2)* | 41.9 | 4.68E-18 |
| PANSS Negative scale (N1-N7) | 16.8(7.3) | 17.4(7.4) | 18.3(7.7) | 4.6 | 9.98E-03 | 17.3(7.4) | 17.1(7.5) | 17.4(7.5) | 0.1 | 0.884 |
| PANSS General scale (G1-G16) | 37.6(10.0) | 34.1(12.2) | 35.3(13.3)* | 10.6 | 2.80E-05 | 37.7(10.8) | 33.7(12.4) | 32.7(12.0)* | 15.6 | 2.30E-07 |
| PANSS Total score | 73.9(19.7) | 67.5(23.2) | 70.2(25.0)* | 9.3 | 9.40E-05 | 74.5(20.9) | 66.7(23.4) | 65.4(22.4)* | 15.7 | 2.05E-07 |
| PANSS excitement dimension (P4, P7, G44, G14) | 8.8(3.4) | 7.9(3.3) | 8.2(3.4) | 4.9 | 8.01E-03 | 8.7(3.3) | 7.8(3.4) | 7.8(3.7) | 4.12 | 0.017 |
| PANSS depression/anxiety dimension (G1, G2, G3, G6, G15) | 11.2(3.7) | 11.7(4.2) | 12.5(4.9)* | 5.9 | 2.86E-03 | 11.4(4.0) | 10.8(4.4) | 11.3(4.9)* | 0.7 | 0.511 |
| PANSS cognitive dimension (P2, N5, G5, G10, G11) | 10.1(3.7) | 10.8(4.0) | 12.0(4.6) | 13.5 | 1.74E-06 | 10.3(3.8) | 10.9(3.9) | 11.4(4.7) | 2.8 | 0.061 |

\* indicates significant difference between the subtype1 and subtype2 using two sample t test (p<0.05).

## Figures Legend

**Extend Data Fig 1.**
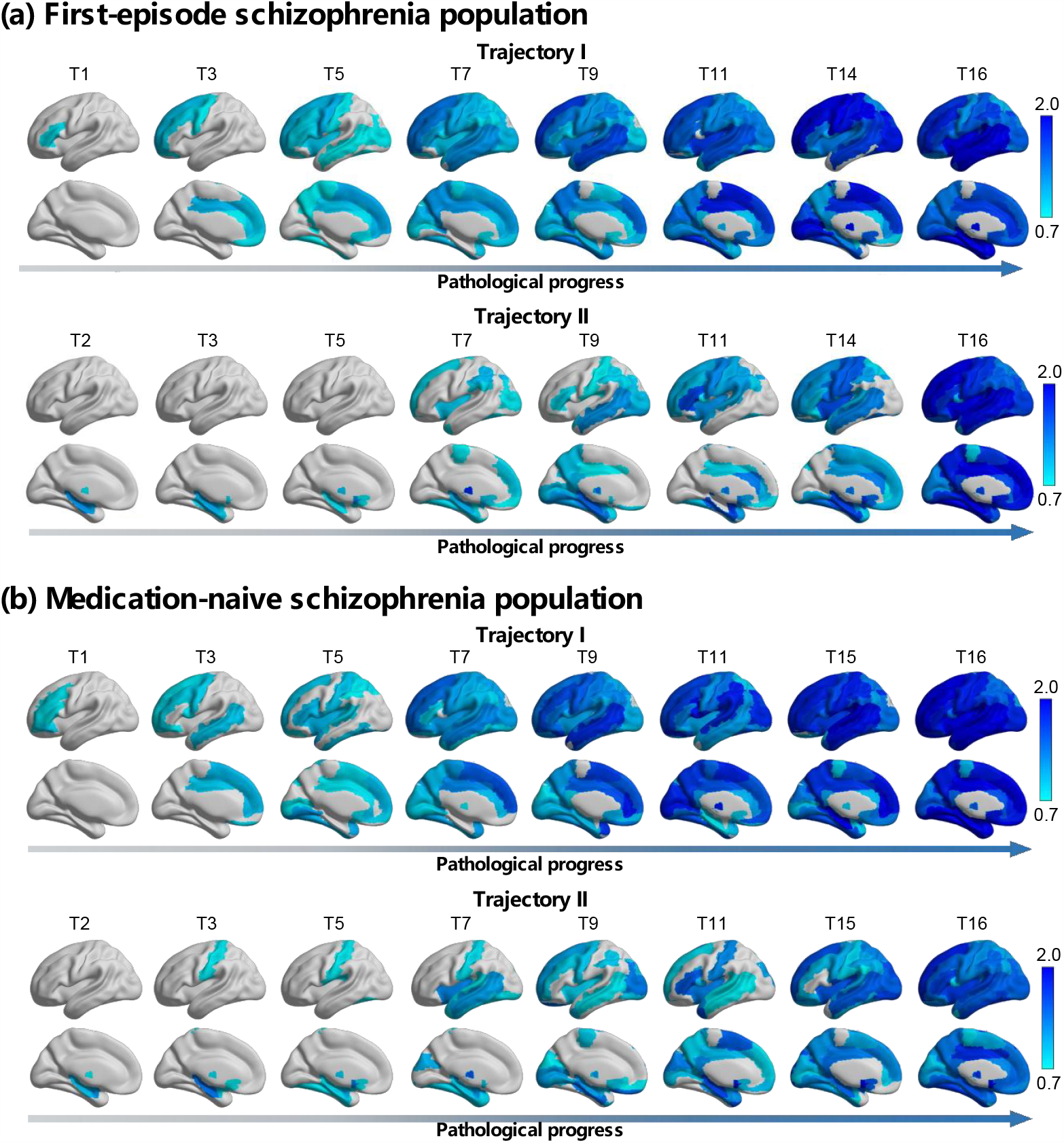
Pathophysiological progression trajectories in first-episode population and medication-naÏve population. Trajectories are repeated based on the subsample data from the first-episode schizophrenia patients whose illness duration was less than two years (N=1,112, 513 females, mean age=25.4±12.4 years), and another subsample data from medication-naÏve patients with schizophrenia (N=718, 353 females, mean age=23.7±12.1 years).

**Extend Data Fig 2.**
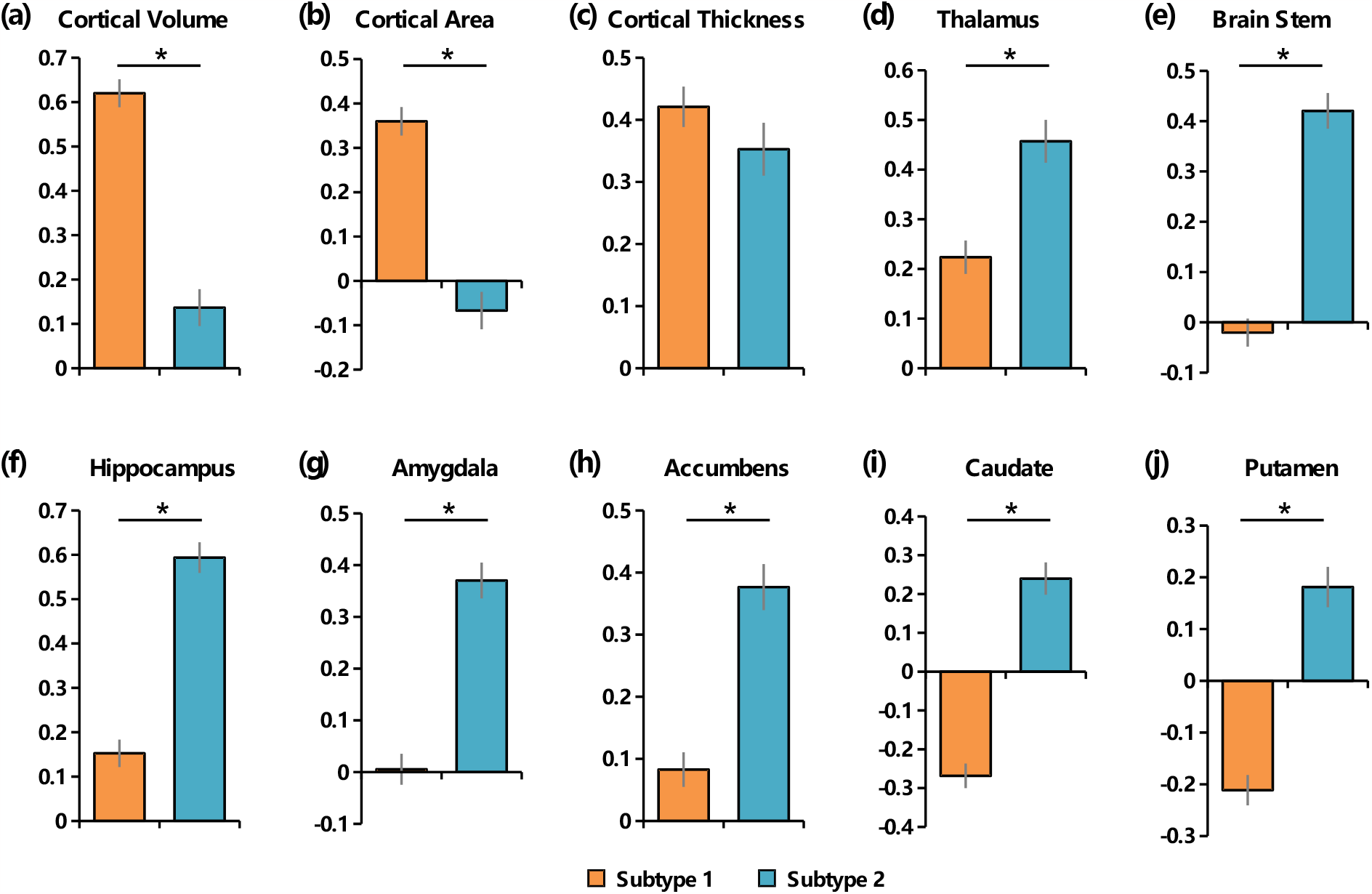
Comparisons of morphological z-score between the two subtypes. A larger positive z-score indicates a larger deviation of reduction relative to healthy control group. Two sample t test is conducted to examine inter-subtype difference for the (a) averaged cortical volume (t=9.36, p<10e-16, Cohen’s d=0.446); (b) averaged cortical area (t=8.09, p<10e-16, Cohen’s d=0.386); (c) averaged cortical thickness (t=1.29, p=0.198, Cohen’s d=0.061); (d) thalamus volume (t=-4.28, p=1.97e-5, Cohen’s d=-0.205); (e) brain stem volume (t=-9.79, p<10e-16, Cohen’s d=-0.469); (f) hippocampus volume (t=-9.25, p<10e-16, Cohen’s d=-0.449); (g) amgydala volume (t=-7.83, p=8.44e-15, Cohen’s d=-0.379); (h) accumbens volume (t=-6.40, p=1.94e-10, Cohen’s d=-0.305); (i) caudate volume (t=-9.82, p<10e-16, Cohen’s d=-0.468); (j) putamen volume (t=-8.14, p<10e-16, Cohen’s d=-0.389).

**Extend Data Fig 3.**
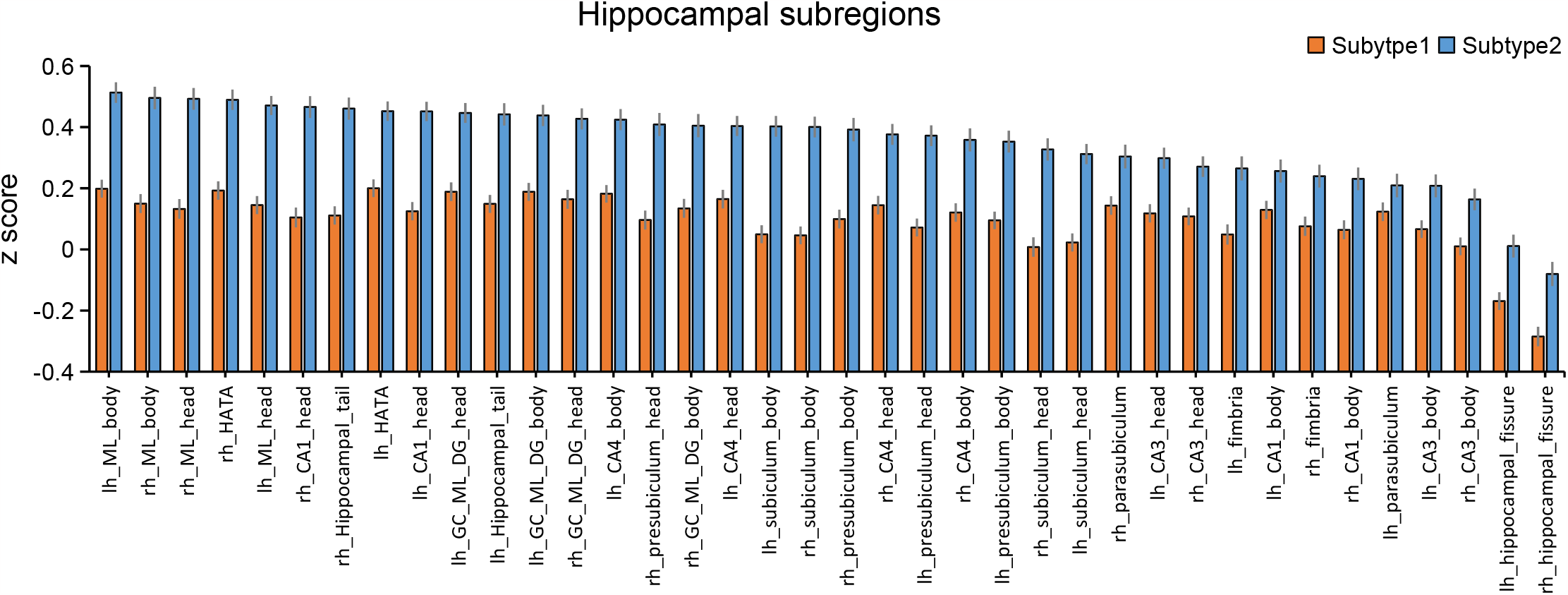
Hippocampus subregional morphological z-score for the two subtypes. A larger positive z-score indicates a larger deviation of reduction relative to healthy control group.

**Extend Data Fig 4.**
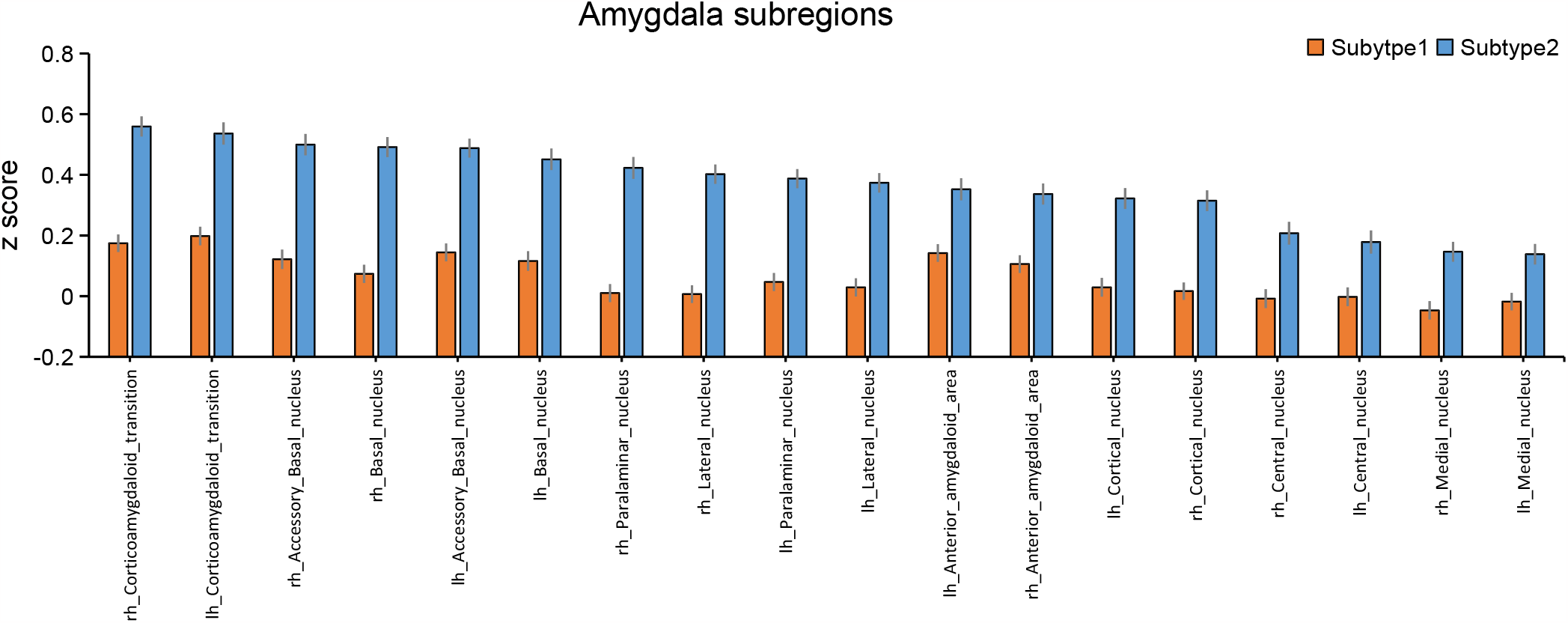
Amygdala subregional morphological z-score for the two subtypes. A larger positive z-score indicates a larger deviation of reduction relative to healthy control group.

**Extend Data Fig 5.**
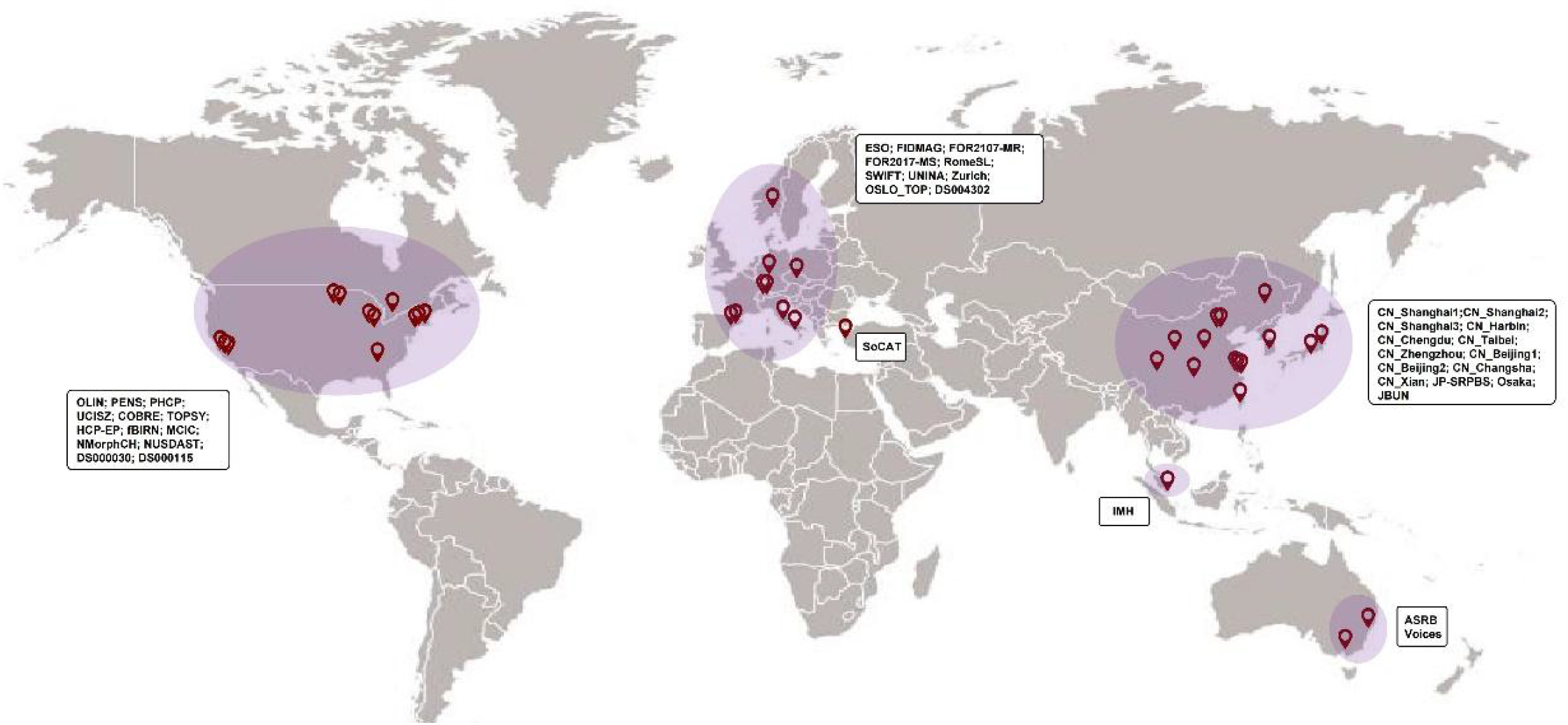
Geographic map of included datasets.

## Notes

### Competing Interest Statement

The authors have declared no competing interest.

### Author Declarations

The present study was carried out under the approve from the Medical Research Ethics Committees of Fudan University (Number: FE222711).

## References

1. The, L., ICD-11: a brave attempt at classifying a new world. The Lancet, 2018. 391(10139): p. 2476.

2. Oren, O., B.J. Gersh, and D.L. Bhatt, Artificial intelligence in medical imaging: switching from radiographic pathological data to clinically meaningful endpoints. Lancet Digit Health, 2020. 2(9): p. e486–e488.

3. Rajpurkar, P., et al., AI in health and medicine. Nat Med, 2022. 28(1): p. 31–38.

4. Organization, W.H., The global burden of disease: 2004 update. 2008: World Health Organization.

5. Howes, O.D. and E.C. Onwordi, The synaptic hypothesis of schizophrenia version III: a master mechanism. Mol Psychiatry, 2023.

6. McCutcheon, R.A., J.H. Krystal, and O.D. Howes, Dopamine and glutamate in schizophrenia: biology, symptoms and treatment. World Psychiatry, 2020. 19(1): p. 15–33.

7. Wolfers, T., et al., Mapping the Heterogeneous Phenotype of Schizophrenia and Bipolar Disorder Using Normative Models. JAMA Psychiatry, 2018. 75(11): p. 1146–1155.

8. Fusar-Poli, P., et al., Heterogeneity of Psychosis Risk Within Individuals at Clinical High Risk: A Meta-analytical Stratification. JAMA Psychiatry, 2016. 73(2): p. 113–20.

9. McCutcheon, R.A., et al., The efficacy and heterogeneity of antipsychotic response in schizophrenia: A meta-analysis. Mol Psychiatry, 2021. 26(4): p. 1310–1320.

10. Collado-Torres, L., et al., Regional Heterogeneity in Gene Expression, Regulation, and Coherence in the Frontal Cortex and Hippocampus across Development and Schizophrenia. Neuron, 2019. 103(2): p. 203–216 e8.

11. Brugger, S.P. and O.D. Howes, Heterogeneity and Homogeneity of Regional Brain Structure in Schizophrenia: A Meta-analysis. JAMA Psychiatry, 2017. 74(11): p. 1104–1111.

12. Braff, D.L., et al., Lack of use in the literature from the last 20 years supports dropping traditional schizophrenia subtypes from DSM-5 and ICD-11. Schizophr Bull, 2013. 39(4): p. 751–3.

13. Wen, J., et al., Multi-scale semi-supervised clustering of brain images: Deriving disease subtypes. Med Image Anal, 2022. 75: p. 102304.

14. Lalousis, P.A., et al., Heterogeneity and Classification of Recent Onset Psychosis and Depression: A Multimodal Machine Learning Approach. Schizophr Bull, 2021. 47(4): p. 1130–1140.

15. Chand, G.B., et al., Two distinct neuroanatomical subtypes of schizophrenia revealed using machine learning. Brain, 2020. 143(3): p. 1027–1038.

16. Yang, Z., et al., A deep learning framework identifies dimensional representations of Alzheimer’s Disease from brain structure. Nat Commun, 2021. 12(1): p. 7065.

17. Dwyer, D.B., et al., Brain Subtyping Enhances The Neuroanatomical Discrimination of Schizophrenia. Schizophr Bull, 2018. 44(5): p. 1060–1069.

18. Luo, C., et al., Subtypes of schizophrenia identified by multi-omic measures associated with dysregulated immune function. Mol Psychiatry, 2021. 26(11): p. 6926–6936.

19. Young, A.L., et al., Uncovering the heterogeneity and temporal complexity of neurodegenerative diseases with Subtype and Stage Inference. Nat Commun, 2018. 9(1): p. 4273.

20. Vogel, J.W., et al., Four distinct trajectories of tau deposition identified in Alzheimer’s disease. Nat Med, 2021. 27(5): p. 871–881.

21. Young, A.L., et al., Characterizing the Clinical Features and Atrophy Patterns of MAPT-Related Frontotemporal Dementia With Disease Progression Modeling. Neurology, 2021. 97(9): p. e941–e952.

22. Jiang, Y., et al., Neuroimaging biomarkers define neurophysiological subtypes with distinct trajectories in schizophrenia. Nature Mental Health, 2023. 1(3): p. 186–199.

23. van Erp, T.G.M., et al., Cortical Brain Abnormalities in 4474 Individuals With Schizophrenia and 5098 Control Subjects via the Enhancing Neuro Imaging Genetics Through Meta Analysis (ENIGMA) Consortium. Biol Psychiatry, 2018. 84(9): p. 644–654.

24. van Erp, T.G., et al., Subcortical brain volume abnormalities in 2028 individuals with schizophrenia and 2540 healthy controls via the ENIGMA consortium. Mol Psychiatry, 2016. 21(4): p. 585.

25. Okada, N., et al., Subcortical volumetric alterations in four major psychiatric disorders: a mega-analysis study of 5604 subjects and a volumetric data-driven approach for classification. Mol Psychiatry, 2023.

26. Koshiyama, D., et al., White matter microstructural alterations across four major psychiatric disorders: mega-analysis study in 2937 individuals. Mol Psychiatry, 2020. 25(4): p. 883–895.

27. Howes, O.D., et al., Neuroimaging in schizophrenia: an overview of findings and their implications for synaptic changes. Neuropsychopharmacology, 2023. 48(1): p. 151–167.

28. Alnaes, D., et al., Brain Heterogeneity in Schizophrenia and Its Association With Polygenic Risk. JAMA Psychiatry, 2019. 76(7): p. 739–748.

29. Howes, O.D. and S. Kapur, A neurobiological hypothesis for the classification of schizophrenia: type A (hyperdopaminergic) and type B (normodopaminergic). Br J Psychiatry, 2014. 205(1): p. 1–3.

30. Jiang, Y., et al., Progressive Reduction in Gray Matter in Patients with Schizophrenia Assessed with MR Imaging by Using Causal Network Analysis. Radiology, 2018. 287(2): p. 729.

31. Kirschner, M., et al., Orbitofrontal-Striatal Structural Alterations Linked to Negative Symptoms at Different Stages of the Schizophrenia Spectrum. Schizophr Bull, 2021. 47(3): p. 849–863.

32. Thompson, P.M., et al., Mapping adolescent brain change reveals dynamic wave of accelerated gray matter loss in very early-onset schizophrenia. Proc Natl Acad Sci U S A, 2001. 98(20): p. 11650–5.

33. Thompson, P.M., et al., Time-lapse mapping of cortical changes in schizophrenia with different treatments. Cereb Cortex, 2009. 19(5): p. 1107–23.

34. Fillman, S.G., et al., Elevated peripheral cytokines characterize a subgroup of people with schizophrenia displaying poor verbal fluency and reduced Broca’s area volume. Mol Psychiatry, 2016. 21(8): p. 1090–8.

35. Crow, T.J., Is schizophrenia the price that Homo sapiens pays for language? Schizophr Res, 1997. 28(2-3): p. 127–41.

36. Palaniyappan, L. and P.F. Liddle, Does the salience network play a cardinal role in psychosis? An emerging hypothesis of insular dysfunction. J Psychiatry Neurosci, 2012. 37(1): p. 17–27.

37. Del Re, E.C., et al., Baseline Cortical Thickness Reductions in Clinical High Risk for Psychosis: Brain Regions Associated with Conversion to Psychosis Versus Non-Conversion as Assessed at One-Year Follow-Up in the Shanghai-At-Risk-for-Psychosis (SHARP) Study. Schizophr Bull, 2021. 47(2): p. 562–574.

38. Pantelis, C., et al., Neuroanatomical abnormalities before and after onset of psychosis: a cross-sectional and longitudinal MRI comparison. Lancet, 2003. 361(9354): p. 281–8.

39. Slifstein, M., et al., Deficits in prefrontal cortical and extrastriatal dopamine release in schizophrenia: a positron emission tomographic functional magnetic resonance imaging study. JAMA Psychiatry, 2015. 72(4): p. 316–24.

40. Steen, R.G., et al., Brain volume in first-episode schizophrenia: systematic review and meta-analysis of magnetic resonance imaging studies. Br J Psychiatry, 2006. 188: p. 510–8.

41. van Erp, T.G., et al., Subcortical brain volume abnormalities in 2028 individuals with schizophrenia and 2540 healthy controls via the ENIGMA consortium. Mol Psychiatry, 2016. 21(4): p. 547–53.

42. Balu, D.T., et al., Multiple risk pathways for schizophrenia converge in serine racemase knockout mice, a mouse model of NMDA receptor hypofunction. Proc Natl Acad Sci U S A, 2013. 110(26): p. E2400–9.

43. McCutcheon, R.A., T. Reis Marques, and O.D. Howes, Schizophrenia-An Overview. JAMA Psychiatry, 2020. 77(2): p. 201–210.

44. Brugger, S.P., et al., Heterogeneity of Striatal Dopamine Function in Schizophrenia: Meta-analysis of Variance. Biol Psychiatry, 2020. 87(3): p. 215–224.

45. Banaj, N., et al., Cortical morphology in patients with the deficit and non-deficit syndrome of schizophrenia: a worldwide meta- and mega-analyses. Mol Psychiatry, 2023.

46. Mouchlianitis, E., R. McCutcheon, and O.D. Howes, Brain-imaging studies of treatment-resistant schizophrenia: a systematic review. Lancet Psychiatry, 2016. 3(5): p. 451–63.

47. Jiang, Y., et al., Structural and Functional MRI Brain Changes in Patients with Schizophrenia Following Electroconvulsive Therapy: A Systematic Review. Curr Neuropharmacol, 2022. 20(6): p. 1241–1252.

48. Wang, J., et al., ECT-induced brain plasticity correlates with positive symptom improvement in schizophrenia by voxel-based morphometry analysis of grey matter. Brain Stimul, 2019. 12(2): p. 319–328.

49. Jiang, Y., et al., Insular changes induced by electroconvulsive therapy response to symptom improvements in schizophrenia. Prog Neuropsychopharmacol Biol Psychiatry, 2019. 89: p. 254–262.

50. Ho, B.C., et al., Long-term antipsychotic treatment and brain volumes: a longitudinal study of first-episode schizophrenia. Arch Gen Psychiatry, 2011. 68(2): p. 128–37.

51. Lewandowski, K.E., et al., Neuroprogression across the Early Course of Psychosis. J Psychiatr Brain Sci, 2020. 5.

52. Tanaka, S.C., et al., A multi-site, multi-disorder resting-state magnetic resonance image database. Sci Data, 2021. 8(1): p. 227.

53. Keator, D.B., et al., The Function Biomedical Informatics Research Network Data Repository. Neuroimage, 2016. 124(Pt B): p. 1074–1079.

54. Gollub, R.L., et al., The MCIC collection: a shared repository of multi-modal, multi-site brain image data from a clinical investigation of schizophrenia. Neuroinformatics, 2013. 11(3): p. 367–88.

55. Alpert, K., et al., The Northwestern University Neuroimaging Data Archive (NUNDA). Neuroimage, 2016. 124(Pt B): p. 1131–1136.

56. Kogan, A., et al., Northwestern University schizophrenia data sharing for SchizConnect: A longitudinal dataset for large-scale integration. Neuroimage, 2016. 124(Pt B): p. 1196–1201.

57. Poldrack, R.A., et al., A phenome-wide examination of neural and cognitive function. Sci Data, 2016. 3: p. 160110.

58. Repovs, G. and D.M. Barch, Working memory related brain network connectivity in individuals with schizophrenia and their siblings. Front Hum Neurosci, 2012. 6: p. 137.

59. Soler-Vidal, J., et al., Brain correlates of speech perception in schizophrenia patients with and without auditory hallucinations. PLOS ONE, 2022. 17(12): p. e0276975.

60. Kay, S.R., A. Fiszbein, and L.A. Opler, The positive and negative syndrome scale (PANSS) for schizophrenia. Schizophr Bull, 1987. 13(2): p. 261–76.

61. Lindenmayer, J.P., R. Bernstein-Hyman, and S. Grochowski, Five-factor model of schizophrenia. Initial validation. J Nerv Ment Dis, 1994. 182(11): p. 631–8.

62. Rolls, E.T., et al., Automated anatomical labelling atlas 3. Neuroimage, 2020. 206: p. 116189.

63. Pomponio, R., et al., Harmonization of large MRI datasets for the analysis of brain imaging patterns throughout the lifespan. Neuroimage, 2020. 208: p. 116450.

64. Desikan, R.S., et al., An automated labeling system for subdividing the human cerebral cortex on MRI scans into gyral based regions of interest. Neuroimage, 2006. 31(3): p. 968–80.

65. Iglesias, J.E., et al., A computational atlas of the hippocampal formation using ex vivo, ultra-high resolution MRI: Application to adaptive segmentation of in vivo MRI. Neuroimage, 2015. 115: p. 117–37.

66. Saygin, Z.M., et al., High-resolution magnetic resonance imaging reveals nuclei of the human amygdala: manual segmentation to automatic atlas. Neuroimage, 2017. 155: p. 370–382.

67. Iglesias, J.E., et al., A probabilistic atlas of the human thalamic nuclei combining ex vivo MRI and histology. Neuroimage, 2018. 183: p. 314–326.

68. Iglesias, J.E., et al., Bayesian segmentation of brainstem structures in MRI. Neuroimage, 2015. 113: p. 184–95.

