## Supplementary Tables for "Two neurostructural subtypes: results of machine learning on brain images from 4,291 individuals with schizophrenia"

**Supplementary Table S1. Sample demographics by participating cohort.**

| Cohort | Location | N | | Removed subjects by quality control | Mean Age (SZ) | Mean Age (HC) | F/M (SZ) | F/M (HC) | Mean Duration Of Illness (years) | PANSS Positive Subscale | PANSS Negative Subscale | PANSS General Subscale | PANSS Total Score | SAPSTOT | SANSTOT |
|---|---|---|---|---|---|---|---|---|---|---|---|---|---|---|---|
| | Country/Region | Case | Control | | | | | | | | | | | | |
| ASRB | Australia | 68 | 71 | 0 | 41.7±11.1 | 36.0±11.0 | 28/40 | 32/39 | 18.8±9.5 | 13.8±5.7 | 14.5±6.1 | 27.2±9.4 | 55.5±18.4 | 14.7±11.3 | 20.5±12.9 |
| ESO | Czech Republic | 40 | 40 | 0 | 29.5±7.0 | 29.1±6.5 | 20/20 | 20/20 | 0.6±0.8 | 14.2±5.6 | 16.1±5.1 | 33.6±9.5 | 63.8±17.4 | - | - |
| FIDMAG | Spain | 158 | 123 | 1 | 39.5±11.8 | 37.5±10.1 | 35/123 | 69/54 | 15.4±11.2 | 16.8±5.7 | 22.6±6.8 | 36.8±9.3 | 76.2±18.2 | - | 37.2±13.0 |
| FOR2107-MR | German | 90 | 563 | 4 | 38.2±11.9 | 36.3±13.0 | 43/47 | 347/216 | 16.0±10.7 | - | - | - | - | 12.9±13.2 | 13.1±10.3 |
| FOR2017-MS | German | 37 | 386 | 3 | 36.8±11.0 | 31.3±12.2 | 18/19 | 255/131 | 15.3±10.1 | - | - | - | - | 15.2±13.3 | 14.2±14.4 |
| IMH | Singapore | 141 | 0 | 0 | 33.2±9.0 | - | 46/95 | - | 6.5±7.2 | 10.7±3.9 | 9.0±3.0 | 20.5±3.8 | 40.2±8.5 | - | - |
| JBUN | Korea | 116 | 0 | 2 | 34.8±12.9 | - | 60/56 | - | 5.7±7.6 | 14.0±6.4 | 13.2±7.0 | 27.0±8.6 | 54.2±19.1 | - | - |
| OLIN | USA | 73 | 264 | 2 | 41.1±13.1 | 39.9±14.0 | 29/44 | 124/140 | 16.9±12.8 | - | - | - | - | - | - |
| Osaka | Japan | 310 | 1180 | 8 | 35.4±12.8 | 33.6±14.1 | 143/167 | 581/599 | 11.4±9.4 | 19.6±5.9 | 20.7±5.8 | 44.4±11.0 | 84.7±21.0 | - | - |
| PENS | USA | 20 | 16 | 0 | 47.4±9.5 | 45.9±10.1 | 7/13 | 10/6 | 21.7±9.4 | - | - | - | - | 22.0±14.7 | 26.7±13.7 |
| PHCP | USA | 41 | 38 | 0 | 42.2±11.6 | 38.4±13.7 | 11/30 | 17/21 | 17.2±10.4 | - | - | - | - | 21.1±14 | 35.6±14.1 |
| RomeSL | Italy | 162 | 114 | 8 | 39.5±11.3 | 37.4±11.6 | 53/109 | 43/71 | 15.0±10.7 | 20.9±6.4 | 21±7.4 | 45.0±11.5 | 86.9±20.2 | 31.7±19.5 | 28.9±16.0 |
| SoCAT | Turkey | 113 | 79 | 5 | 36.6±8.4 | 37.3±9.0 | 37/76 | 31/48 | 14.9±6.8 | 11.6±5.1 | 19.7±7.5 | 30.2±6.3 | 61.3±13.9 | - | 44.2±21.4 |
| SWIFT | Switzerland | 24 | 13 | 0 | 34.2±10.9 | 29.3±4.0 | 7/17 | 8/5 | 9.5±7.8 | 16.4±5.4 | 12.8±5.0 | 28.8±6.9 | 58.0±12.0 | - | - |
| UCISZ | USA | 26 | 29 | 0 | 42.9±10.8 | 41.7±12.3 | 5/21 | 7/22 | 17.3±10.2 | 15.5±4.2 | 16.2±6.0 | 28.2±6.9 | 59.9±12.2 | 14.4±9.2 | 24.0±14.5 |
| UNINA | Italy | 49 | 55 | 0 | 37.5±9.7 | 42.4±15.7 | 15/34 | 26/29 | 14.7±8.2 | 19.2±5.3 | 22.3±5.9 | 44.0±9.8 | 85.5±17.2 | - | - |
| Zurich | Switzerland | 60 | 28 | 0 | 30.5±8.5 | 32.5±9.3 | 15/45 | 10/18 | 8.4±7.3 | 10.7±2.7 | 14.5±5.9 | 23.4±4.8 | 48.7±10.4 | - | 24.9±15.7 |
| COBRE* | USA | 79 | 86 | 9 | 37.8±13.8 | 38.7±11.9 | 15/64 | 23/63 | 15.6±12.6 | 15.2±5.0 | 15.2±5.4 | 29.2±9.1 | 59.6±15.6 | - | - |
| TOPSY | Canada | 58 | 31 | 4 | 23.3±4.6 | 21.5±3.5 | 12/46 | 12/19 | 0.6±1.1 | - | - | - | - | - | - |
| Voices | Australia | 38 | 47 | 4 | 43.4±10.4 | 31.5±12.0 | 21/17 | 25/22 | 1.8±0.9 | 17.8±5.6 | 13.8±4.8 | 34.2±10.6 | 65.9±18.2 | 18.9±14.0 | 17.7±12.9 |
| OSLO_TOP | Norway | 389 | 769 | 5 | 31.2±9.3 | 33.5±8.8 | 159/230 | 344/425 | - | - | - | - | - | - | - |
| CN-Shanghai1* | China | 361 | 267 | 7 | 24.3±8.0 | 24.0±4.9 | 167/194 | 144/123 | 0.9±1.4 | 20.2±5.7 | 15.4±7.4 | 38.1±8.1 | 73.7±15.7 | - | - |
| CN-Shanghai2 | China | 159 | 133 | 1 | 24.9±7.0 | 23.7±5.9 | 84/75 | 67/66 | 1.4±2.0 | 23.7±5.3 | 18.1±6.8 | 41.4±7.2 | 83.2±13.7 | - | - |
| CN-Shanghai3 | China | 42 | 23 | 0 | 29.9±7.4 | 31.2±5.9 | 23/19 | 12/11 | 6.6±5.7 | 19.9±3.1 | 18.4±6.4 | 33.1±4.9 | 71.4±9.0 | - | - |
| CN-Harbin | China | 49 | 0 | 0 | 26.7±8.1 | - | 22/27 | - | 2.4±3.2 | 20.2±4.5 | 22.4±6.4 | 40.4±7.4 | 83.0±14.5 | - | - |
| CN-Chengdu* | China | 101 | 127 | 3 | 40.2±11.8 | 38.1±14.9 | 31/70 | 42/85 | 15.5±10.3 | 13.2±5.8 | 21.0±6.3 | 28.2±5.7 | 62.3±13.1 | - | - |
| CN-Taibei* | China | 158 | 254 | 8 | 43.9±11.0 | 38.0±12.0 | 93/65 | 136/118 | 15.7±10.1 | 9.6±3.2 | 10.0±5.5 | 20.7±4.4 | 40.3±10.7 | - | - |
| CN-Zhengzhou* | China | 194 | 59 | 1 | 23.2±8.6 | 26.5±5.5 | 97/97 | 32/27 | - | 22.3±5.1 | 23.1±6.3 | 44.1±7.9 | 89.5±15.0 | - | - |

| | | | | | | | | | | | | | | | |
|---|---|---|---|---|---|---|---|---|---|---|---|---|---|---|---|
| CN-Beijing1 | China | 130 | 0 | 9 | 21.3±4.2 | - | 75/55 | - | - | 22.4±5.0 | 20.9±7.2 | 38.8±7.0 | 82.1±15.1 | - | - |
| CN-Beijing2 | China | 217 | 721 | 3 | 26.3±6.6 | 24.8±4.2 | 85/132 | 365/356 | - | - | - | - | - | - | - |
| CN-Changsha | China | 120 | 92 | 2 | 23.4±5.5 | 23.5±5.2 | 39/81 | 51/41 | - | 15.3±5.8 | 18.3±8.8 | 31.9±10.6 | 65.4±22.0 | - | - |
| CN-Xian | China | 39 | 51 | 0 | 23.9±8.6 | 21.1±4.0 | 17/22 | 18/33 | 1.2±1.8 | 22.9±4.9 | 21.4±6.4 | 44.8±8.8 | 89.1±14.6 | - | - |
| HCP-EP | USA | 54 | 39 | 1 | 21.9±3.0 | 25.4±4.4 | 15/39 | 14/25 | - | 11.7±3.9 | 16.0±5.6 | 24.9±4.5 | 52.7±10.1 | - | - |
| JP-SRPBS | Japan | 139 | 887 | 10 | 38.3±10.8 | 36.5±15.5 | 58/81 | 382/505 | 14.4±9.6 | 14.6±5.3 | 17.0±6.4 | 31.4±9.4 | 62.9±18.9 | - | - |
| fBIRN* | USA | 42 | 65 | 6 | 38.1±11.8 | 38.3±11.2 | 14/28 | 29/36 | - | - | - | - | - | - | - |
| MCIC | USA | 109 | 95 | 1 | 33.8±11.2 | 32.9±12.2 | 26/83 | 30/65 | 11.1±10.9 | - | - | - | - | - | - |
| NMorphCH* | USA | 44 | 43 | 0 | 32.5±6.9 | 31.5±8.4 | 14/30 | 22/21 | - | - | - | - | - | - | - |
| NUSDAST* | USA | 123 | 127 | 1 | 33.8±12.5 | 32.6±13.8 | 42/81 | 62/65 | - | - | - | - | - | - | - |
| DS000030 | USA | 50 | 121 | 1 | 36.5±8.9 | 31.6±8.8 | 12/38 | 56/65 | - | - | - | - | - | - | - |
| DS000115* | USA | 22 | 18 | 0 | 24.2±3.8 | 20.7±4.8 | 6/16 | 8/10 | - | - | - | - | - | - | - |
| DS004302 | Spain | 46 | 24 | 0 | 42.5±10.7 | 39.5±14.3 | 10/36 | 7/17 | - | - | - | - | - | - | - |
| Total | - | 4291 | 7078 | 109 | 32.5±11.9 | 33.0±12.7 | 1709/2582 | 3461/3617 | 10.5±10.4 | 17.2±6.8 | 17.5±7.6 | 34.8±11.6 | 69.5±22.4 | 22.0±17.5 | 27.1±18.0 |

Note: * some of these cohorts overlap with a previous paper (https://doi.org/10.1038/s44220-023-00024-0)

### Supplementary Table S2. Dataset-specific information.

| Cohort | Diagnosis measurement | Exclusion/inclusion criteria | Scanner manufacturer and type | Imaging protocols |
|---|---|---|---|---|
| ASRB | ICD-10 | All participants were fluent English speakers and aged 18-65 years old. No history of an organic brain disorder, brain injury accompanied by > 24 h of amnesia, mental retardation defined as an IQ < 70, movement disorder, current substance dependence, or electro-convulsive therapy in the preceding 6 months. The control participants additionally had no personal history of psychotic disorder or family history of psychotic disorder in their first-degree biological relatives. | 1.5T Siemens Avanto | High-resolution T1-weighted structural magnetic resonance imaging (sMRI) brain scans (MPRAGE) were acquired using an optimized magnetization prepared rapid acquisition gradient echo on 1.5 T Siemens Avanto scanners (Siemens, Erlangen, Germany) across five Australian research sites. Image parameters were set to 176 slices of 1mm thickness, no gap with field-of-view 250 x 250 mm2, repetition time 1980 ms, echo time 4.3 ms, data acquisition matrix 256 x 256, with a flip matrix of 15°, resulting in a voxel size of 0.98×0.98×1.0mm3. |
| ESO | ICD-10 (F20.x, F23, F25) | Early or first-episode psychosis, Czech language as a mother tongue, 18-60 years old. Neurocognitive disorders (organic mental disorder), mental disorders caused by addiction, mental retardation (IQ<80), severe neurological disorder, head injury, hypertension, cerebrovascular disease, epilepsy, migraine, endocrine disorders. | 3T Siemens Tim Trio | MP-RAGE 3D, 1mm thickness, acquisition matrix 256 x 256, TR=2300ms, TE=4.63ms,TI=900ms |
| FIDMAG | DSM-IV criteria based on interview and review of clinical history | Patients had a diagnosis of schizophrenia. All participants were in the 18-65 age range. Controls were excluded if they reported a history of mental illness and/or treatment with psychotropic medication. Patients were excluded if have had a history of brain trauma or neurological disease or had shown alcohol/ substance abuse within 12 months before participation | 1.5T GE Signa | 180 axial slices; 1mm slice thickness, no gap, matrix size 512x512; 0.5x0.5x1mm3 voxel resolution; TE 4ms, TR 2000ms, flip angle 15. |
| FOR2107-MR | DSM-IV-TR using SCID-I semi-structured interview | All participatnts were in the 18-65 age range; patients were diagnosed of schizophrenia, schizoaffective disorder or any schizophreniform disorder. Exclusion criteria all: any history of neurological (head trauma or unconsciousness) and medical condition (severe somatic disorders), IQ<80; Exclusion criteria controls: any current or former psychiatric disorder; Exclusion criteria patients: Current benzodiazepine treatment (wash out of at least three half-lives before study participation). | 3T Siemens Magnetom TrioTim Syngo | MPRAGE imaging sequence. 1 acquisition. Flip angle: 9 degrees. TE: 2.26 ms. TR: 1900 ms. TI: 900 ms. Acceleration factor: 2Field of view: 256. Image dimensions: 256x256x176 voxels. Voxel size: 1x1x1 mm. |
| FOR2017-MS | DSM-IV-TR using SCID-I semi-structured interview | All participatnts were in the 18-65 age range; patients were diagnosed of schizophrenia, schizoaffective disorder or any schizophreniform disorder. Exclusion criteria all: any history of neurological (head trauma or unconsciousness) and medical condition (severe somatic disorders), IQ<80; Exclusion criteria controls: any current or former psychiatric disorder; Exclusion criteria patients: Current benzodiazepine treatment (wash out of at least three half-lives before study participation). | 3T Siemens PRISMA | MPRAGE imaging sequence. 1 acquisition. Flip angle: 8 degrees. TE: 2.28 ms. TR: 2130 ms. TI: 900 ms. Acceleration factor: 2. Field of view: 256. Image dimensions: 256x256x192 voxels. Voxel size: 1.0x1.0x1.0mm |

| | | | | |
|---|---|---|---|---|
| IMH | SCID DSM-IV Axis I Disorders | (1) Diagnosis of SZ (Patients), (2) Age: 21-65, (3) English speaking, (4) Provision of informed written consent. History of significant head injury; significant Neurological diseases (such as epilepsy, cerebrovascular accident) or Medical Illnesses; significant DSM IV alcohol or substance use or dependence; contraindications to MRI (e.g. pacemaker, orbital foreign body, recent surgery/procedure with metallic devices/implants deployed); pregnant women; claustrophobia. | 3T Philips Achieva | T1 scans: 180 axial slices of 0.9mm thickness with no gap, FOV = 230x230 mm2, matrix 256x204, voxel size =0.89x0.89x0.9 mm3, TR=7.2 s, TE=3.3 ms, FA=8°. |
| JBUN | DSM-V | A diagnosis of schizophrenia: all patients were between 18 – 71 years old. Patients with an (a) alcohol or substance use disorder; (b) intellectual disability (IQ ≤70); (c) current or past neurological disease, serious medical illness, or pregnancy; and (d) claustrophobia were excluded. | 3T Siemens MAGNETOM Verio syngo | T1-weighted, 3-dimensional Magnetization Prepared Rapid Gradient Echo (TR = 1900ms; TE=2.45ms; FOV=250mm; FA=9°; voxel size=1x1x1mm3; Slice thickness=1mm). |
| OLIN | SCID | SCID-NP | 3T Siemens Allegra | T1-weighted, 3D magnetization prepared rapid gradient-echo (MPRAGE) sequence (TR/TE/TI=2200/4.13/766 ms, flip angle=13°, voxel size [isotropic]=0.8mm, image size=240 x 320 x 208 voxels), with axial slices parallel to the AC-PC line. |
| Osaka | SCID-P; DSM-IV | SCID-NP | 1.5T GE Signa Excite; 3T GE Signa HDxt | 3D-IR-FSPGR, TR/TE/TI=12.6/4.2/400ms, flip angle=15°, 256x256x124 matrix, FOV=240x240mm, slice thickness=1.4mm, Nex=1, No Asset, QD Head coil; 3D-IR-FSPGR, TR/TE/TI=7.2/2.9/400ms, flip angle=11°, 256x256x172 matrix, FOV=240x240mm, slice thickness=1.0mm, Nex=1, No Asset, 8ch Brain coil. |
| PENS | DSM-IV | Same as PHCP study: Adults aged 18–65, including people with major mental illness (schizophrenia, schizoaffective disorder, bipolar I disorder with psychotic features), their first-degree biological relatives, and unrelated healthy controls. | 3 Tesla Siemens Prisma scanner using a 32 channel head coil | Structural MRI using a 10-min T1-weighted MPRAGE sequence (TE = 2.12 ms, TR = 2,400 ms, flip angle = 8, resolution = 256) |
| PHCP | DSM-IV | All participants spoke English as their primary language and did not have: a legal guardian (or otherwise lack capacity to provide informed consent), alcohol/drug abuse in the past month or alcohol/drug dependence in the last 6 months, a diagnosed Learning Disability or estimated IQ lower than 70 (if either condition was diagnosed based on testing by a trained professional or the latter by research staff), a current or past central nervous system disease (including: seizures, epilepsy, encephalitis, MS, Parkinson's, stroke), history of head injury with skull fracture or loss of consciousness greater than 30 min, history of electro-convulsive therapy (ECT) in the last year, tardive dyskinesia (as evidenced by medical record), obstructed or compromised vision (e.g., lazy eye that is uncorrected or was corrected after age 17 / strabismus / cross eyes / permanent eye injury / abnormality in visual field / cataract), hearing problems (e.g., cannot hear without hearing aid / severe tinnitus), or a condition likely making it impossible to perform tasks (e.g., paralysis, severe arthritis). | Siemens 3 T Prisma scanner with a Siemens 32 channel head coil | A multi-echo T1w MPRAGE sequence and a variable-flip-angle, turbo-spin-echo T2w scan with volumetric navigators to aid real-time motion correction and selective reacquisition were acquired with scanning protocol identical to that of the Lifespan Human Connectome Project (see Harms et al., 2018). As in the Lifespan HCP (Harms et al., 2018), up to 30 k-space lines for the T1w scan and up to 25 k -space lines for the T2w scan were allotted for reacquisition. T1w MPRAGE multi-echo (300 × 320 matrix, FOV=240 × 256mm, resolution=0.8mm, flip angle=8, TE=1.81, 3.6, 5.39, 7.18 ms, TR=2500ms, slices/orientation=208 sag, AF=2, time=8min:22sec |

| | | | | |
|---|---|---|---|---|
| | | Additionally, patients were between the ages of 18 and 65 years old with a diagnosis of schizophrenia, schizoaffective disorder, or bipolar I disorder with a history of psychotic symptomatology (i.e., delusions or hallucinations) with no indication that symptoms were caused by substance use or a general medical condition. While patients were screened and excluded for current substance use issues, a history of such issues as well as current/lifetime comorbidities of any kind were permitted for enrollment in the study in order to have a sample representative of patients with psychosis in the general population while simultaneously limiting nuisance effects. Finally, as this was a family study, patients and relatives were not adopted.<br><br>Biological relatives were 18–69 years old with a first-degree biological relative with schizophrenia, schizoaffective disorder, or bipolar I disorder with psychosis, and living within one day’s drive or planning to visit the vicinity of the University of Minnesota. Because relatives included parents of PwP, we expanded the age range of the relatives group to accommodate recruitment efforts. Relatives were enrolled regardless of psychopathology. Approximately half (45.95%) of the first-degree biological relatives in the study carried their own mental health diagnosis (e.g., major depression). Occasionally, relatives with substance dependence in partial remission (3 participants, 3.9%), current substance dependence (1 participant, 1.3%), or psychotic psychopathology (1 participant, 1.3%) were included in the study.<br><br>Controls were aged 18–65 and had no history of schizophrenia, schizoaffective disorder, or bipolar I disorder with psychotic features, or other psychotic symptoms or history of major depressive disorder. Additionally, controls had no first-degree biological relative with a history of psychiatric hospitalization for a psychotic or affective disorder. | | |
| RomeSL | SCID DSM-IV Axis I Disorders (SCID-I) and SCID DSM-IV Axis II Personality Disorders (SCID-II) | Inclusion criteria were (i) age between 18 and 65 years; (ii) at least five years of education; and (iii) suitability for MRI scanning. Exclusion criteria were (i) history of alcohol or drug abuse in the two years before the assessment; (ii) lifetime drug dependence; (iii) traumatic head injury with loss of consciousness; (iv) past or present major medical illness or neurological disorders; (v) any (for HC) or additional (for patients) psychiatric disorder or mental retardation; (vi) dementia or cognitive deterioration according to DSM-IV-TR criteria, and Mini-Mental State Examination (MMSE) score < 25, consistent with normative data in the Italian population; (vii) not able and willing to give written informed consent. | Siemens 3T Allegra | 3D MPRAGE: TE/TR = 2.4/7.92 ms, flip angle=15°, voxel size 1×1×1 mm. |

| | | | | |
|---|---|---|---|---|
| SoCAT | DSM-V; DSM-IV | Inclusion: (1) Aged between 19-45, (2) had been followed up for at least one year with schizophrenia diagnosis according to the medical records, (3) had confirmed diagnosis of schizophrenia and no other axis I diagnosis after evaluation with SCID, (4) were clinically stable over the past three months (no change in symptoms requiring interventions such as medication change or hospitalization) and had cognitive abilities enough to read and understand the informed consent form. Exclusion: (1) an unstable medical disease (e.g., diabetes mellitus, hypertension, etc.), (2) a history of head trauma with loss of consciousness, (3) contraindications to MRI (e.g. pacemaker, orbital foreign body, recent surgery/procedure with metallic devices/implants deployed); pregnant women; claustrophobia. | 3T Siemens Magnetom Verio; 1.5T Siemens SymphonyVision | Sagittal T1-weighted 3D magnetization-prepared rapid gradient echo acquisition (MPRAGE) sequence (TE = 2.21 msec, TR = 1600 msec, TI = 900 msec, FA = 9, FOV = 256, voxel size = 0.5x0.5x1 mm, number of slices = 160, no inter-slice gap). Sagittal T1-weighted 3D magnetization-prepared rapid gradient echo acquisition (MPRAGE) sequence (TE = 3.93 msec, TR = 2300 msec, TI = 1100 msec, FA = 12, FOV = 256, voxel size = 0.5x0.5x1 mm, number of slices = 160, no inter-slice gap). |
| SWIFT | SCID DSM-IV-TR | Age between 18-65 y/o, good German language skills that allowed them to understand the consent procedure and to undergo the clinical assessment, right-handedness according to the Edinburgh Inventory. For patients: Diagnosis with a schizophrenia spectrum disorder. Participants were excluded if they were left-handed, pregnant, showed any contraindications for MRI (e.g. metal-containing implants such as pacemaker or cochlear implants, claustrophobia), had a history of serious neurological issues, or reported current abuse of alcohol and/or psychoactive substances (apart from nicotine). Additionally, controls had no current major psychiatric DSM-IV Axis I diagnoses, as assessed with the screening questionnaire of the Structured Clinical Interview for DSM-IV Axis I Disorders. | 3T Siemens Verio | MPRAGE: 160 sagittal slices, 1mm slice thickness, 256x256 matrix size, 1x1x1 mm3 voxel size. TR = 2.3ms, TE = 2.98ms, TI = 900ms. |
| UCISZ | SCID DSM-IV-TR criteria | All subjects diagnosed with schizophrenia were clinically stable outpatients whose antipsychotic medications and doses had not changed within the last two months. Schizophrenia and healthy volunteers with a history of major medical illness, drug dependence in the last five years (except for nicotine), current substance abuse disorder, or MRI contraindications, were excluded. Individuals with schizophrenia who had significant tardive dyskinesia and healthy volunteers with a current or past history of major neurological or psychiatric illness or with a first-degree relative with an Axis-I psychotic disorder diagnosis were also excluded. | 3T Philips Achieva | T1TFE:200 sagittal slices, 320x274 matrix size, .75mm isotropic, TR = 11ms, TE=4.562ms, flip angle = 18° |
| UNINA | DSM-5 | Schizophrenia:(1) Diagnosis of SZ (Patients); (2) Age: 18-60; (3) Provision of informed written consent; (4) Disease duration >2 years; (5) no medication switch or dose changes in the last 6 months (i.e., >10% baseline dose); (6) no evidence of current or recent (3 months) worsening of psychotic symptoms; absence of: (7) macroscopic brain structural anomalies; (8) major systemic disorder (such as cardiovascular, endocrine, metabolic disorders); (9) other psychiatric disorder (including addiction disorder, substance use disorder, or frequent substance use in the 6 months preceding the recruitment); (10) moderate or severe neurological disorder; (11) intellectual disability; (12) pregnancy or lactation; (13) enrollment in any sort of experimental clinical trial within 3 months | 3T Siemens Tim Trio | 3D T1-weighted Magnetization Prepared Rapid Acquisition Gradient Echo sequence (MPRAGE; TR=1900 ms; TE=3.4 ms; TI=900 ms; Flip Angle=9°; resolution=1x1x1 mm3; 160 axial slices). |

| | | | | |
|---|---|---|---|---|
| | | from recruitment.<br>Controls:(1) No psychiatric diagnosis; (2) Age: 18-60; (3) Provision of informed written consent; 4) Absence of: macroscopic brain structural anomalies; major systemic disorder (such as cardiovascular, endocrine, metabolic disorders); moderate or severe neurological disorder; intellectual disability; pregnancy or lactation | | |
| Zurich | MINI DSM IV | A diagnosis of schizophrenia. We excluded patients with any other DSM-IV Axis I disorder (in particular, current substance use disorder and major depressive disorder), those medicated with lorazepam at a dose higher than 1 mg, those with florid psychotic symptoms (i.e., any positive subscale item scores higher than 4 on the PANSS scale and those with extrapyramidal side effects (i.e., a total score higher than 2 on the MSAS). Healthy controls were screened for any neuropsychiatric disorders using the structured Mini-International Neuropsychiatric Interview to ensure that they had no previous or present psychiatric illness. Both patients and healthy controls were required to have a normal physical and neurologic status and no history of major head injury or neurologic disorder. | 3T Philips | 3D T1-weighted images were acquired with an ultra-fast gradient echo T1-weighted sequence (TR=8.4ms, TE=3.8ms, flip angle=8°) in 160 sagittal plan slices (1mm slice thickness, no slice gap) of 240×240mm2 resulting in 1x1x1mm3voxels. |
| COBRE | SCID DSM-IV Axis I Disorders | All participants were in the 18-65 age range and had a diagnosis of schizophrenia. Healthy individuals were included if they did not have a personal or family history of psychiatric disorders. History of neurological disorder, history of mental retardation, history of severe head trauma with more than 5 minutes loss of consciousness, history of substance abuse or dependence within the last 12 months and MRI contraindications. | 3T Siemens TIM Trio | T1-weighted images were acquired with a 5-echo multi-echo MPRAGE sequence [TE (echo times) = 1.64, 3.5, 5.36, 7.22, 9.08 ms, TR (repetition time) = 2.53 s, TI (inversion time) = 1.2 s, 7° flip angle, number of excitations (NEX) = 1, slice thickness = 1 mm, FOV (field of view) = 256 mm, resolution = 256x256]. |
| TOPSY | DSM-V | Inclusion criteria for FEP: individuals experiencing FEP, with lifetime antipsychotic treatment less than 14 days. Exclusion criteria for FEP: meeting criteria for a mood disorder (bipolar or major depressive) with psychotic features, or possible drug-induced psychosis. Healthy control (HC) participants were free from personal history of mental illness or family history of psychotic disorders, matched based on age, sex, and parental education. Exclusion criteria for both FEP and HC: substance use disorder in the past year based on DSM-5 criteria, history of major head injury, significant medical illness, or contraindications to MRI. | Siemens MAGNETOM 7.0T MRI Plus (Siemens, Erlangen, Germany) | Anatomical images were acquired using a sagittal 3D MP2RAGE1 sequence with TE=2.83 ms, TR=6000 ms, TI=800ms/2700 ms, flip angle=4/5 degrees, matrix=320x320x208, iPAT=3, partial Fourier=6/8, voxel size=0.8x0.8x0.8, and a 3D SA2RAGE2, with TE=0.81 ms, TR=2400 ms, TI=45/1800 ms, flip angle=4o/5o, matrix=320x320x208, iPAT=3, partial Fourier=6/8, voxel size=0.8mmx0.8mmx0.8mm. |
| Voices | MINI | Inclusion: diagnosis of schizophrenia or schizoaffective disorder. Age: 18-65, fluent in English, Able to provide informed consent.<br>Exclusion: History of significant head injury or neurological disease affecting cognition (such as epilepsy). Current DSM IV alcohol or substance use or dependence; contraindications to MRI (e.g. pacemaker, orbital foreign body, recent surgery/procedure with metallic devices/implants deployed); currently pregnant | 3T Siemens Tim Trio | Images were optimized using a magnetization-prepared rapid acquisition gradient echo (MP-RAGE) sequence, and consisted of 176 sagittal slices/brain of 1 mm thickness without gap; field of view = 256 × 256 mm2; repetition time/echo time =1900/2.52 ms; data matrix size =256 × 256; voxel dimensions =1.0 × 1.0 ×1.0 mm3. All scans were conducted at a single site. |

| | | | | |
|---|---|---|---|---|
| OSLO_TOP | The Structured Clinical Interview for DSM-IV axis 1 disorders (SCID-IV). | Healthy controls were randomly drawn from the national population registry in the same geographical area as the patients, and invited by letter to participate. They were screened prior to participation. Absence of current or previous history of a psychiatric disorder was determined by self-report. Current symptomatology was screened for on the day of inclusion using the Prime MD and alcohol and drug use were screened for using AUDIT/DUDIT.<br>The exclusion criteria for healthy controls were: - Age outside of the range 18-65 years. - Current or previous psychiatric disorder. - History of severe mental illness in a first-degree relative. - Any alcohol or drug abuse or dependence | 1.5T Siemens Magnetom Sonata | Two sagittal T1-weighted magnetization prepared rapid gradient echo (MPRAGE) volumes were acquired with the Siemens tfl3d1_ns pulse sequence (TE = 3.93 ms, TR = 2730 ms, TI = 1000 ms, flip angle = 7°; FOV = 24 cm, voxel size= 1.33 x 0.94 x 1 mm3, number of partitions = 160). |
| CN-Shanghai1 | DSM-IV | All the subjects were Mandarin-speaking Han Chinese individuals from Shanghai metropolitan area. The FES patients were identified according to DSM-IV criteria by qualified psychiatrists using all available clinical information including a diagnostic interview of patients and their family, clinical case notes, and clinician’s observations. All healthy subjects were assessed in accordance with DSM-IV criteria as being free of schizophrenia and other axis I disorders, and none had neurological diseases, head trauma, or substance abuse. All participants were right-handed; had no history of substance abuse or suicidal ideation; and had no MRI contraindications. | GE Sigma 3T | GE Sigma 3.0 T MR (GE Medical Systems, Milwaukee, Wisconsin). TR = 7.8 ms; TE = 3.65 ms; flip angle = 7°; matrix = 256×256; voxel size = 1×1×1 mm. |
| CN-Shanghai2 | DSM-IV | The exclusion criteria were as follows: (1) brain trauma, substance-related disorders, major medical or neurologic disorders; (2) other mental disorders meeting DSM-IV criteria; (3) drug or alcohol abuse; (4) pregnancy, breastfeeding or other unstable clinical state including aggressive behaviour; (5) history of electroconvulsive therapy or transcranial magnetic stimulation within six months; and (6) other contraindications to MRI scanning. To eliminate potential familial effects, healthy subjects whose first- or second-degree relatives had a history of mental disorders were also excluded. | 3T Siemens MR B17 | A 3-Tesla MRI scanner (Siemens MR B17) was used to acquire data in the Shanghai Mental Health Centre. High-spatial-resolution T1-weighted images were collected by a magnetization-prepared rapid acquisition gradient echo (MPRAGE) sequence. The main parameters included repetition time/echo time, 2530/2.56 msec; flip angle, 7°; field of view, 256 × 256 mm2; matrix size, 256 × 256; section thickness, 1 mm (no gap); and voxel size, 1×1×1mm3. |
| CN-Shanghai3 | DSM-IV-TR | Inpatients with schizophrenia were recruited from the Shanghai Mental Health Center (SMHC). All the patients were recruited from October 2013 to January 2015. All patients met criteria for schizophrenia or schizoaffective disorder based on the Structured Clinical Interview for Diagnostic and Statistical Manual of Mental Disorders (DSM-IVTR) as conducted by senior psychiatrists. The severity of psychotic symptoms was assessed by the Positive and Negative Syndrome Scale (PANSS). All patients had a total PANSS score of 60 or more and had not received ECT during the past six months. All health controls did not have a lifetime psychiatric disorder or family history of psychosis in their first-degree relatives. Participants were excluded if they had brain injuries, organic mental disorders, neurologic abnormalities, other serious physical illnesses, dementia, substance abuse or dependence, or contraindications to MRI. | 3-T Siemens Magnetom Verio Syngo MR B17 | High-resolution T1-weighted images were acquired using a 3-T Siemens Magnetom Verio Syngo MR B17 scanner. High-resolution T1-weighted images were collected with a magnetization-prepared rapid acquisition gradient echo (MPRAGE) sequence (TR = 2530 ms; TE = 2.56 ms; flip angle = 7°; inversion time = 1100 ms; FOV = 256 mm × 256 mm; matrix = 256 × 256; slice thickness = 1 mm; 224 slices; and voxel size = 1.0 × 1.0 × 1.0 mm). |

| | | | | |
|---|---|---|---|---|
| CN-Harbin | DCM-IV SCID | Patient recruitment criterion was based on the Diagnostic and Statistical Manual of Mental Disorders, Fourth Edition and Structured Clinical Interview for DSM (SCID), which includes: (1) Age range between 14-45 years old; (2) Intelligence quotient (IQ)>69; (3) The first onset and no systematic antipsychotic treatment before admission; (4) Psychotic symptom assessment scale (positive and negative symptom scale score) is greater than or equal to 60; clinical overall impression CGI severity is greater than or equal to 4 points; (5) Exclude other Axis I mental disorders except schizophrenia, and have no DSM-5 drug or alcohol dependence within the past three months. Exclusion criteria for patients included the following: (1) Sensory-motor disturbances (hearing impairment, blindness), neurological disorders (brain injury, epilepsy), or other medical conditions (2) claustrophobic and unable to undergo MRI scans; (3) patients with repetitive transcranial magnetic stimulation or MRI Contraindications to scanning, such as those with metal implants in the body; (4) suicide attempters, pregnant or breastfeeding women. | 3.0 Tesla GE Discovery MR750 | All scans were acquired on the same 3.0 Tesla GE Discovery MR750 scanner equipped with a 32-channel head coil. |
| CN-Chengdu | DSM-IV | Patients with SZ were recruited from the Clinical Hospital of Chengdu Brain Science Institute. Each patient was diagnosed based on the Diagnostic and Statistical Manual of Mental Disorders, Fourth Edition (DSM-IV). Subjects with a history of brain injuries, substance-related disorders, major medical or neurological disorder were excluded. To exclude the potential effect of similar genetic backgrounds, the history of psychiatric disorder in a first- or second-degree relative was an additional exclusion criterion for healthy controls. | 3-Tesla GE DISCOVERY MR 750 | A 3-Tesla MRI scanner (GE DISCOVERY MR 750, USA) was used to collect imaging data in the University of Electronic Science and Technology of China. High-resolution T1-weighted images were acquired using a three dimensional fast spoiled gradient echo (T1-3D FSPGR) sequence. The main parameters include: TR = 6.008 ms; TE = 1.984 ms; flip angle (FA) =90°; field of view (FOV) = 25.6 cm × 25.6 cm; matrix size = 256 × 256; slice thickness = 1 mm (no gap). |
| CN-Taibei | DSM-IV | Patients and matched healthy controls were recruited from the Veteran General Hospital in Taipei, Taiwan. All participants were diagnosed according to the Diagnostic and Statistical Manual of Mental Disorder-IV criteria for schizophrenia, and each participant's history of medical disease, psychiatric illness, and medication use was evaluated by interview and medical charts carefully. Any participants with the following conditions were excluded: (1) a comorbid substance-related disorder, (2) presence of neurobiological disorders, such as dementia, head injury, stroke, or Parkinson’s disease; (3) presence of hypertension, diabetes, hyperlipidemia or coronary heart disease; (4) severe medical illness, such as malignancy, heart failure, or renal failure; (4) presence of ferromagnetic foreign bodies or implants that were anywhere in the body. | 3T Siemens | Siemens MAGNETOM Tim Trio 3.0T MRI Scanner (Siemens Healthcare, Erlangen, Germany) with a 12- channel head coil, using 3-dimensional magnetization prepared rapid gradient-echo sequence. TR: 2530 ms, TE: 3.5 ms, TI: 1,100 ms, FoV: 256 mm, flip angle: 7 degree, 192 sagittal slices, voxel size = 1.0 mm3 cubic, no gap. |
| CN-Zhengzhou | DSM-IV | All patients were identified according to the Diagnosis and Statistic Manual of Mental Disorders, fourth edition (DSM-IV) criteria for schizophrenia by qualified psychiatrists using all available clinical information including a diagnostic interview, clinical case notes, and clinician’s observations. The severity of positive and negative symptoms was assessed by trained and experienced psychiatrists using PANSS scoring. Individuals were excluded from the study if they were diagnosed with | GE MR750 3T | GE MR750 3T MRI scanner with standard quadrature head coil. TR/TE= 8.2 / 3.2 ms, FOV= 256×256 mm 2, Matrix= 256×256, slice thickness= 1.0 mm, gap= 0mm, 188 contiguous sagittal slices；flip angle = 12°. |

| | | | | |
|---|---|---|---|---|
| | | schizoaffective disorder, mood disorders, other cognitive disorders, epilepsy, had severe physical diseases, alcohol/drug dependence, or been treated with electroconvulsive therapy. Healthy subjects were assessed in accordance with DSM-IV criteria as being free of schizophrenia and other Axis I disorder, and none had neurological diseases, head trauma, substance abuse, suicidal ideation, and MRI contraindications. | | |
| CN-Beijing1 | DSM-IV | The inclusion criteria were: (1) both in- and out-patients, (2) both genders, (3) diagnosis of schizophrenia established with Structured Clinical Interview for the DSM-IV Axis I Disorder, patient edition, (4) aged between 18 and 45 years, (5) onset at age ≥ 15 years, (6) first episode of schizophrenia, (7) no previous psychiatric treatment ('drug-naïve' status), and (8) ability to understand the contents of interview and provide written informed consent.<br>The exclusion criteria were: (1) a history or diagnosis of major medical conditions, (2) a history of alcohol and / or drug abuse or dependence, and (3) contra-indication to olanzapine, aripiprazole, or risperidone. | Siemens Trio Trim 3.0 T | Main parameters: TR: 9.8 ms, TE: 3.8 ms, TI: 450 ms, FoV: 512 x 512, flip angle: 13, slice thickness=1.2 mm. |
| CN-Beijing2 | DSM-IV | A consensus diagnosis of schizophrenia was made by two experienced senior psychiatrists according to the Diagnosis and Statistic Manual of Mental Disorders, fourth edition (DSM-IV) criteria for schizophrenia or schizophreniform disorder using the Structured Clinical Interview for DSM-IV-TR Axis I Disorders, Patient Edition (SCID-I/P). All subjects initially recruited with schizophreniform disorder were finally diagnosed with schizophrenia after being followed up for at least six months. Individuals were excluded from the study if they were diagnosed with schizoaffective disorder, mood disorders, delusional disorder, brief psychotic disorder, psychosis associated with substance use or medical conditions, learning disability, pervasive developmental disorder, delirium, dementia, amnesia or other cognitive disorders; had severe, unstable physical diseases (such as diabetes, thyroid diseases, hypertension and cardiac diseases), a well-documented history of epilepsy, a DSM-IV diagnosis of alcohol or drug dependence; had been treated with electroconvulsive therapy within the last six months; were pregnant or breastfeeding; had previously attempted suicide; or had experienced symptoms of severe excitement and agitation within one week before MRI scanning. Healthy control individuals were recruited from each hospital and screened using SCID-I, Non-Patient Edition (SCID-I/NP). Individuals with any history of mental disorders or first- or second-degree relatives with any history of mental disorders were excluded. | Siemens Trio Trim 3.0 T | T1-weighted images were collected with matrix size of 256×256, resolution of 1×1 mm2, and slice thickness of 1 mm. |
| CN-Changsha | DSM-IV SCID-P | The inclusion/exclusion criteria of patients: (1) diagnosis of SCZ confirmed using the Structured Clinical Interview for DSM-IV-patient version (SCID-P) 1; (2) Han Chinese ethnicity and right-handed; (3) minimum 9 years of school education (4) good general physical health with no known | Philips Gyroscan Achieva 3T | 3-D structural MRI images (T1-weighted) were acquired from the sagittal plane using spoiled gradient echo (SPGR) pulse sequence on a Philips Gyroscan Achieva 3T MRI scanner, scanning parameter: TR=12 ms, |

| | | | | |
|---|---|---|---|---|
| | | endocrine, metabolic or inflammatory disorders; (5) no known neurological disorder; (6) no substance dependence in the last year; (7) benzodiazepine treatment, if any, stopped more than 24 h prior to scanning and (8) no contraindications for MRI.<br>The inclusion/exclusion criteria of healthy subjects in two samples were identical to those of the patient group except that they did not meet the DSM-IV diagnostic criteria of any mental disorders screened using the SCID non-patient version. We also ensured that for healthy subjects, no first-degree relatives had a history of any psychiatric disorders. | | TE=4.2 ms, flip angle=15°, 172 slices, matrix size=256×256, and the field of view (FOV)=24×24 cm2. |
| CN-Xian | DSM-IV, DSM-V | Inclusion criteria were the following: (1) diagnosis of schizophrenia using the Structured Clinical Interview for the Diagnostic and Statistical Manual of Mental Disorders, Fourth Edition (SCID-IV), (2) patients were taking stable doses of antipsychotic medication for at least 8 weeks before the study; (3) no history of significant head trauma or neurological disorders and no focal brain lesions by T1- or T2-weighted MRI; (4) no alcohol or drug abuse; and (5) patient has no recent aggression or other forms of behavioral dysfunction. | GE Discovery MR750 3.0 T | High-resolution T1-weighted MRI was acquired using a GE Discovery MR750 3.0 T scanner located in the Department of Radiology, and all subjects underwent T2WI scans to rule out organic diseases. The scanning parameters were repetition time 8.2 ms, echo time 3.2 ms, flip angle 12°, field of view 256 × 256 mm, matrix 256 × 256, slice thickness 1 mm, and sagittal slices 196. |
| HCP-EP | DSM-V | Available at https://www.humanconnectome.org/study/human-connectome-project-for-early-psychosis | | |
| JP-SRPBS | | Available at https://bicr-resource.atr.jp/srpbsfc/ | | |
| fBIRN | SCID DSM-IV-TR | All subjects diagnosed with schizophrenia were clinically stable outpatients whose antipsychotic medications and doses had not changed within the last two months. Schizophrenia and healthy volunteers with a history of major medical illness, drug dependence in the last five years (except for nicotine), current substance abuse disorder, or MRI contraindications, were excluded. Individuals with schizophrenia who had significant tardive dyskinesia and healthy volunteers with a current or past history of major neurological or psychiatric illness or with a first-degree relative with a DSM IV Axis-I psychotic disorder diagnosis were also excluded. | 3T Siemens Tim Trio; 3T GE | High-resolution structural imaging scans were acquired on six 3T Siemens Tim® Trio System and one 3T General Electric Discovery MR750 scanner. MP-RAGE scan parameters for the Siemens scanner were: scan plane=sagittal, TR/TE/TI=2300/2.94/1100ms, GRAPPA acceleration factor=2, flip angle=9°, resolution=256×256x160, FOV=220mm2, voxel size=0.86x0.86x1.2mm, and NEX=1. IR-SPGR scan parameters for the General Electric scanner were: scan plane=sagittal, TR/TE/TI=5.95/1.99/450ms, ASSET acceleration factor=2, a flip angle=12°, resolution=256×256x166, FOV=220mm2, voxel size=0.86x0.86x1.2mm, and NEX=1. All scans covered the entire brain. |
| MCIC | SCID DSM-IV (SCID-NP for controls) or CASH were used to diagnose primary and co-morbid psychiatric disorders in | All subjects were between the ages of 18 and 60 and spoke English as their native language. To be included in the schizophrenia cohort, patients had to meet diagnostic criteria for schizophrenia, schizoaffective disorder, or schizophreniform disorder. Concerted effort was made to recruit patients early in the course of their illness and especially those who were antipsychotic drug naïve. The healthy control subjects with no current or past history of psychiatric illness including substance abuse or dependence were matched within site to the patient cohort for age, sex, and parental education. Control subjects who had not been diagnosed with any psychiatric disorders, but had been medicated with antidepressants, anti-anxiety medication or medication for sleep disturbance | 1.5, 3T Siemens and GE | T1 scans: TR = 2530 ms for 3 T, TR = 12 ms for 1.5 T; TE = 3.79 ms for 3 T, TE = 4.76 ms for 1.5 T; FA = 7 for 3 T, FA = 20 for 1.5 T; TI = 1100 for 3 T; Bandwidth = 181 for 3 T, Bandwidth = 110 for 1.5 T; 0.625×0.625 mm voxel size; slice thickness 1.5 mm; FOV 256×256×128 cm matrix; FOV = 16 cm (could be increased to 18 cm when needed for full brain coverage). |

| | | | | |
|---|---|---|---|---|
| | controls and patients | were included in the study provided that the duration of their medication did not exceed 2 months of lifetime use and no medication was used within the 6 months preceding the baseline MRI scan. Control subjects who met criteria for current or past history of substance abuse or dependence were excluded from the study. Patients, however, were not excluded from the study unless criteria were met for current (i.e., within the past month) abuse or dependence (except for 6 patients who were found to meet criteria for current abuse after the study data was collected). Both patients and controls were excluded if they had (1) an IQ less than 70 based on a standardized IQ test, (2) history of a head injury resulting in prolonged loss of consciousness, neurosurgical procedure, neurological disease, history of skull fracture, severe or disabling medical conditions, or (3) a contraindication for MRI scanning such as pregnancy, metal in body or head including implanted pacemaker, medication pump, vagal stimulator, deep brain stimulator, implanted TENS unit, or ventriculo-peritoneal shunt | | |
| NMorphCH | This is a longitudinal study examining the clinical, cognitive and neuroimaging (MRI) data from schizophrenia and control subjects at baseline and after two years. The study was conducted at Northwestern University. Neuroimaging data includes T1, T2, DTI, resting-state fMRI and n-back task fMRI. More details are provide at http://www.schizconnect.org/documentation | | 3T | MPRAGE:voxel size =1x1x1.6 mm3; Matrix size 256x256; TR=3.15ms; TE=20ms; Flip ange=8.0 |
| NUSDAST | The Northwestern University Schizophrenia Data and Software Tool (NUSDAST) is a repository of schizophrenia neuroimaging data collected from over 450 individuals with schizophrenia, healthy controls and their respective siblings, most with 2-year longitudinal follow-up. More details are provided at: Wang L, Kogan A, Cobia D, et al. Northwestern University schizophrenia data and software tool (NUSDAST). Frontiers in neuroinformatics, 2013, 7: 25. | | Siemens 1.5 T Vision scanner | 3D MPRAGE (TR = 9.7 ms, TE = 4 ms, flip = 10°, ACQ = 1, 256 × 256 matrix, 1 × l mm in-plane resolution, 128 slices, slice thickness 1.25 mm) |
| DS000030 | DSM-IV | For both healthy and patient groups, participants were men or women ages 21–50 years; NIH racial/ethnic category either White, not Hispanic or Latino; or Hispanic or Latino, of any racial group; primary language either English or Spanish; completed at least 8 years of formal education; no significant medical illness; adequate cooperation to complete assessments; visual acuity 20/60 or better; and urinalysis negative for drugs of abuse (Cocaine; Methamphetamine; Morphine; THC; and Benzodiazepines).<br>Participants in the healthy group were excluded if they had lifetime diagnoses of Schizophrenia or Other Psychotic Disorder, Bipolar I or II Disorder, or Substance Abuse or Dependence (not counting caffeine or nicotine); or current Major Depressive Disorder; suicidality; Anxiety Disorder (Obsessive Compulsive Disorder, Panic Disorder, Generalized Anxiety Disorder, Post-Traumatic Stress Disorder), Attention Deficit Hyperactivity Disorder (ADHD). ADHD criteria were assessed using Adult ADHD Interview; healthy participants were screened for sub-threshold ADHD, defined as 4 or more ADHD inattentive or hyperactive/impulsive symptoms in either childhood and adulthood; in addition, they could not have had medication treatment for ADHD within the prior 12 | Siemens Trio (2 Imaging Sites) | A T1-weighted high-resolution anatomical scan (MPRAGE) were collected with the following parameter: slice thickness = 1mm, 176 slices, TR=1.9s, TE=2.26ms, matrix=256 x 256, FOV=250mm. |

| | | | | |
|---|---|---|---|---|
| | | months. Each of the patient groups (Schizophrenia, Bipolar Disorder, and ADHD) excluded anyone with one of these other diagnoses; stable medications were permitted for the patients. For MRI studies we excluded participants who were left handed, who believed they might be pregnant, or had other contraindications to scanning (e.g., claustrophobia, metal in body, body too large to fit in scanner).<br>More details are provided at https://www.nature.com/articles/sdata2016110 | | |
| DS000115 | DSM-IV | All subjects were diagnosed on the basis of a consensus between a research psychiatrist who conducted a semi-structured interview and a trained research assistant who used the Structured Clinical Interview for DSM-IV Axis I Disorders. Participants were excluded if they: (a) met DSM-IV criteria for substance dependence or severe/moderate abuse during the prior 6 months; (b) had a clinically unstable or severe medical disorder; (c) had a history of head injury with documented neurological sequelae or loss of consciousness; or (d) met DSM-IV criteria for mental retardation. The individuals with schizophrenia were all outpatients, and were stabilized on antipsychotic medication for at least 2 weeks. Controls were required to have no lifetime history of Axis I psychotic or mood disorders and no first-degree relatives with a psychotic disorder. | 3T Siemens Trio | T1 structural image was acquired using a sagittal MP-RAGE 3D sequence (TR = 2400 ms, TE = 3.16 ms, flip = 8°; voxel size = 1 mm × 1 mm × 1 mm). |
| DS004302 | DSM-V | Patients meeting DSM-5 criteria for schizophrenia were initially recruited from four psychiatric hospitals in Barcelona. Diagnoses were made using the Structured Clinical Interview for DSM Disorders (SCID). Patients were excluded if they (a) were younger than 18 or older than 65, (b) had a history of brain trauma or neurological disease or (c) had shown alcohol/substance abuse/dependence within 12 months prior to participation. Social use of alcohol was permitted, as was non-habitual use of cannabis. Electroconvulsive therapy in the past 6 months was also an exclusion criterion. All participants were right-handed and were taking antipsychotic medication. Healthy controls met the same exclusion criteria as the patients, and they were also interviewed using the SCID to exclude current and past psychiatric disorders. They were questioned and excluded if they reported a history of treatment with psychotropic medication beyond non-habitual use of night sedation. Controls were also excluded if they reported a history of major psychiatric disorder in a first-degree relative. | 3T Philips Ingenia scanner | Images were acquired with a 3T Philips Ingenia scanner (Philips Medical Systems, Best, The Netherlands). High-resolution anatomical volume with an FFE (Fast Field Echo) sequence for anatomical reference and inspection (TR = 9.90ms; TE = 4.60ms; Flip angle = 8°; voxel size = 1 × 1mm; slice thickness = 1mm; slice number = 180; FOV = 240mm). |

**Supplementary Table S3. Demographic and clinical characteristics of participants in the first-episode subsample and medicationn-naive subsample.**

| | First-episode cohort (n=1122) | | Medication-naïve cohort (n=718) | |
|---|---|---|---|---|
| | n | mean(SD) | n | mean(SD) |
| Sex (Female/Male) | 513/609 | - | 353/365 | - |
| Age (years) | 1122 | 25.4(8.6) | 718 | 23.7(7.8) |
| Illness duration (years) | 710 | 0.78(0.67) | 185 | 1.3(2.2) |
| PANSS Positive scale (P1-P7) | 926 | 19.5(6.4) | 605 | 21.1(5.5) |
| PANSS Negative scale (N1-N7) | 926 | 17.0(7.3) | 605 | 18.5(7.8) |
| PANSS General scale (G1-G16) | 926 | 37.6(10.3) | 605 | 39.6(8.1) |
| PANSS Total score | 926 | 74.1(20.2) | 605 | 79.2(16.6) |
| PANSS excitement dimension (P4, P7, G44, G14) | 597 | 8.8(3.4) | 533 | 8.7(3.3) |
| PANSS depression/anxiety dimension (G1, G2, G3, G6, G15) | 597 | 11.3(3.8) | 533 | 11.0(3.6) |
| PANSS cognitive dimension (P2, N5, G5, G10, G11) | 597 | 10.1(3.7) | 533 | 10.0(3.7) |

**Supplementary Table S4. Demographic and clinical characteristics of participants in six cohorts by their locations.**

| | East Asian ancestry cohort (n=2235) | | European ancestry cohort (n=1987) | | China cohort (n=1542) | | Japan cohort (n=438) | | Europe cohort (n=1044) | | North American cohort (n=729) | |
|---|---|---|---|---|---|---|---|---|---|---|---|---|
| | n | mean(SD) | n | mean(SD) | n | mean(SD) | n | mean(SD) | n | mean(SD) | n | mean(SD) |
| Sex (Female/Male) | 1017/1218 | - | 666/1321 | - | 718/824 | - | 195/243 | - | 374/670 | - | 207/522 | - |
| Age (years) | 2235 | 29.8(11.5) | 1987 | 35.3(11.6) | 1542 | 27.4(10.5) | 438 | 36.0(12.1) | 1044 | 35.1(11.1) | 729 | 34.4(12.4) |
| Illness duration (years) | 1228 | 7.7(9.1) | 1105 | 13.5(11.0) | 587 | 5.7(8.6) | 386 | 11.9(9.4) | 580 | 13.3(10.7) | 359 | 13.6(12.0) |
| PANSS Positive scale (P1-P7) | 1847 | 17.8(6.9) | 804 | 15.8(6.3) | 1180 | 18.9(6.9) | 413 | 18.2(6.1) | 448 | 17.4(6.5) | 152 | 14.0(4.9) |
| PANSS Negative scale (N1-N7) | 1847 | 17.1(7.8) | 804 | 18.3(7.2) | 1180 | 17.6(8.0) | 413 | 19.6(6.2) | 448 | 19.8(7.3) | 152 | 15.7(5.6) |
| PANSS General scale (G1-G16) | 1847 | 35.2(11.6) | 804 | 34.0(11.6) | 1180 | 35.9(10.4) | 413 | 40.6(12.1) | 448 | 37.9(12.3) | 152 | 27.7(7.8) |
| PANSS Total score | 1847 | 70.1(22.9) | 804 | 68.0(21.2) | 1180 | 72.4(21.0) | 413 | 78.4(22.6) | 448 | 75.1(22.2) | 152 | 57.4(13.9) |
| PANSS excitement dimension (P4, P7, G44, G14) | 770 | 8.7(3.4) | 552 | 7.5(3.4) | 691 | 9.0(3.4) | 79 | 6.1(2.1) | 406 | 8.2(3.5) | 79 | 4.9(1.4) |
| PANSS depression/anxiety dimension (G1, G2, G3, G6, G15) | 770 | 11.0(3.5) | 552 | 11.6(4.9) | 691 | 11.2(3.5) | 79 | 9.7(3.2) | 406 | 12.3(4.8) | 79 | 8.2(3.0) |
| PANSS cognitive dimension (P2, N5, G5, G10, G11) | 770 | 10.2(3.7) | 552 | 11.1(4.3) | 691 | 10.3(3.7) | 79 | 9.0(3.2) | 406 | 11.7(4.6) | 79 | 9.7(2.8) |
| Cohorts included | IMH; JBUN; Osaka; CN-Shanghai1; CN-Shanghai2; CN-Shanghai3; CN-Harbin; CN-Chengdu; CN-Taibei; CN-Zhengzhou; CN-Beijing1; CN-Beijing2; CN-Changsha; CN-Xian; JP-SRPBS | | ASRB; ESO; FIDMAG; FOR2107-MR; FOR2017-MS; OLIN; PENS; PHCP; RomeSL; SoCAT; SWIFT; UCISZ; UNINA; Zurich; COBRE; TOPSY; Voices; OSLO_TOP; HCP-EP; fBIRN; MCIC; NMorphCH; NUSDAST; DS000030; DS000115; DS004302 | | CN-Shanghai1; CN-Shanghai2; CN-Shanghai3; CN-Harbin; CN-Chengdu; CN-Taibei; CN-Zhengzhou; CN-Beijing1; CN-Beijing2; CN-Changsha; CN-Xian | | Osaka; JP-SRPBS | | ESO; FIDMAG; FOR2107-MR; FOR2017-MS; RomeSL; SWIFT; UNINA; Zurich; OSLO_TOP; DS004302 | | OLIN; PENS; PHCP; UCISZ; COBRE; TOPSY; HCP-EP; fBIRN; MCIC; NMorphCH; NUSDAST; DS000030; DS000115 | |

**Supplementary Table S5. Morphological z-scores of all brain regions and inter-subtype comparisons.**

| Number | Region | Morphological measure (Freesurfer) | Subtype1 (n=1,131) | | Subtype2 (n=709) | | Subtype1 vs. Subtype2 | | | |
|---|---|---|---|---|---|---|---|---|---|---|
| | | | Mean | SD | Mean | SD | Cohen's d | T | P | FWE |
| 1 | Left_Lateral_Ventricle | Subcortical volume | -0.186 | 0.992 | -0.459 | 1.355 | 0.230 | 4.971 | 7.297E-07 | * |
| 2 | Left_Thalamus | Subcortical volume | 0.196 | 1.123 | 0.449 | 1.090 | -0.228 | -4.745 | 2.251E-06 | * |
| 3 | Left_Caudate | Subcortical volume | -0.264 | 1.034 | 0.279 | 1.073 | -0.516 | -10.815 | 0.000E+00 | * |
| 4 | Left_Putamen | Subcortical volume | -0.204 | 1.023 | 0.213 | 0.990 | -0.415 | -8.623 | 0.000E+00 | * |
| 5 | Left_Pallidum | Subcortical volume | -0.273 | 0.987 | 0.083 | 0.931 | -0.371 | -7.699 | 2.232E-14 | * |
| 6 | Left_Hippocampus | Subcortical volume | 0.169 | 0.988 | 0.586 | 0.821 | -0.460 | -9.393 | 0.000E+00 | * |
| 7 | Left_Amygdala | Subcortical volume | 0.096 | 0.968 | 0.399 | 0.897 | -0.325 | -6.724 | 2.358E-11 | * |
| 8 | Left_Accumbens_area | Subcortical volume | 0.080 | 1.040 | 0.383 | 1.051 | -0.290 | -6.056 | 1.691E-09 | * |
| 9 | Right_Lateral_Ventricle | Subcortical volume | -0.206 | 1.143 | -0.429 | 1.317 | 0.181 | 3.835 | 1.297E-04 | * |
| 10 | Right_Thalamus | Subcortical volume | 0.224 | 1.132 | 0.457 | 1.147 | -0.205 | -4.279 | 1.970E-05 | * |
| 11 | Right_Caudate | Subcortical volume | -0.268 | 1.061 | 0.240 | 1.109 | -0.468 | -9.820 | 0.000E+00 | * |
| 12 | Right_Putamen | Subcortical volume | -0.211 | 0.983 | 0.181 | 1.040 | -0.388 | -8.144 | 6.661E-16 | * |
| 13 | Right_Pallidum | Subcortical volume | -0.291 | 0.985 | 0.051 | 0.987 | -0.347 | -7.245 | 6.323E-13 | * |
| 14 | Right_Hippocampus | Subcortical volume | 0.152 | 1.041 | 0.594 | 0.918 | -0.449 | -9.246 | 0.000E+00 | * |
| 15 | Right_Amygdala | Subcortical volume | 0.005 | 1.004 | 0.370 | 0.920 | -0.379 | -7.825 | 8.438E-15 | * |
| 16 | Right_Accumbens_area | Subcortical volume | 0.083 | 0.937 | 0.377 | 0.989 | -0.305 | -6.402 | 1.941E-10 | * |
| 17 | Left_Lateral_nucleus | Amygdala segmentation volume | 0.029 | 0.919 | 0.374 | 0.822 | -0.395 | -8.144 | 6.661E-16 | * |
| 18 | Left_Basal_nucleus | Amygdala segmentation volume | 0.116 | 0.952 | 0.451 | 0.829 | -0.375 | -7.713 | 1.998E-14 | * |
| 19 | Left_Accessory_Basal_nucleus | Amygdala segmentation volume | 0.144 | 0.964 | 0.488 | 0.861 | -0.376 | -7.753 | 1.477E-14 | * |
| 20 | Left_Anterior_amygdaloid_area_AAA | Amygdala segmentation volume | 0.142 | 0.985 | 0.352 | 0.887 | -0.224 | -4.620 | 4.110E-06 | * |
| 21 | Left_Central_nucleus | Amygdala segmentation volume | -0.002 | 0.932 | 0.179 | 0.919 | -0.195 | -4.065 | 5.014E-05 | * |
| 22 | Left_Medial_nucleus | Amygdala segmentation volume | -0.018 | 0.943 | 0.138 | 0.954 | -0.165 | -3.437 | 6.009E-04 | |
| 23 | Left_Cortical_nucleus | Amygdala segmentation volume | 0.029 | 0.981 | 0.322 | 0.963 | -0.301 | -6.274 | 4.382E-10 | * |
| 24 | Left_Corticoamygdaloid_transitio | Amygdala segmentation volume | 0.198 | 0.962 | 0.536 | 0.843 | -0.374 | -7.685 | 2.465E-14 | * |
| 25 | Left_Paralaminar_nucleus | Amygdala segmentation volume | 0.047 | 0.979 | 0.388 | 0.848 | -0.372 | -7.644 | 3.375E-14 | * |
| 26 | Right_Lateral_nucleus | Amygdala segmentation volume | 0.007 | 0.964 | 0.402 | 0.898 | -0.425 | -8.792 | 0.000E+00 | * |
| 27 | Right_Basal_nucleus | Amygdala segmentation volume | 0.074 | 1.020 | 0.491 | 0.938 | -0.426 | -8.812 | 0.000E+00 | * |
| 28 | Right_Accessory_Basal_nucleus | Amygdala segmentation volume | 0.122 | 1.001 | 0.500 | 0.953 | -0.387 | -8.027 | 1.776E-15 | * |
| 29 | Right_Anterior_amygdaloid_area_AAA | Amygdala segmentation volume | 0.106 | 0.976 | 0.337 | 0.952 | -0.239 | -4.980 | 6.955E-07 | * |
| 30 | Right_Central_nucleus | Amygdala segmentation volume | -0.008 | 0.972 | 0.207 | 0.970 | -0.222 | -4.629 | 3.931E-06 | * |

| | | | | | | | | | | |
|---|---|---|---|---|---|---|---|---|---|---|
| 31 | Right_Medial_nucleus | Amygdala segmentation volume | -0.047 | 0.989 | 0.146 | 0.998 | -0.194 | -4.059 | 5.134E-05 | * |
| 32 | Right_Cortical_nucleus | Amygdala segmentation volume | 0.017 | 1.001 | 0.315 | 1.011 | -0.296 | -6.192 | 7.311E-10 | * |
| 33 | Right_Corticoamygdaloid_transitio | Amygdala segmentation volume | 0.174 | 1.023 | 0.559 | 0.961 | -0.388 | -8.038 | 1.665E-15 | * |
| 34 | Right_Paralaminar_nucleus | Amygdala segmentation volume | 0.010 | 1.013 | 0.423 | 0.924 | -0.426 | -8.789 | 0.000E+00 | * |
| 35 | Left_Hippocampal_tail | Hippocampus segmentation volume | 0.149 | 0.989 | 0.442 | 0.973 | -0.299 | -6.217 | 6.244E-10 | * |
| 36 | Left_subiculum_body | Hippocampus segmentation volume | 0.050 | 0.979 | 0.403 | 0.900 | -0.376 | -7.761 | 1.388E-14 | * |
| 37 | Left_CA1_body | Hippocampus segmentation volume | 0.129 | 1.012 | 0.256 | 0.999 | -0.126 | -2.633 | 8.538E-03 | |
| 38 | Left_subiculum_head | Hippocampus segmentation volume | 0.023 | 0.989 | 0.312 | 0.881 | -0.309 | -6.361 | 2.517E-10 | * |
| 39 | Left_hippocampal_fissure | Hippocampus segmentation volume | -0.169 | 0.975 | 0.011 | 0.991 | -0.183 | -3.829 | 1.332E-04 | * |
| 40 | Left_presubiculum_head | Hippocampus segmentation volume | 0.072 | 0.984 | 0.372 | 0.900 | -0.319 | -6.585 | 5.916E-11 | * |
| 41 | Left_CA1_head | Hippocampus segmentation volume | 0.125 | 1.000 | 0.451 | 0.842 | -0.353 | -7.226 | 7.249E-13 | * |
| 42 | Left_presubiculum_body | Hippocampus segmentation volume | 0.095 | 0.965 | 0.353 | 0.948 | -0.269 | -5.608 | 2.359E-08 | * |
| 43 | Left_parasubiculum | Hippocampus segmentation volume | 0.123 | 1.014 | 0.209 | 1.029 | -0.084 | -1.756 | 7.933E-02 | |
| 44 | Left_molecular_layer_HP_head | Hippocampus segmentation volume | 0.145 | 0.995 | 0.471 | 0.833 | -0.355 | -7.266 | 5.458E-13 | * |
| 45 | Left_molecular_layer_HP_body | Hippocampus segmentation volume | 0.198 | 0.980 | 0.513 | 0.892 | -0.336 | -6.929 | 5.851E-12 | * |
| 46 | Left_GC_ML_DG_head | Hippocampus segmentation volume | 0.189 | 1.022 | 0.446 | 0.859 | -0.273 | -5.587 | 2.657E-08 | * |
| 47 | Left_CA3_body | Hippocampus segmentation volume | 0.066 | 0.979 | 0.208 | 0.994 | -0.144 | -3.000 | 2.735E-03 | |
| 48 | Left_GC_ML_DG_body | Hippocampus segmentation volume | 0.188 | 0.987 | 0.438 | 0.929 | -0.261 | -5.399 | 7.551E-08 | * |
| 49 | Left_CA4_head | Hippocampus segmentation volume | 0.165 | 1.023 | 0.404 | 0.876 | -0.251 | -5.149 | 2.898E-07 | * |
| 50 | Left_CA4_body | Hippocampus segmentation volume | 0.182 | 0.967 | 0.425 | 0.913 | -0.258 | -5.348 | 9.993E-08 | * |
| 51 | Left_fimbria | Hippocampus segmentation volume | 0.049 | 1.119 | 0.265 | 1.053 | -0.199 | -4.122 | 3.918E-05 | * |
| 52 | Left_CA3_head | Hippocampus segmentation volume | 0.118 | 1.003 | 0.298 | 0.923 | -0.187 | -3.868 | 1.137E-04 | * |
| 53 | Left_HATA | Hippocampus segmentation volume | 0.200 | 0.978 | 0.452 | 0.847 | -0.276 | -5.658 | 1.771E-08 | * |
| 54 | Right_Hippocampal_tail | Hippocampus segmentation volume | 0.111 | 1.001 | 0.461 | 0.968 | -0.355 | -7.380 | 2.383E-13 | * |
| 55 | Right_subiculum_body | Hippocampus segmentation volume | 0.046 | 0.974 | 0.400 | 0.907 | -0.376 | -7.791 | 1.099E-14 | * |
| 56 | Right_CA1_body | Hippocampus segmentation volume | 0.064 | 1.037 | 0.231 | 0.982 | -0.165 | -3.420 | 6.398E-04 | |
| 57 | Right_subiculum_head | Hippocampus segmentation volume | 0.007 | 1.074 | 0.327 | 0.974 | -0.312 | -6.435 | 1.574E-10 | * |
| 58 | Right_hippocampal_fissure | Hippocampus segmentation volume | -0.285 | 1.071 | -0.081 | 1.050 | -0.193 | -4.013 | 6.240E-05 | * |
| 59 | Right_presubiculum_head | Hippocampus segmentation volume | 0.096 | 1.051 | 0.409 | 1.004 | -0.304 | -6.311 | 3.461E-10 | * |
| 60 | Right_CA1_head | Hippocampus segmentation volume | 0.105 | 1.088 | 0.466 | 0.948 | -0.354 | -7.278 | 4.993E-13 | * |
| 61 | Right_presubiculum_body | Hippocampus segmentation volume | 0.099 | 1.037 | 0.392 | 1.013 | -0.285 | -5.935 | 3.495E-09 | * |
| 62 | Right_parasubiculum | Hippocampus segmentation volume | 0.143 | 1.029 | 0.304 | 1.034 | -0.155 | -3.245 | 1.195E-03 | |
| 63 | Right_molecular_layer_HP_head | Hippocampus segmentation volume | 0.133 | 1.083 | 0.492 | 0.946 | -0.354 | -7.275 | 5.101E-13 | * |

| | | | | | | | | | | |
|---|---|---|---|---|---|---|---|---|---|---|
| 64 | Right_molecular_layer_HP_body | Hippocampus segmentation volume | 0.150 | 1.035 | 0.495 | 0.980 | -0.343 | -7.106 | 1.707E-12 | * |
| 65 | Right_GC_ML_DG_head | Hippocampus segmentation volume | 0.164 | 1.045 | 0.427 | 0.918 | -0.268 | -5.507 | 4.160E-08 | * |
| 66 | Right_CA3_body | Hippocampus segmentation volume | 0.010 | 0.972 | 0.164 | 0.946 | -0.160 | -3.330 | 8.841E-04 | |
| 67 | Right_GC_ML_DG_body | Hippocampus segmentation volume | 0.134 | 1.041 | 0.405 | 1.016 | -0.263 | -5.479 | 4.856E-08 | * |
| 68 | Right_CA4_head | Hippocampus segmentation volume | 0.144 | 1.023 | 0.376 | 0.917 | -0.239 | -4.917 | 9.574E-07 | * |
| 69 | Right_CA4_body | Hippocampus segmentation volume | 0.121 | 1.018 | 0.358 | 1.003 | -0.235 | -4.894 | 1.073E-06 | * |
| 70 | Right_fimbria | Hippocampus segmentation volume | 0.076 | 1.061 | 0.240 | 0.996 | -0.159 | -3.303 | 9.763E-04 | |
| 71 | Right_CA3_head | Hippocampus segmentation volume | 0.108 | 0.978 | 0.271 | 0.897 | -0.174 | -3.586 | 3.439E-04 | |
| 72 | Right_HATA | Hippocampus segmentation volume | 0.192 | 1.018 | 0.489 | 0.885 | -0.311 | -6.396 | 2.017E-10 | * |
| 73 | Left_AV | Thalamus segmentation volume | 0.063 | 1.074 | 0.249 | 1.014 | -0.179 | -3.704 | 2.188E-04 | |
| 74 | Left_CeM | Thalamus segmentation volume | 0.029 | 1.080 | 0.334 | 1.067 | -0.284 | -5.923 | 3.773E-09 | * |
| 75 | Left_CL | Thalamus segmentation volume | 0.025 | 0.927 | 0.137 | 0.868 | -0.124 | -2.566 | 1.036E-02 | |
| 76 | Left_CM | Thalamus segmentation volume | 0.009 | 1.032 | 0.287 | 0.930 | -0.283 | -5.835 | 6.341E-09 | * |
| 77 | Left_LD | Thalamus segmentation volume | 0.161 | 0.920 | 0.286 | 0.934 | -0.135 | -2.813 | 4.954E-03 | |
| 78 | Left_LGN | Thalamus segmentation volume | 0.083 | 1.029 | 0.315 | 0.999 | -0.229 | -4.759 | 2.094E-06 | * |
| 79 | Left_LP | Thalamus segmentation volume | 0.136 | 0.990 | 0.237 | 0.984 | -0.102 | -2.132 | 3.312E-02 | |
| 80 | Left_L_Sg | Thalamus segmentation volume | -0.026 | 1.005 | 0.109 | 0.947 | -0.138 | -2.863 | 4.248E-03 | |
| 81 | Left_MDl | Thalamus segmentation volume | 0.187 | 1.024 | 0.314 | 1.163 | -0.116 | -2.448 | 1.444E-02 | |
| 82 | Left_MDm | Thalamus segmentation volume | 0.247 | 1.063 | 0.430 | 1.158 | -0.165 | -3.467 | 5.380E-04 | |
| 83 | Left_MGN | Thalamus segmentation volume | 0.040 | 1.121 | 0.250 | 1.027 | -0.195 | -4.031 | 5.788E-05 | * |
| 84 | Left_MV_Re_ | Thalamus segmentation volume | 0.066 | 0.990 | 0.376 | 1.018 | -0.308 | -6.446 | 1.463E-10 | * |
| 85 | Left_Pc | Thalamus segmentation volume | 0.099 | 1.068 | 0.283 | 1.063 | -0.173 | -3.612 | 3.117E-04 | |
| 86 | Left_Pf | Thalamus segmentation volume | 0.058 | 0.954 | 0.201 | 0.842 | -0.159 | -3.264 | 1.118E-03 | |
| 87 | Left_Pt | Thalamus segmentation volume | 0.077 | 1.079 | 0.269 | 0.927 | -0.191 | -3.920 | 9.163E-05 | * |
| 88 | Left_PuA | Thalamus segmentation volume | 0.137 | 1.107 | 0.387 | 1.111 | -0.225 | -4.704 | 2.737E-06 | * |
| 89 | Left_PuI | Thalamus segmentation volume | -0.034 | 1.048 | 0.246 | 1.022 | -0.271 | -5.628 | 2.099E-08 | * |
| 90 | Left_PuL | Thalamus segmentation volume | -0.062 | 1.120 | 0.012 | 1.177 | -0.064 | -1.350 | 1.770E-01 | |
| 91 | Left_PuM | Thalamus segmentation volume | 0.057 | 1.108 | 0.360 | 1.097 | -0.275 | -5.736 | 1.133E-08 | * |
| 92 | Left_VA | Thalamus segmentation volume | 0.049 | 1.001 | 0.223 | 1.023 | -0.171 | -3.587 | 3.432E-04 | |
| 93 | Left_VAmc | Thalamus segmentation volume | -0.020 | 1.011 | 0.234 | 0.976 | -0.256 | -5.316 | 1.193E-07 | * |
| 94 | Left_VLa | Thalamus segmentation volume | 0.114 | 1.038 | 0.238 | 1.008 | -0.121 | -2.514 | 1.202E-02 | |
| 95 | Left_VLp | Thalamus segmentation volume | 0.118 | 1.046 | 0.268 | 0.992 | -0.147 | -3.043 | 2.379E-03 | |
| 96 | Left_VM | Thalamus segmentation volume | 0.085 | 0.967 | 0.254 | 0.928 | -0.178 | -3.698 | 2.237E-04 | |

| | | | | | | | | | | |
|---|---|---|---|---|---|---|---|---|---|---|
| 97 | Left_VPL | Thalamus segmentation volume | 0.069 | 1.030 | 0.253 | 1.008 | -0.180 | -3.751 | 1.812E-04 | |
| 98 | Right_AV | Thalamus segmentation volume | 0.124 | 1.042 | 0.235 | 1.050 | -0.106 | -2.211 | 2.719E-02 | |
| 99 | Right_CeM | Thalamus segmentation volume | 0.046 | 1.106 | 0.313 | 1.083 | -0.243 | -5.068 | 4.434E-07 | * |
| 100 | Right_CL | Thalamus segmentation volume | 0.038 | 0.975 | 0.137 | 0.930 | -0.103 | -2.139 | 3.256E-02 | |
| 101 | Right_CM | Thalamus segmentation volume | 0.049 | 1.018 | 0.294 | 0.925 | -0.253 | -5.219 | 2.000E-07 | * |
| 102 | Right_LD | Thalamus segmentation volume | 0.211 | 0.938 | 0.327 | 0.926 | -0.125 | -2.596 | 9.516E-03 | |
| 103 | Right_LGN | Thalamus segmentation volume | 0.134 | 1.133 | 0.308 | 1.092 | -0.157 | -3.261 | 1.130E-03 | |
| 104 | Right_LP | Thalamus segmentation volume | 0.153 | 1.031 | 0.270 | 1.051 | -0.113 | -2.353 | 1.871E-02 | |
| 105 | Right_L_Sg | Thalamus segmentation volume | -0.051 | 1.018 | 0.142 | 0.955 | -0.195 | -4.046 | 5.432E-05 | * |
| 106 | Right_MDl | Thalamus segmentation volume | 0.236 | 1.046 | 0.302 | 1.131 | -0.060 | -1.267 | 2.053E-01 | |
| 107 | Right_MDm | Thalamus segmentation volume | 0.284 | 1.071 | 0.417 | 1.172 | -0.118 | -2.498 | 1.257E-02 | |
| 108 | Right_MGN | Thalamus segmentation volume | 0.069 | 1.115 | 0.265 | 1.045 | -0.181 | -3.751 | 1.813E-04 | |
| 109 | Right_MV_Re_ | Thalamus segmentation volume | 0.085 | 1.087 | 0.373 | 1.047 | -0.270 | -5.608 | 2.362E-08 | * |
| 110 | Right_Pc | Thalamus segmentation volume | 0.129 | 1.028 | 0.339 | 1.052 | -0.202 | -4.221 | 2.552E-05 | * |
| 111 | Right_Pf | Thalamus segmentation volume | 0.038 | 0.958 | 0.189 | 0.878 | -0.164 | -3.382 | 7.345E-04 | |
| 112 | Right_Pt | Thalamus segmentation volume | 0.143 | 1.041 | 0.339 | 0.953 | -0.196 | -4.055 | 5.228E-05 | * |
| 113 | Right_PuA | Thalamus segmentation volume | 0.196 | 1.039 | 0.338 | 1.038 | -0.137 | -2.861 | 4.277E-03 | |
| 114 | Right_PuI | Thalamus segmentation volume | 0.025 | 1.013 | 0.196 | 0.985 | -0.171 | -3.567 | 3.709E-04 | |
| 115 | Right_PuL | Thalamus segmentation volume | -0.025 | 1.069 | 0.026 | 1.005 | -0.048 | -1.003 | 3.162E-01 | |
| 116 | Right_PuM | Thalamus segmentation volume | 0.128 | 1.027 | 0.319 | 1.020 | -0.186 | -3.887 | 1.052E-04 | * |
| 117 | Right_VA | Thalamus segmentation volume | 0.060 | 1.042 | 0.231 | 1.035 | -0.165 | -3.438 | 5.987E-04 | |
| 118 | Right_VAmc | Thalamus segmentation volume | 0.000 | 1.040 | 0.255 | 0.973 | -0.253 | -5.233 | 1.859E-07 | * |
| 119 | Right_VLa | Thalamus segmentation volume | 0.149 | 1.063 | 0.262 | 1.065 | -0.107 | -2.225 | 2.620E-02 | |
| 120 | Right_VLp | Thalamus segmentation volume | 0.160 | 1.066 | 0.271 | 1.066 | -0.104 | -2.173 | 2.987E-02 | |
| 121 | Right_VM | Thalamus segmentation volume | 0.107 | 1.011 | 0.236 | 0.992 | -0.129 | -2.691 | 7.187E-03 | |
| 122 | Right_VPL | Thalamus segmentation volume | 0.115 | 0.995 | 0.239 | 1.029 | -0.123 | -2.570 | 1.025E-02 | |
| 123 | Medulla | Brain Stem segmentation volume | -0.108 | 0.952 | 0.122 | 0.917 | -0.246 | -5.111 | 3.529E-07 | * |
| 124 | Pons | Brain Stem segmentation volume | -0.059 | 0.950 | 0.393 | 0.909 | -0.486 | -10.095 | 0.000E+00 | * |
| 125 | SCP | Brain Stem segmentation volume | -0.030 | 1.010 | 0.233 | 0.954 | -0.268 | -5.546 | 3.353E-08 | * |
| 126 | Midbrain | Brain Stem segmentation volume | -0.092 | 1.009 | 0.371 | 0.973 | -0.467 | -9.701 | 0.000E+00 | * |
| 127 | Left_bankssts_volume | Cortical volume | 0.169 | 0.979 | 0.080 | 1.028 | 0.089 | 1.866 | 6.225E-02 | |
| 128 | Left_caudalanteriorcingulate_volume | Cortical volume | 0.146 | 0.948 | 0.030 | 1.034 | 0.117 | 2.474 | 1.346E-02 | |
| 129 | Left_caudalmiddlefrontal_volume | Cortical volume | 0.314 | 0.955 | -0.066 | 1.075 | 0.374 | 7.912 | 4.330E-15 | * |

| | | | | | | | | | | |
|---|---|---|---|---|---|---|---|---|---|---|
| 130 | Left_cuneus_volume | Cortical volume | 0.121 | 0.939 | 0.052 | 0.974 | 0.073 | 1.521 | 1.284E-01 | |
| 131 | Left_entorhinal_volume | Cortical volume | 0.045 | 0.997 | 0.159 | 0.963 | -0.116 | -2.413 | 1.594E-02 | |
| 132 | Left_fusiform_volume | Cortical volume | 0.286 | 1.071 | 0.210 | 1.039 | 0.072 | 1.496 | 1.349E-01 | |
| 133 | Left_inferiorparietal_volume | Cortical volume | 0.267 | 0.995 | 0.066 | 1.016 | 0.200 | 4.188 | 2.943E-05 | * |
| 134 | Left_inferiortemporal_volume | Cortical volume | 0.251 | 1.050 | 0.183 | 1.029 | 0.065 | 1.362 | 1.733E-01 | |
| 135 | Left_isthmuscingulate_volume | Cortical volume | 0.155 | 0.888 | 0.001 | 0.965 | 0.166 | 3.495 | 4.850E-04 | |
| 136 | Left_lateraloccipital_volume | Cortical volume | 0.275 | 0.978 | 0.141 | 1.007 | 0.136 | 2.841 | 4.552E-03 | |
| 137 | Left_lateralorbitofrontal_volume | Cortical volume | 0.368 | 1.003 | 0.009 | 1.046 | 0.350 | 7.347 | 3.036E-13 | * |
| 138 | Left_lingual_volume | Cortical volume | 0.160 | 0.989 | 0.158 | 1.017 | 0.002 | 0.045 | 9.643E-01 | |
| 139 | Left_medialorbitofrontal_volume | Cortical volume | 0.177 | 0.984 | -0.017 | 1.028 | 0.192 | 4.036 | 5.671E-05 | * |
| 140 | Left_middletemporal_volume | Cortical volume | 0.295 | 1.076 | 0.115 | 1.065 | 0.168 | 3.496 | 4.842E-04 | |
| 141 | Left_parahippocampal_volume | Cortical volume | 0.066 | 0.705 | 0.189 | 0.682 | -0.177 | -3.671 | 2.487E-04 | |
| 142 | Left_paracentral_volume | Cortical volume | 0.285 | 0.948 | -0.007 | 1.008 | 0.299 | 6.282 | 4.175E-10 | * |
| 143 | Left_parsopercularis_volume | Cortical volume | 0.275 | 0.976 | -0.037 | 0.994 | 0.316 | 6.614 | 4.903E-11 | * |
| 144 | Left_parsorbitalis_volume | Cortical volume | 0.249 | 1.053 | -0.006 | 1.045 | 0.243 | 5.061 | 4.589E-07 | * |
| 145 | Left_parstriangularis_volume | Cortical volume | 0.265 | 0.952 | -0.042 | 1.031 | 0.309 | 6.511 | 9.589E-11 | * |
| 146 | Left_pericalcarine_volume | Cortical volume | 0.033 | 0.958 | -0.014 | 0.980 | 0.048 | 1.013 | 3.111E-01 | |
| 147 | Left_postcentral_volume | Cortical volume | 0.387 | 0.969 | 0.074 | 1.004 | 0.317 | 6.645 | 3.970E-11 | * |
| 148 | Left_posteriorcingulate_volume | Cortical volume | 0.253 | 0.960 | 0.030 | 1.196 | 0.206 | 4.410 | 1.094E-05 | * |
| 149 | Left_precentral_volume | Cortical volume | 0.415 | 0.968 | 0.098 | 1.009 | 0.320 | 6.710 | 2.584E-11 | * |
| 150 | Left_precuneus_volume | Cortical volume | 0.281 | 0.986 | -0.018 | 1.018 | 0.298 | 6.250 | 5.089E-10 | * |
| 151 | Left_rostralanteriorcingulate_volume | Cortical volume | 0.215 | 0.871 | 0.035 | 0.965 | 0.195 | 4.120 | 3.951E-05 | * |
| 152 | Left_rostralmiddlefrontal_volume | Cortical volume | 0.278 | 0.999 | -0.021 | 0.964 | 0.305 | 6.340 | 2.879E-10 | * |
| 153 | Left_superiorfrontal_volume | Cortical volume | 0.485 | 0.973 | -0.018 | 1.110 | 0.483 | 10.225 | 0.000E+00 | * |
| 154 | Left_superiorparietal_volume | Cortical volume | 0.251 | 1.014 | -0.064 | 1.000 | 0.312 | 6.507 | 9.845E-11 | * |
| 155 | Left_superiortemporal_volume | Cortical volume | 0.333 | 0.993 | 0.190 | 1.063 | 0.140 | 2.942 | 3.306E-03 | |
| 156 | Left_supramarginal_volume | Cortical volume | 0.293 | 0.971 | 0.096 | 0.932 | 0.208 | 4.310 | 1.716E-05 | * |
| 157 | Left_frontalpole_volume | Cortical volume | 0.147 | 1.022 | -0.003 | 1.038 | 0.146 | 3.043 | 2.378E-03 | |
| 158 | Left_temporalpole_volume | Cortical volume | 0.010 | 0.982 | 0.051 | 0.987 | -0.042 | -0.874 | 3.823E-01 | |
| 159 | Left_transversetemporal_volume | Cortical volume | 0.261 | 0.979 | 0.045 | 1.076 | 0.209 | 4.413 | 1.078E-05 | * |
| 160 | Left_insula_volume | Cortical volume | 0.288 | 0.736 | 0.157 | 0.759 | 0.175 | 3.669 | 2.501E-04 | |
| 161 | Right_bankssts_volume | Cortical volume | 0.232 | 1.035 | 0.084 | 1.112 | 0.137 | 2.891 | 3.890E-03 | |
| 162 | Right_caudalanteriorcingulate_volume | Cortical volume | 0.119 | 0.978 | 0.028 | 1.099 | 0.088 | 1.852 | 6.413E-02 | |

| | | | | | | | | | | |
|---|---|---|---|---|---|---|---|---|---|---|
| 163 | Right_caudalmiddlefrontal_volume | Cortical volume | 0.275 | 0.928 | -0.095 | 0.981 | 0.388 | 8.145 | 6.661E-16 | * |
| 164 | Right_cuneus_volume | Cortical volume | 0.160 | 0.917 | 0.062 | 0.951 | 0.104 | 2.187 | 2.889E-02 | |
| 165 | Right_entorhinal_volume | Cortical volume | 0.039 | 0.987 | 0.110 | 0.991 | -0.072 | -1.500 | 1.337E-01 | |
| 166 | Right_fusiform_volume | Cortical volume | 0.285 | 0.993 | 0.217 | 1.026 | 0.067 | 1.413 | 1.578E-01 | |
| 167 | Right_inferiorparietal_volume | Cortical volume | 0.261 | 1.007 | 0.163 | 0.996 | 0.098 | 2.038 | 4.170E-02 | |
| 168 | Right_inferiortemporal_volume | Cortical volume | 0.279 | 1.011 | 0.164 | 1.008 | 0.114 | 2.371 | 1.786E-02 | |
| 169 | Right_isthmuscingulate_volume | Cortical volume | 0.183 | 0.985 | 0.034 | 1.061 | 0.146 | 3.069 | 2.181E-03 | |
| 170 | Right_lateraloccipital_volume | Cortical volume | 0.266 | 0.984 | 0.145 | 1.039 | 0.120 | 2.510 | 1.215E-02 | |
| 171 | Right_lateralorbitofrontal_volume | Cortical volume | 0.323 | 1.064 | 0.000 | 1.077 | 0.301 | 6.299 | 3.747E-10 | * |
| 172 | Right_lingual_volume | Cortical volume | 0.142 | 1.020 | 0.153 | 0.948 | -0.011 | -0.218 | 8.277E-01 | |
| 173 | Right_medialorbitofrontal_volume | Cortical volume | 0.295 | 0.879 | 0.061 | 0.958 | 0.255 | 5.381 | 8.378E-08 | * |
| 174 | Right_middletemporal_volume | Cortical volume | 0.294 | 1.023 | 0.121 | 1.042 | 0.167 | 3.492 | 4.910E-04 | |
| 175 | Right_parahippocampal_volume | Cortical volume | 0.057 | 1.033 | 0.321 | 0.967 | -0.264 | -5.474 | 5.002E-08 | * |
| 176 | Right_paracentral_volume | Cortical volume | 0.321 | 0.973 | 0.036 | 0.976 | 0.293 | 6.120 | 1.143E-09 | * |
| 177 | Right_parsopercularis_volume | Cortical volume | 0.273 | 0.997 | -0.010 | 1.049 | 0.277 | 5.808 | 7.424E-09 | * |
| 178 | Right_parsorbitalis_volume | Cortical volume | 0.212 | 1.061 | -0.001 | 1.114 | 0.196 | 4.104 | 4.244E-05 | * |
| 179 | Right_parstriangularis_volume | Cortical volume | 0.241 | 1.039 | -0.033 | 1.040 | 0.264 | 5.509 | 4.113E-08 | * |
| 180 | Right_pericalcarine_volume | Cortical volume | 0.025 | 0.971 | -0.036 | 0.988 | 0.063 | 1.315 | 1.887E-01 | |
| 181 | Right_postcentral_volume | Cortical volume | 0.328 | 0.993 | 0.010 | 1.017 | 0.317 | 6.625 | 4.539E-11 | * |
| 182 | Right_posteriorcingulate_volume | Cortical volume | 0.312 | 1.017 | 0.051 | 1.319 | 0.222 | 4.764 | 2.050E-06 | * |
| 183 | Right_precentral_volume | Cortical volume | 0.376 | 1.042 | -0.022 | 1.014 | 0.387 | 8.047 | 1.554E-15 | * |
| 184 | Right_precuneus_volume | Cortical volume | 0.314 | 1.043 | -0.014 | 1.048 | 0.314 | 6.553 | 7.290E-11 | * |
| 185 | Right_rostralanteriorcingulate_volume | Cortical volume | 0.169 | 0.992 | 0.108 | 1.055 | 0.060 | 1.255 | 2.096E-01 | |
| 186 | Right_rostralmiddlefrontal_volume | Cortical volume | 0.233 | 1.023 | 0.009 | 0.955 | 0.226 | 4.672 | 3.206E-06 | * |
| 187 | Right_superiorfrontal_volume | Cortical volume | 0.421 | 1.031 | -0.023 | 1.027 | 0.432 | 9.009 | 0.000E+00 | * |
| 188 | Right_superiorparietal_volume | Cortical volume | 0.282 | 1.015 | -0.022 | 0.995 | 0.303 | 6.300 | 3.718E-10 | * |
| 189 | Right_superiortemporal_volume | Cortical volume | 0.323 | 1.097 | 0.158 | 1.093 | 0.150 | 3.136 | 1.741E-03 | |
| 190 | Right_supramarginal_volume | Cortical volume | 0.310 | 1.039 | 0.001 | 1.044 | 0.297 | 6.203 | 6.819E-10 | * |
| 191 | Right_frontalpole_volume | Cortical volume | 0.182 | 1.029 | 0.043 | 1.012 | 0.136 | 2.839 | 4.577E-03 | |
| 192 | Right_temporalpole_volume | Cortical volume | -0.010 | 0.980 | 0.067 | 0.960 | -0.079 | -1.645 | 1.002E-01 | |
| 193 | Right_transversetemporal_volume | Cortical volume | 0.310 | 0.998 | 0.099 | 1.047 | 0.206 | 4.325 | 1.609E-05 | * |
| 194 | Right_insula_volume | Cortical volume | 0.251 | 0.694 | 0.178 | 0.746 | 0.101 | 2.117 | 3.437E-02 | |
| 195 | Left_bankssts_area | Cortical Surface Area | 0.127 | 1.028 | 0.037 | 1.079 | 0.085 | 1.792 | 7.324E-02 | |

| | | | | | | | | | | |
|---|---|---|---|---|---|---|---|---|---|---|
| 196 | Left_caudalanteriorcingulate_area | Cortical Surface Area | 0.141 | 0.921 | -0.016 | 0.997 | 0.164 | 3.449 | 5.764E-04 | |
| 197 | Left_caudalmiddlefrontal_area | Cortical Surface Area | 0.210 | 0.972 | -0.120 | 1.057 | 0.325 | 6.857 | 9.547E-12 | * |
| 198 | Left_cuneus_area | Cortical Surface Area | 0.120 | 0.979 | 0.046 | 0.983 | 0.075 | 1.570 | 1.166E-01 | |
| 199 | Left_entorhinal_area | Cortical Surface Area | 0.059 | 0.922 | 0.105 | 0.887 | -0.052 | -1.070 | 2.846E-01 | |
| 200 | Left_fusiform_area | Cortical Surface Area | 0.177 | 1.083 | 0.119 | 1.057 | 0.054 | 1.128 | 2.595E-01 | |
| 201 | Left_inferiorparietal_area | Cortical Surface Area | 0.162 | 1.033 | -0.001 | 1.042 | 0.156 | 3.266 | 1.111E-03 | |
| 202 | Left_inferiortemporal_area | Cortical Surface Area | 0.137 | 1.065 | 0.058 | 1.111 | 0.072 | 1.517 | 1.295E-01 | |
| 203 | Left_isthmuscingulate_area | Cortical Surface Area | 0.063 | 0.839 | -0.087 | 1.080 | 0.155 | 3.322 | 9.119E-04 | |
| 204 | Left_lateraloccipital_area | Cortical Surface Area | 0.165 | 0.988 | 0.061 | 0.988 | 0.105 | 2.200 | 2.794E-02 | |
| 205 | Left_lateralorbitofrontal_area | Cortical Surface Area | 0.223 | 0.997 | -0.060 | 1.008 | 0.282 | 5.888 | 4.624E-09 | * |
| 206 | Left_lingual_area | Cortical Surface Area | 0.082 | 1.011 | 0.087 | 1.012 | -0.005 | -0.105 | 9.165E-01 | |
| 207 | Left_medialorbitofrontal_area | Cortical Surface Area | 0.096 | 1.025 | -0.051 | 1.060 | 0.141 | 2.956 | 3.157E-03 | |
| 208 | Left_middletemporal_area | Cortical Surface Area | 0.151 | 1.090 | -0.020 | 1.114 | 0.155 | 3.248 | 1.182E-03 | |
| 209 | Left_parahippocampal_area | Cortical Surface Area | 0.006 | 0.698 | 0.132 | 0.571 | -0.197 | -4.020 | 6.056E-05 | * |
| 210 | Left_paracentral_area | Cortical Surface Area | 0.132 | 0.952 | -0.066 | 0.967 | 0.206 | 4.298 | 1.814E-05 | * |
| 211 | Left_parsopercularis_area | Cortical Surface Area | 0.189 | 0.986 | -0.094 | 1.023 | 0.282 | 5.900 | 4.330E-09 | * |
| 212 | Left_parsorbitalis_area | Cortical Surface Area | 0.195 | 1.024 | -0.041 | 1.015 | 0.232 | 4.840 | 1.408E-06 | * |
| 213 | Left_parstriangularis_area | Cortical Surface Area | 0.167 | 0.951 | -0.091 | 1.040 | 0.259 | 5.470 | 5.112E-08 | * |
| 214 | Left_pericalcarine_area | Cortical Surface Area | 0.065 | 0.972 | 0.047 | 0.958 | 0.018 | 0.378 | 7.056E-01 | |
| 215 | Left_postcentral_area | Cortical Surface Area | 0.232 | 1.003 | -0.046 | 0.988 | 0.280 | 5.827 | 6.654E-09 | * |
| 216 | Left_posteriorcingulate_area | Cortical Surface Area | 0.162 | 0.953 | -0.024 | 1.459 | 0.151 | 3.312 | 9.441E-04 | |
| 217 | Left_precentral_area | Cortical Surface Area | 0.220 | 0.977 | 0.000 | 0.990 | 0.224 | 4.682 | 3.054E-06 | * |
| 218 | Left_precuneus_area | Cortical Surface Area | 0.151 | 0.995 | -0.087 | 1.034 | 0.234 | 4.905 | 1.016E-06 | * |
| 219 | Left_rostralanteriorcingulate_area | Cortical Surface Area | 0.197 | 0.904 | 0.020 | 0.947 | 0.192 | 4.026 | 5.895E-05 | * |
| 220 | Left_rostralmiddlefrontal_area | Cortical Surface Area | 0.150 | 1.027 | -0.110 | 1.001 | 0.257 | 5.350 | 9.915E-08 | * |
| 221 | Left_superiorfrontal_area | Cortical Surface Area | 0.284 | 0.994 | -0.113 | 1.070 | 0.384 | 8.084 | 1.110E-15 | * |
| 222 | Left_superiorparietal_area | Cortical Surface Area | 0.131 | 1.039 | -0.120 | 1.016 | 0.244 | 5.072 | 4.341E-07 | * |
| 223 | Left_superiortemporal_area | Cortical Surface Area | 0.212 | 0.991 | -0.013 | 1.074 | 0.217 | 4.570 | 5.199E-06 | * |
| 224 | Left_supramarginal_area | Cortical Surface Area | 0.172 | 0.986 | -0.006 | 0.993 | 0.180 | 3.750 | 1.825E-04 | |
| 225 | Left_frontalpole_area | Cortical Surface Area | 0.065 | 1.027 | -0.078 | 1.038 | 0.139 | 2.897 | 3.818E-03 | |
| 226 | Left_temporalpole_area | Cortical Surface Area | 0.013 | 0.962 | -0.010 | 1.030 | 0.022 | 0.468 | 6.397E-01 | |
| 227 | Left_transversetemporal_area | Cortical Surface Area | 0.193 | 0.989 | -0.065 | 1.090 | 0.248 | 5.223 | 1.962E-07 | * |
| 228 | Left_insula_area | Cortical Surface Area | 0.182 | 0.779 | 0.088 | 0.788 | 0.121 | 2.522 | 1.177E-02 | |

| | | | | | | | | | | |
|---|---|---|---|---|---|---|---|---|---|---|
| 229 | Right_bankssts_area | Cortical Surface Area | 0.180 | 1.028 | 0.010 | 1.107 | 0.159 | 3.343 | 8.458E-04 | |
| 230 | Right_caudalanteriorcingulate_area | Cortical Surface Area | 0.091 | 0.963 | -0.027 | 1.087 | 0.115 | 2.436 | 1.496E-02 | |
| 231 | Right_caudalmiddlefrontal_area | Cortical Surface Area | 0.168 | 0.962 | -0.160 | 0.993 | 0.336 | 7.039 | 2.727E-12 | * |
| 232 | Right_cuneus_area | Cortical Surface Area | 0.137 | 0.937 | 0.015 | 0.929 | 0.130 | 2.718 | 6.630E-03 | |
| 233 | Right_entorhinal_area | Cortical Surface Area | 0.004 | 0.932 | 0.037 | 0.998 | -0.034 | -0.707 | 4.795E-01 | |
| 234 | Right_fusiform_area | Cortical Surface Area | 0.171 | 1.045 | 0.086 | 1.076 | 0.079 | 1.659 | 9.731E-02 | |
| 235 | Right_inferiorparietal_area | Cortical Surface Area | 0.164 | 1.028 | 0.046 | 1.034 | 0.115 | 2.395 | 1.671E-02 | |
| 236 | Right_inferiortemporal_area | Cortical Surface Area | 0.198 | 1.033 | 0.016 | 1.081 | 0.172 | 3.604 | 3.217E-04 | |
| 237 | Right_isthmuscingulate_area | Cortical Surface Area | 0.072 | 1.001 | -0.107 | 1.246 | 0.158 | 3.379 | 7.439E-04 | |
| 238 | Right_lateraloccipital_area | Cortical Surface Area | 0.143 | 0.999 | 0.055 | 1.009 | 0.088 | 1.830 | 6.748E-02 | |
| 239 | Right_lateralorbitofrontal_area | Cortical Surface Area | 0.185 | 1.041 | -0.098 | 1.050 | 0.271 | 5.651 | 1.849E-08 | * |
| 240 | Right_lingual_area | Cortical Surface Area | 0.093 | 1.000 | 0.080 | 0.935 | 0.013 | 0.269 | 7.880E-01 | |
| 241 | Right_medialorbitofrontal_area | Cortical Surface Area | 0.202 | 0.935 | 0.030 | 1.013 | 0.176 | 3.709 | 2.146E-04 | |
| 242 | Right_middletemporal_area | Cortical Surface Area | 0.196 | 1.041 | -0.031 | 1.079 | 0.214 | 4.487 | 7.660E-06 | * |
| 243 | Right_parahippocampal_area | Cortical Surface Area | -0.024 | 1.027 | 0.199 | 0.919 | -0.229 | -4.713 | 2.625E-06 | * |
| 244 | Right_paracentral_area | Cortical Surface Area | 0.153 | 0.983 | -0.028 | 0.976 | 0.184 | 3.845 | 1.246E-04 | * |
| 245 | Right_parsopercularis_area | Cortical Surface Area | 0.180 | 0.996 | -0.062 | 1.039 | 0.238 | 4.985 | 6.793E-07 | * |
| 246 | Right_parsorbitalis_area | Cortical Surface Area | 0.139 | 1.024 | -0.044 | 1.042 | 0.177 | 3.698 | 2.234E-04 | |
| 247 | Right_parstriangularis_area | Cortical Surface Area | 0.164 | 1.027 | -0.075 | 1.011 | 0.235 | 4.885 | 1.126E-06 | * |
| 248 | Right_pericalcarine_area | Cortical Surface Area | 0.094 | 0.949 | 0.023 | 0.909 | 0.076 | 1.582 | 1.137E-01 | |
| 249 | Right_postcentral_area | Cortical Surface Area | 0.211 | 1.045 | -0.101 | 1.028 | 0.301 | 6.272 | 4.419E-10 | * |
| 250 | Right_posteriorcingulate_area | Cortical Surface Area | 0.201 | 1.011 | -0.038 | 1.524 | 0.185 | 4.043 | 5.488E-05 | * |
| 251 | Right_precentral_area | Cortical Surface Area | 0.197 | 0.991 | -0.100 | 1.030 | 0.294 | 6.165 | 8.625E-10 | * |
| 252 | Right_precuneus_area | Cortical Surface Area | 0.179 | 1.039 | -0.095 | 1.050 | 0.262 | 5.469 | 5.140E-08 | * |
| 253 | Right_rostralanteriorcingulate_area | Cortical Surface Area | 0.178 | 0.967 | 0.086 | 0.992 | 0.093 | 1.952 | 5.111E-02 | |
| 254 | Right_rostralmiddlefrontal_area | Cortical Surface Area | 0.134 | 1.063 | -0.064 | 0.994 | 0.192 | 3.970 | 7.460E-05 | * |
| 255 | Right_superiorfrontal_area | Cortical Surface Area | 0.229 | 1.063 | -0.107 | 1.014 | 0.323 | 6.706 | 2.654E-11 | * |
| 256 | Right_superiorparietal_area | Cortical Surface Area | 0.164 | 1.052 | -0.087 | 0.999 | 0.245 | 5.083 | 4.087E-07 | * |
| 257 | Right_superiortemporal_area | Cortical Surface Area | 0.184 | 1.092 | -0.019 | 1.084 | 0.187 | 3.891 | 1.036E-04 | * |
| 258 | Right_supramarginal_area | Cortical Surface Area | 0.192 | 1.014 | -0.090 | 1.029 | 0.277 | 5.779 | 8.803E-09 | * |
| 259 | Right_frontalpole_area | Cortical Surface Area | 0.114 | 1.018 | -0.028 | 1.025 | 0.139 | 2.907 | 3.697E-03 | |
| 260 | Right_temporalpole_area | Cortical Surface Area | -0.052 | 0.984 | -0.043 | 0.958 | -0.010 | -0.198 | 8.430E-01 | |
| 261 | Right_transversetemporal_area | Cortical Surface Area | 0.202 | 0.994 | -0.049 | 1.030 | 0.248 | 5.205 | 2.159E-07 | * |

| | | | | | | | | | | |
|---|---|---|---|---|---|---|---|---|---|---|
| 262 | Right_insula_area | Cortical Surface Area | 0.141 | 0.689 | 0.101 | 0.737 | 0.056 | 1.182 | 2.373E-01 | |
| 263 | Left_bankssts_thickness | Cortical Thickness | 0.179 | 1.088 | 0.210 | 1.069 | -0.029 | -0.594 | 5.524E-01 | |
| 264 | Left_caudalanteriorcingulate_thickness | Cortical Thickness | 0.048 | 1.016 | 0.126 | 0.977 | -0.078 | -1.616 | 1.063E-01 | |
| 265 | Left_caudalmiddlefrontal_thickness | Cortical Thickness | 0.362 | 1.056 | 0.215 | 1.082 | 0.138 | 2.879 | 4.037E-03 | |
| 266 | Left_cuneus_thickness | Cortical Thickness | 0.079 | 0.962 | 0.027 | 1.044 | 0.052 | 1.093 | 2.747E-01 | |
| 267 | Left_entorhinal_thickness | Cortical Thickness | 0.069 | 0.954 | 0.091 | 0.954 | -0.023 | -0.474 | 6.352E-01 | |
| 268 | Left_fusiform_thickness | Cortical Thickness | 0.318 | 1.058 | 0.344 | 1.059 | -0.025 | -0.523 | 6.009E-01 | |
| 269 | Left_inferiorparietal_thickness | Cortical Thickness | 0.311 | 1.086 | 0.213 | 1.099 | 0.090 | 1.881 | 6.008E-02 | |
| 270 | Left_inferiortemporal_thickness | Cortical Thickness | 0.310 | 1.021 | 0.366 | 0.991 | -0.055 | -1.151 | 2.499E-01 | |
| 271 | Left_isthmuscingulate_thickness | Cortical Thickness | 0.181 | 0.976 | 0.122 | 0.954 | 0.062 | 1.285 | 1.991E-01 | |
| 272 | Left_lateraloccipital_thickness | Cortical Thickness | 0.216 | 1.061 | 0.153 | 1.094 | 0.058 | 1.210 | 2.265E-01 | |
| 273 | Left_lateralorbitofrontal_thickness | Cortical Thickness | 0.277 | 1.012 | 0.217 | 1.033 | 0.059 | 1.239 | 2.155E-01 | |
| 274 | Left_lingual_thickness | Cortical Thickness | 0.201 | 1.037 | 0.163 | 1.049 | 0.036 | 0.753 | 4.515E-01 | |
| 275 | Left_medialorbitofrontal_thickness | Cortical Thickness | 0.161 | 1.004 | 0.119 | 0.999 | 0.041 | 0.863 | 3.883E-01 | |
| 276 | Left_middletemporal_thickness | Cortical Thickness | 0.319 | 1.148 | 0.315 | 1.145 | 0.004 | 0.078 | 9.382E-01 | |
| 277 | Left_parahippocampal_thickness | Cortical Thickness | 0.109 | 0.983 | 0.157 | 1.004 | -0.048 | -1.002 | 3.164E-01 | |
| 278 | Left_paracentral_thickness | Cortical Thickness | 0.299 | 1.056 | 0.097 | 1.095 | 0.188 | 3.939 | 8.504E-05 | * |
| 279 | Left_parsopercularis_thickness | Cortical Thickness | 0.341 | 1.064 | 0.222 | 1.022 | 0.114 | 2.364 | 1.819E-02 | |
| 280 | Left_parsorbitalis_thickness | Cortical Thickness | 0.227 | 0.958 | 0.137 | 1.075 | 0.089 | 1.875 | 6.097E-02 | |
| 281 | Left_parstriangularis_thickness | Cortical Thickness | 0.327 | 0.954 | 0.174 | 1.021 | 0.154 | 3.246 | 1.193E-03 | |
| 282 | Left_pericalcarine_thickness | Cortical Thickness | -0.014 | 0.980 | -0.050 | 0.964 | 0.037 | 0.776 | 4.379E-01 | |
| 283 | Left_postcentral_thickness | Cortical Thickness | 0.300 | 1.067 | 0.236 | 1.012 | 0.061 | 1.272 | 2.036E-01 | |
| 284 | Left_posteriorcingulate_thickness | Cortical Thickness | 0.194 | 0.989 | 0.115 | 1.093 | 0.076 | 1.606 | 1.085E-01 | |
| 285 | Left_precentral_thickness | Cortical Thickness | 0.335 | 1.104 | 0.163 | 0.986 | 0.164 | 3.384 | 7.297E-04 | |
| 286 | Left_precuneus_thickness | Cortical Thickness | 0.308 | 1.046 | 0.158 | 1.097 | 0.140 | 2.927 | 3.468E-03 | |
| 287 | Left_rostralanteriorcingulate_thickness | Cortical Thickness | 0.046 | 0.972 | 0.111 | 0.952 | -0.068 | -1.414 | 1.576E-01 | |
| 288 | Left_rostralmiddlefrontal_thickness | Cortical Thickness | 0.345 | 1.004 | 0.264 | 1.033 | 0.079 | 1.658 | 9.751E-02 | |
| 289 | Left_superiorfrontal_thickness | Cortical Thickness | 0.408 | 1.036 | 0.255 | 1.076 | 0.145 | 3.043 | 2.375E-03 | |
| 290 | Left_superiorparietal_thickness | Cortical Thickness | 0.276 | 1.071 | 0.139 | 1.091 | 0.127 | 2.650 | 8.113E-03 | |
| 291 | Left_superiortemporal_thickness | Cortical Thickness | 0.274 | 1.094 | 0.397 | 1.112 | -0.112 | -2.336 | 1.962E-02 | |
| 292 | Left_supramarginal_thickness | Cortical Thickness | 0.348 | 1.089 | 0.275 | 1.092 | 0.067 | 1.404 | 1.604E-01 | |
| 293 | Left_frontalpole_thickness | Cortical Thickness | 0.193 | 1.002 | 0.141 | 1.023 | 0.051 | 1.065 | 2.870E-01 | |
| 294 | Left_temporalpole_thickness | Cortical Thickness | 0.024 | 0.971 | 0.133 | 1.002 | -0.111 | -2.316 | 2.064E-02 | |

| | | | | | | | | | |
|---|---|---|---|---|---|---|---|---|---|
| 295 | Left_transversetemporal_thickness | Cortical Thickness | 0.162 | 1.028 | 0.200 | 1.037 | -0.037 | -0.766 | 4.439E-01 | |
| 296 | Left_insula_thickness | Cortical Thickness | 0.225 | 0.984 | 0.148 | 1.029 | 0.077 | 1.614 | 1.066E-01 | |
| 297 | Right_bankssts_thickness | Cortical Thickness | 0.151 | 0.974 | 0.192 | 1.052 | -0.040 | -0.848 | 3.966E-01 | |
| 298 | Right_caudalanteriorcingulate_thickness | Cortical Thickness | 0.037 | 0.996 | 0.134 | 1.031 | -0.096 | -2.009 | 4.468E-02 | |
| 299 | Right_caudalmiddlefrontal_thickness | Cortical Thickness | 0.321 | 1.068 | 0.203 | 1.114 | 0.108 | 2.263 | 2.374E-02 | |
| 300 | Right_cuneus_thickness | Cortical Thickness | 0.103 | 0.973 | 0.065 | 1.010 | 0.039 | 0.821 | 4.119E-01 | |
| 301 | Right_entorhinal_thickness | Cortical Thickness | 0.094 | 0.974 | 0.100 | 1.017 | -0.006 | -0.129 | 8.971E-01 | |
| 302 | Right_fusiform_thickness | Cortical Thickness | 0.313 | 1.028 | 0.409 | 1.000 | -0.094 | -1.965 | 4.959E-02 | |
| 303 | Right_inferiorparietal_thickness | Cortical Thickness | 0.236 | 1.062 | 0.262 | 1.052 | -0.024 | -0.506 | 6.126E-01 | |
| 304 | Right_inferiortemporal_thickness | Cortical Thickness | 0.272 | 1.096 | 0.351 | 1.058 | -0.074 | -1.537 | 1.246E-01 | |
| 305 | Right_isthmuscingulate_thickness | Cortical Thickness | 0.194 | 0.976 | 0.210 | 0.990 | -0.016 | -0.332 | 7.402E-01 | |
| 306 | Right_lateraloccipital_thickness | Cortical Thickness | 0.210 | 1.005 | 0.154 | 1.052 | 0.054 | 1.139 | 2.549E-01 | |
| 307 | Right_lateralorbitofrontal_thickness | Cortical Thickness | 0.232 | 0.986 | 0.233 | 1.040 | -0.001 | -0.017 | 9.862E-01 | |
| 308 | Right_lingual_thickness | Cortical Thickness | 0.139 | 0.973 | 0.166 | 0.967 | -0.028 | -0.588 | 5.563E-01 | |
| 309 | Right_medialorbitofrontal_thickness | Cortical Thickness | 0.181 | 0.998 | 0.108 | 1.040 | 0.072 | 1.506 | 1.323E-01 | |
| 310 | Right_middletemporal_thickness | Cortical Thickness | 0.222 | 1.084 | 0.314 | 1.150 | -0.083 | -1.734 | 8.304E-02 | |
| 311 | Right_parahippocampal_thickness | Cortical Thickness | 0.121 | 1.022 | 0.234 | 1.028 | -0.110 | -2.300 | 2.153E-02 | |
| 312 | Right_paracentral_thickness | Cortical Thickness | 0.333 | 1.085 | 0.142 | 1.075 | 0.177 | 3.687 | 2.335E-04 | |
| 313 | Right_parsopercularis_thickness | Cortical Thickness | 0.340 | 1.037 | 0.192 | 1.032 | 0.143 | 2.976 | 2.956E-03 | |
| 314 | Right_parsorbitalis_thickness | Cortical Thickness | 0.216 | 1.040 | 0.168 | 1.092 | 0.046 | 0.956 | 3.394E-01 | |
| 315 | Right_parstriangularis_thickness | Cortical Thickness | 0.273 | 1.017 | 0.169 | 1.149 | 0.096 | 2.033 | 4.218E-02 | |
| 316 | Right_pericalcarine_thickness | Cortical Thickness | -0.068 | 0.964 | -0.038 | 1.017 | -0.031 | -0.645 | 5.187E-01 | |
| 317 | Right_postcentral_thickness | Cortical Thickness | 0.193 | 1.065 | 0.182 | 0.992 | 0.011 | 0.218 | 8.275E-01 | |
| 318 | Right_posteriorcingulate_thickness | Cortical Thickness | 0.221 | 0.965 | 0.202 | 1.072 | 0.018 | 0.385 | 7.003E-01 | |
| 319 | Right_precentral_thickness | Cortical Thickness | 0.299 | 1.125 | 0.101 | 1.007 | 0.186 | 3.825 | 1.349E-04 | * |
| 320 | Right_precuneus_thickness | Cortical Thickness | 0.325 | 1.034 | 0.170 | 1.083 | 0.146 | 3.072 | 2.156E-03 | |
| 321 | Right_rostralanteriorcingulate_thickness | Cortical Thickness | 0.000 | 1.082 | 0.001 | 1.106 | -0.001 | -0.026 | 9.795E-01 | |
| 322 | Right_rostralmiddlefrontal_thickness | Cortical Thickness | 0.265 | 1.036 | 0.245 | 1.036 | 0.019 | 0.405 | 6.858E-01 | |
| 323 | Right_superiorfrontal_thickness | Cortical Thickness | 0.407 | 1.051 | 0.250 | 1.070 | 0.149 | 3.114 | 1.874E-03 | |
| 324 | Right_superiorparietal_thickness | Cortical Thickness | 0.276 | 1.034 | 0.159 | 1.062 | 0.112 | 2.337 | 1.957E-02 | |
| 325 | Right_superiortemporal_thickness | Cortical Thickness | 0.271 | 1.075 | 0.319 | 1.090 | -0.045 | -0.941 | 3.468E-01 | |
| 326 | Right_supramarginal_thickness | Cortical Thickness | 0.299 | 1.098 | 0.240 | 1.135 | 0.053 | 1.108 | 2.679E-01 | |
| 327 | Right_frontalpole_thickness | Cortical Thickness | 0.192 | 1.008 | 0.173 | 1.017 | 0.019 | 0.401 | 6.882E-01 | |

| | | | | | | | | | |
|---|---|---|---|---|---|---|---|---|---|
| 328 | Right_temporalpole_thickness | Cortical Thickness | 0.047 | 0.979 | 0.173 | 1.036 | -0.124 | -2.612 | 9.074E-03 |
| 329 | Right_transversetemporal_thickness | Cortical Thickness | 0.216 | 1.012 | 0.231 | 1.056 | -0.015 | -0.311 | 7.555E-01 |
| 330 | Right_insula_thickness | Cortical Thickness | 0.273 | 0.983 | 0.189 | 1.008 | 0.084 | 1.748 | 8.062E-02 |

**Supplementary Table S6. Difference of volume of striatum between the subtype1 and subtype2 in a medication-naïve subsample.**

| Region | Morphological measure | Subtype1(n=445) | | Subtype2(n=309) | | Subtype1 vs. Subtype2 | | |
|---|---|---|---|---|---|---|---|---|
| | | Mean | SD | Mean | SD | Cohen's d | T | P |
| Left_Caudate | Subcortical volume | -0.284 | 1.003 | 0.299 | 1.089 | -0.557 | -7.617 | 0.000 |
| Left_Putamen | Subcortical volume | -0.197 | 1.069 | 0.230 | 1.056 | -0.402 | -5.442 | 0.000 |
| Right_Caudate | Subcortical volume | -0.274 | 1.048 | 0.251 | 1.148 | -0.478 | -6.538 | 0.000 |
| Right_Putamen | Subcortical volume | -0.192 | 1.025 | 0.203 | 1.084 | -0.374 | -5.104 | 0.000 |

**Supplementary Table 7. SuStaIn features.**

| Feature ID | SuStaIn Feature | AAL3 ROI Name | AAL3 ROI Full Name |
|---|---|---|---|
| 1 | Hippocampus | lHIP | Left Hippocampus |
| 1 | Hippocampus | rHIP | Right Hippocampus |
| 2 | Parahippocampus | lPHG | Left Parahippocampal gyrus |
| 2 | Parahippocampus | rPHG | Right Parahippocampal gyrus |
| 3 | Amygdala | lAMYG | Left Amygdala |
| 3 | Amygdala | rAMYG | Right Amygdala |
| 4 | Caudate | lCAU | Left Caudate nucleus |
| 4 | Caudate | rCAU | Right Caudate nucleus |
| 5 | Putamen | lPUT | Left Lenticular nucleus-Putamen |
| 5 | Putamen | rPUT | Right Lenticular nucleus-Putamen |
| 6 | Pallidum | lPAL | Left Lenticular nucleus-Pallidum |
| 6 | Pallidum | rPAL | Right Lenticular nucleus-Pallidum |
| 7 | Thalamus | ltAV | Left Thalamus-Anteroventral Nucleus |
| 7 | Thalamus | rtAV | Right Thalamus-Anteroventral Nucleus |
| 7 | Thalamus | ltLP | Left Lateral posterior |
| 7 | Thalamus | rtLP | Right Lateral posterior |
| 7 | Thalamus | ltVA | Left Ventral anterior |
| 7 | Thalamus | rtVA | Right Ventral anterior |
| 7 | Thalamus | ltVL | Left Ventral lateral |
| 7 | Thalamus | rtVL | Right Ventral lateral |
| 7 | Thalamus | ltVPL | Left Ventral posterolateral |
| 7 | Thalamus | rtVPL | Right Ventral posterolateral |
| 7 | Thalamus | ltIL | Left Intralaminar |
| 7 | Thalamus | rtIL | Right Intralaminar |
| 7 | Thalamus | ltMDm | Left Mediodorsal medial magnocellular |
| 7 | Thalamus | rtMDm | Right Mediodorsal medial magnocellular |
| 7 | Thalamus | ltMDl | Left Mediodorsal lateral parvocellular |
| 7 | Thalamus | rtMDl | Right Mediodorsal lateral parvocellular |
| 7 | Thalamus | ltLGN | Left Lateral geniculate |
| 7 | Thalamus | rtLGN | Right Lateral geniculate |
| 7 | Thalamus | ltMGN | Left Medial Geniculate |

| | | | |
|---|---|---|---|
| 7 | Thalamus | rtMGN | Right Medial Geniculate |
| 7 | Thalamus | ltPuA | Left Pulvinar anterior |
| 7 | Thalamus | rtPuA | Right Pulvinar anterior |
| 7 | Thalamus | ltPuM | Left Pulvinar medial |
| 7 | Thalamus | rtPuM | Right Pulvinar medial |
| 7 | Thalamus | ltPuL | Left Pulvinar lateral |
| 7 | Thalamus | rtPuL | Right Pulvinar lateral |
| 7 | Thalamus | ltPuI | Left Pulvinar inferior |
| 7 | Thalamus | rtPuI | Right Pulvinar inferior |
| 8 | Accumbens | lNacc | Left Nucleus accumbens |
| 8 | Accumbens | rNacc | Right Nucleus accumbens |
| 9 | Cingulate | lMCC | Left Middle cingulate & paracingulate gyri |
| 9 | Cingulate | rMCC | Right Middle cingulate & paracingulate gyri |
| 9 | Cingulate | lPCC | Left Posterior cingulate gyrus |
| 9 | Cingulate | rPCC | Right Posterior cingulate gyrus |
| 9 | Cingulate | lACCsub | Left Anterior cingulate cortex-subgenual |
| 9 | Cingulate | rACCsub | Right Anterior cingulate cortex-subgenual |
| 9 | Cingulate | lACCpre | Left Anterior cingulate cortex-pregenual |
| 9 | Cingulate | rACCpre | Right Anterior cingulate cortex-pregenual |
| 9 | Cingulate | lACCsup | Left Anterior cingulate cortex-supracallosal |
| 9 | Cingulate | rACCsup | Right Anterior cingulate cortex-supracallosal |
| 10 | Frontal Cortex | lSFG | Left Superior frontal gyrus-dorsolateral |
| 10 | Frontal Cortex | rSFG | Right Superior frontal gyrus-dorsolateral |
| 10 | Frontal Cortex | lMFG | Left Middle frontal gyrus |
| 10 | Frontal Cortex | rMFG | Right Middle frontal gyrus |
| 10 | Frontal Cortex | lIFGorb | Left IFG pars orbitalis |
| 10 | Frontal Cortex | rIFGorb | Right IFG pars orbitalis |
| 10 | Frontal Cortex | lROL | Left Rolandic operculum |
| 10 | Frontal Cortex | rROL | Right Rolandic operculum |
| 10 | Frontal Cortex | lOLF | Left Olfactory cortex |
| 10 | Frontal Cortex | rOLF | Right Olfactory cortex |
| 10 | Frontal Cortex | lSFGmedial | Left Superior frontal gyrus-medial |
| 10 | Frontal Cortex | rSFGmedial | Right Superior frontal gyrus-medial |

| | | | |
|---|---|---|---|
| 10 | Frontal Cortex | lPFCventmed | Left Superior frontal gyrus-medial orbital |
| 10 | Frontal Cortex | rPFCventmed | Right Superior frontal gyrus-medial orbital |
| 10 | Frontal Cortex | lREC | Left Gyrus rectus |
| 10 | Frontal Cortex | rREC | Right Gyrus rectus |
| 10 | Frontal Cortex | lOFCmed | Left Medial orbital gyrus |
| 10 | Frontal Cortex | rOFCmed | Right Medial orbital gyrus |
| 10 | Frontal Cortex | lOFCant | Left Anterior orbital gyrus |
| 10 | Frontal Cortex | rOFCant | Right Anterior orbital gyrus |
| 10 | Frontal Cortex | lOFCpost | Left Posterior orbital gyrus |
| 10 | Frontal Cortex | rOFCpost | Right Posterior orbital gyrus |
| 10 | Frontal Cortex | lOFClat | Left Lateral orbital gyrus |
| 10 | Frontal Cortex | rOFClat | Right Lateral orbital gyrus |
| 11 | Parietal Cortex | lSPG | Left Superior parietal gyrus |
| 11 | Parietal Cortex | rSPG | Right Superior parietal gyrus |
| 11 | Parietal Cortex | lIPG | Left Inferior parietal gyrus-excluding supramarginal and angular gyri |
| 11 | Parietal Cortex | rIPG | Right Inferior parietal gyrus-excluding supramarginal and angular gyri |
| 11 | Parietal Cortex | lSMG | Left SupraMarginal gyrus |
| 11 | Parietal Cortex | rSMG | Right SupraMarginal gyrus |
| 11 | Parietal Cortex | lANG | Left Angular gyrus |
| 11 | Parietal Cortex | rANG | Right Angular gyrus |
| 11 | Parietal Cortex | lPCUN | Left Precuneus |
| 11 | Parietal Cortex | rPCUN | Right Precuneus |
| 11 | Parietal Cortex | lPCL | Left Paracentral lobule |
| 11 | Parietal Cortex | rPCL | Right Paracentral lobule |
| 12 | Temporal Cortex | lFFG | Left Fusiform gyrus |
| 12 | Temporal Cortex | rFFG | Right Fusiform gyrus |
| 12 | Temporal Cortex | lHES | Left Heschls gyrus |
| 12 | Temporal Cortex | rHES | Right Heschls gyrus |
| 12 | Temporal Cortex | lSTG | Left Superior temporal gyrus |
| 12 | Temporal Cortex | rSTG | Right Superior temporal gyrus |
| 12 | Temporal Cortex | lTPOsup | Left Temporal pole: superior temporal gyrus |
| 12 | Temporal Cortex | rTPOsup | Right Temporal pole: superior temporal gyrus |
| 12 | Temporal Cortex | lMTG | Left Middle temporal gyrus |

| | | | |
|---|---|---|---|
| 12 | Temporal Cortex | rMTG | Right Middle temporal gyrus |
| 12 | Temporal Cortex | lTPOmid | Left Temporal pole: middle temporal gyrus |
| 12 | Temporal Cortex | rTPOmid | Right Temporal pole: middle temporal gyrus |
| 12 | Temporal Cortex | lITG | Left Inferior temporal gyrus |
| 12 | Temporal Cortex | rITG | Right Inferior temporal gyrus |
| 13 | Occipital Cortex | lCAL | Left Calcarine fissure and surrounding cortex |
| 13 | Occipital Cortex | rCAL | Right Calcarine fissure and surrounding cortex |
| 13 | Occipital Cortex | lCUN | Left Cuneus |
| 13 | Occipital Cortex | rCUN | Right Cuneus |
| 13 | Occipital Cortex | lLING | Left Lingual gyrus |
| 13 | Occipital Cortex | rLING | Right Lingual gyrus |
| 13 | Occipital Cortex | lSOG | Left Superior occipital gyrus |
| 13 | Occipital Cortex | rSOG | Right Superior occipital gyrus |
| 13 | Occipital Cortex | lMOG | Left Middle occipital gyrus |
| 13 | Occipital Cortex | rMOG | Right Middle occipital gyrus |
| 13 | Occipital Cortex | lIOG | Left Inferior occipital gyrus |
| 13 | Occipital Cortex | rIOG | Right Inferior occipital gyrus |
| 14 | Insula | lINS | Left Insula |
| 14 | Insula | rINS | Right Insula |
| 15 | Cerebellum | lCERCRU1 | Left Crus I of cerebellar hemisphere |
| 15 | Cerebellum | rCERCRU1 | Right Crus I of cerebellar hemisphere |
| 15 | Cerebellum | lCERCRU2 | Left Crus II of cerebellar hemisphere |
| 15 | Cerebellum | rCERCRU2 | Right Crus II of cerebellar hemisphere |
| 15 | Cerebellum | lCER3 | Left Lobule III of cerebellar hemisphere |
| 15 | Cerebellum | rCER3 | Right Lobule III of cerebellar hemisphere |
| 15 | Cerebellum | lCER4_5 | Left Lobule IV-V of cerebellar hemisphere |
| 15 | Cerebellum | rCER4_5 | Right Lobule IV-V of cerebellar hemisphere |
| 15 | Cerebellum | lCER6 | Left Lobule VI of cerebellar hemisphere |
| 15 | Cerebellum | rCER6 | Right Lobule VI of cerebellar hemisphere |
| 15 | Cerebellum | lCER7b | Left Lobule VIIB of cerebellar hemisphere |
| 15 | Cerebellum | rCER7b | Right Lobule VIIB of cerebellar hemisphere |
| 15 | Cerebellum | lCER8 | Left Lobule VIII of cerebellar hemisphere |
| 15 | Cerebellum | rCER8 | Right Lobule VIII of cerebellar hemisphere |

| | | | |
|---|---|---|---|
| 15 | Cerebellum | lCER9 | Left Lobule IX of cerebellar hemisphere |
| 15 | Cerebellum | rCER9 | Right Lobule IX of cerebellar hemisphere |
| 15 | Cerebellum | lCER10 | Left Lobule X of cerebellar hemisphere |
| 15 | Cerebellum | rCER10 | Right Lobule X of cerebellar hemisphere |
| 15 | Cerebellum | VER1_2 | Lobule I-II of vermis |
| 15 | Cerebellum | VER3 | Lobule III of vermis |
| 15 | Cerebellum | VER4_5 | Lobule IV-V of vermis |
| 15 | Cerebellum | VER6 | Lobule VI of vermis |
| 15 | Cerebellum | VER7 | Lobule VII of vermis |
| 15 | Cerebellum | VER8 | Lobule VIII of vermis |
| 15 | Cerebellum | VER9 | Lobule IX of vermis |
| 15 | Cerebellum | VER10 | Lobule X of vermis |
| 16 | Sensorimotor | lPreCG | Left Precentral gyrus |
| 16 | Sensorimotor | rPreCG | Right Precentral gyrus |
| 16 | Sensorimotor | lSMA | Left Supplementary motor area |
| 16 | Sensorimotor | rSMA | Right Supplementary motor area |
| 16 | Sensorimotor | lPoCG | Left Postcentral gyrus |
| 16 | Sensorimotor | rPoCG | Right Postcentral gyrus |
| 17 | Broca'area | lIFGoperc | Left Inferior frontal gyrus-opercular part |
| 17 | Broca'area | rIFGoperc | Right Inferior frontal gyrus-opercular part |
| 17 | Broca'area | lIFGtriang | Left Inferior frontal gyrus-triangular part |
| 17 | Broca'area | rIFGtriang | Right Inferior frontal gyrus-triangular part |
